# Transmission dynamics of SARS-CoV-2 in a strictly-Orthodox Jewish community in the UK

**DOI:** 10.1101/2021.10.28.21265615

**Authors:** William Waites, Carl AB Pearson, Katherine M Gaskell, Thomas House, Lorenzo Pellis, Marina Johnson, Victoria Gould, Adam Hunt, Neil RH Stone, Ben Kasstan, Tracey Chantler, Sham Lal, Chrissy h. Roberts, David Goldblatt, CMMID COVID-19 Working Group, Michael Marks, Rosalind M Eggo

## Abstract

Some social settings such as households and workplaces, have been identified as high risk for SARS-CoV-2 transmission. Identifying and quantifying the importance of these settings is critical for designing interventions. A tightly-knit religious community in the UK experienced a very large COVID-19 epidemic in 2020, reaching 64.3% seroprevalence within 10 months, and we surveyed this community both for serological status and individual-level attendance at particular settings. Using these data, and a network model of people and places represented as a stochastic graph rewriting system, we estimated the relative contribution of transmission in households, schools and religious institutions to the epidemic, and the relative risk of infection in each of these settings. All congregate settings were important for transmission, with some such as primary schools and places of worship having a higher share of transmission than others. We found that the model needed a higher general-community transmission rate for women (3.3-fold), and lower susceptibility to infection in children to recreate the observed serological data. The precise share of transmission in each place was related to assumptions about the internal structure of those places. Identification of key settings of transmission can allow public health interventions to be targeted at these locations.

## 1 Introduction

The transmission dynamics of SARS-CoV-2 in settings such as households (1–5), schools (6–8) and workplaces (9, 10) has been the subject of considerable interest, since understanding the relative risk of transmission by setting (11–15) enables more effective targeting of public health interventions (16–22) to minimise the extent and impact of the epidemic. Characterisation of transmission dynamics and evaluation of interventions is often done with agent-based or network models where the network structure is either generated synthetically (23, 24) or inferred from mobility data (25, 26). However, because dynamics are formulated in terms of interactions between individuals, the role of setting is implicit and can only be measured indirectly.

There is a need for further investigation into the importance of different transmission settings. Ideally information on attendance at those settings would be coupled with evidence of infection, to allow the relative importance of each setting to be disentangled.

We developed a transmission model where setting is explicit to examine the role of differing types of places and their relative contribution to SARS-CoV-2 transmission in a strictly-Orthodox Jewish community in the UK.

We previously documented (27) 64.3% (95% CI 61.6-67.0%) SARS-CoV-2 seroprevalence in November 2020 in this community which was more than five times the estimated seroprevalence of the wider metropolitan area (28). Because we collected data on attendance at community institutions from all members of 394 households, approximately 10% of the total community’s population. These data afford a unique opportunity to estimate the relative contributions of different settings to SARS-CoV-2 transmission and to understand what dynamics could have given rise to the particular pattern of seroprevalence observed.

To analyse transmission, we represented the community as a bipartite network of people and places. We constructed a transmission model using an extended version of the *kappa*-calculus (29) to implement a transmission model as a stochastic graph rewriting system (30, 31), a generalalisation of how explicit epidemic dynamics are usually formulated on networks (32). In this model, individuals have disease progression states and transmission is mediated by place, with a separate transmission process for each setting or kind of place. These place-mediated transmission processes are augmented with population-wide well-mixed transmission processes akin to general community transmission outside of the defined set of places. We fit the transmission rate parameters of this model to the measured distributions of positive test results from the seroprevalence survey. We used the fitted model to analyse the contribution of different places to transmission within the community.

## 2 Results

### 2.1 Structure of the social network of people and places

We surveyed 1,942 people in 374 households from a community of approximately 20,000 people in November and early December 2020. 33% of the population were under 10 years of age and 60% under 20, which which is a higher percentage than the surrounding metropolitan area. Survey data included household membership and composition, which school children attended, which place of worship individuals attended, and which ritual bath adult men attended (adult men attend ritual baths collectively, women attend individually but no specific data were available about the latter). 1,377 people from 309 households also provided a blood sample from which we found 64.3% had IgG antibodies to the SARS-CoV-2 spike protein, ranging from 50% in under 10 year olds to 75% in over 10 year olds (27).

We observed a strong partitioning of attendance at places by both age and sex (Figure 1 B,C). Schools, both primary (under 13) and secondary (13 and over), were predominantly segregated by sex. The vast majority of individuals reporting a connection with a place of worship and all of those reporting attendance at a ritual bath were adult (18+) men.

**Figure 1:**
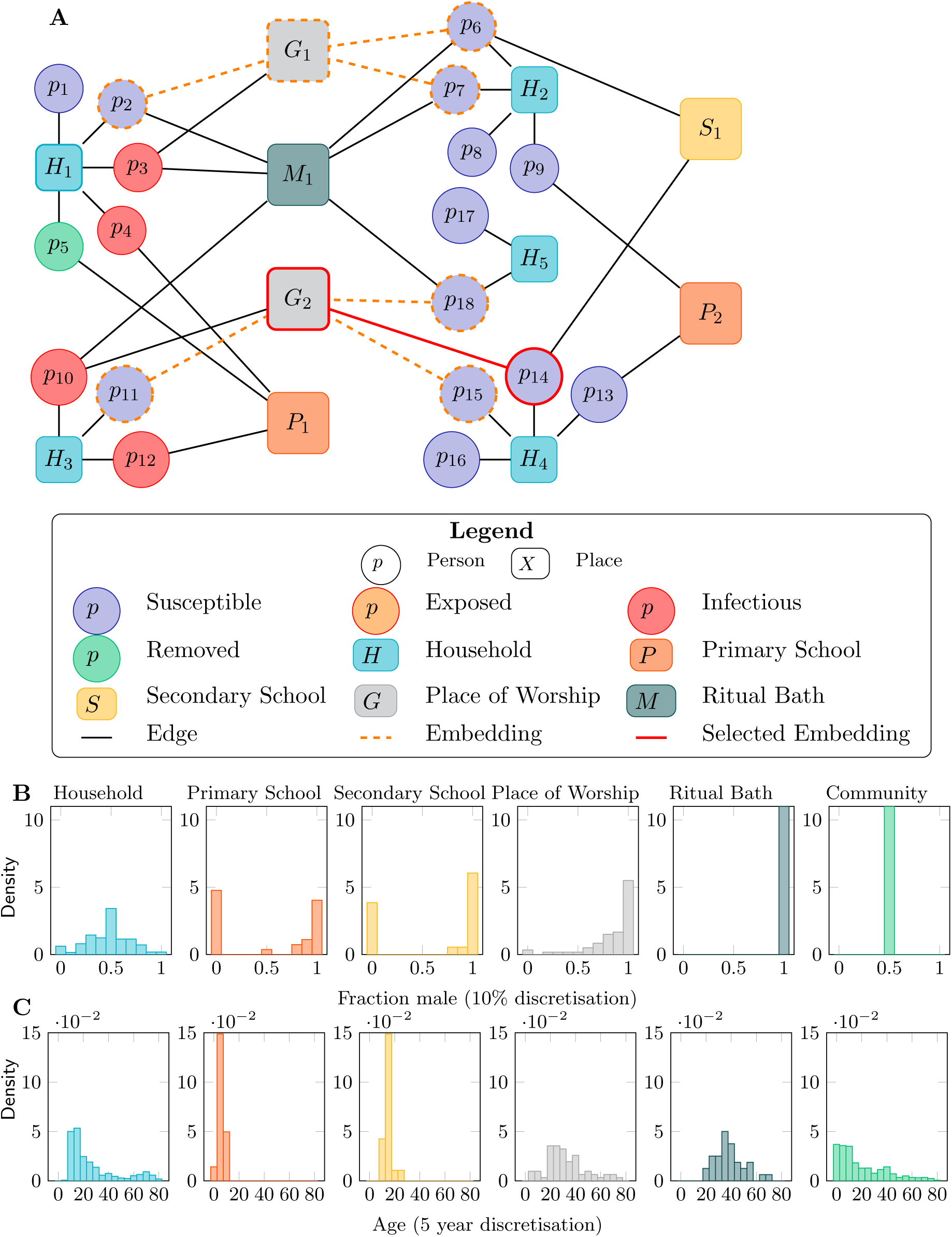
The data and formulation of the people and places network. **A**. Illustrative representation of a bipartite people and places network. Circles represent people and squares represent places. All people are connected to a household (H). Some people are connected to primary (P) or secondary (S) schools, places of worship (G) or ritual baths (M). The orange dotted, and red solid edges represent embeddings of a transmission rule (Equation 6) capturing the situation immediately before a transmission event that will result in the individual p14 becoming infected. **B**. The distribution of the fraction of individuals in each setting who were male. Households were mixed, but the attendees of both primary and secondary schools were strongly bimodal: either predominantly male or predominantly female. Attendance at places of worship and ritual baths was predominantly reported by males. Overall, the community was balanced to within a few percent. **C**. The distribution of average ages in the various settings (for the community in general, this is simply the age distribution of individuals). Note that attendance at primary school is disjoint with attendance at secondary schools, and attendance at schools was mostly disjoint with places of worship or ritual baths.

We used the information reported in the household survey to generate a bipartite network of people and places (illustrative example in Figure 1A. In this network there is an edge between every individual and each place with which they reported an association. We found that the greatest mean degree was in primary schools, and the least within households (Table 1).

**Table 1:** Characteristics of the network in edges and degree for each setting. For general community transmission and transmission to adult women, the figure in the second column is the number of individuals. Final column is transmissibility estimate and 95 % credible interval for each location. *β* has units of rate of transmission per embedding (see below) unit time.

| Setting | Total edges / individuals | Mean degree | Median degree | 95th percentile degree | Max degree | Transmission rate: $\beta$ [95% CI] |
| --- | --- | --- | --- | --- | --- | --- |
| Household | 1942 | 5.2 | 5 | 10 | 14 | 0.16 [0.13-0.19] |
| Primary school | 686 | 22.9 | 16.5 | 73.3 | 103 | 0.19 [0.15-0.22] |
| Secondary school | 155 | 7.8 | 6.5 | 21.0 | 22 | 0.26 [0.21-0.30] |
| Place of worship | 768 | 11.1 | 5 | 37.6 | 84 | 0.21 [0.16-0.25] |
| Ritual bath | 392 | 11.2 | 4 | 54 | 73 | 0.23 [0.19-0.27] |
| Community | 1942 | N/A | N/A | N/A | N/A | 0.034 [0.028-0.040] |
| Adult Female | 537 | N/A | N/A | N/A | N/A | 0.078 [0.048-0.114] |

### 2.2 Transmission and relative risk

Using a transmission model with varying susceptibility to infection by age, we fitted the rate parameters for each setting to the empirical distributions of household infection from the serosurvey using the sequential Monte-Carlo method for approximate Bayesian computation(33) (Figure 2C, Table 1). As well as place-mediated transmission processes, we included several transmission processes directly between individuals. These transmission processes distinguish susceptibility by age group to account for reduced susceptibility of children (34) and account for a difference in general community transmission to women because we lack data about places frequented by adult women.

**Figure 2:**
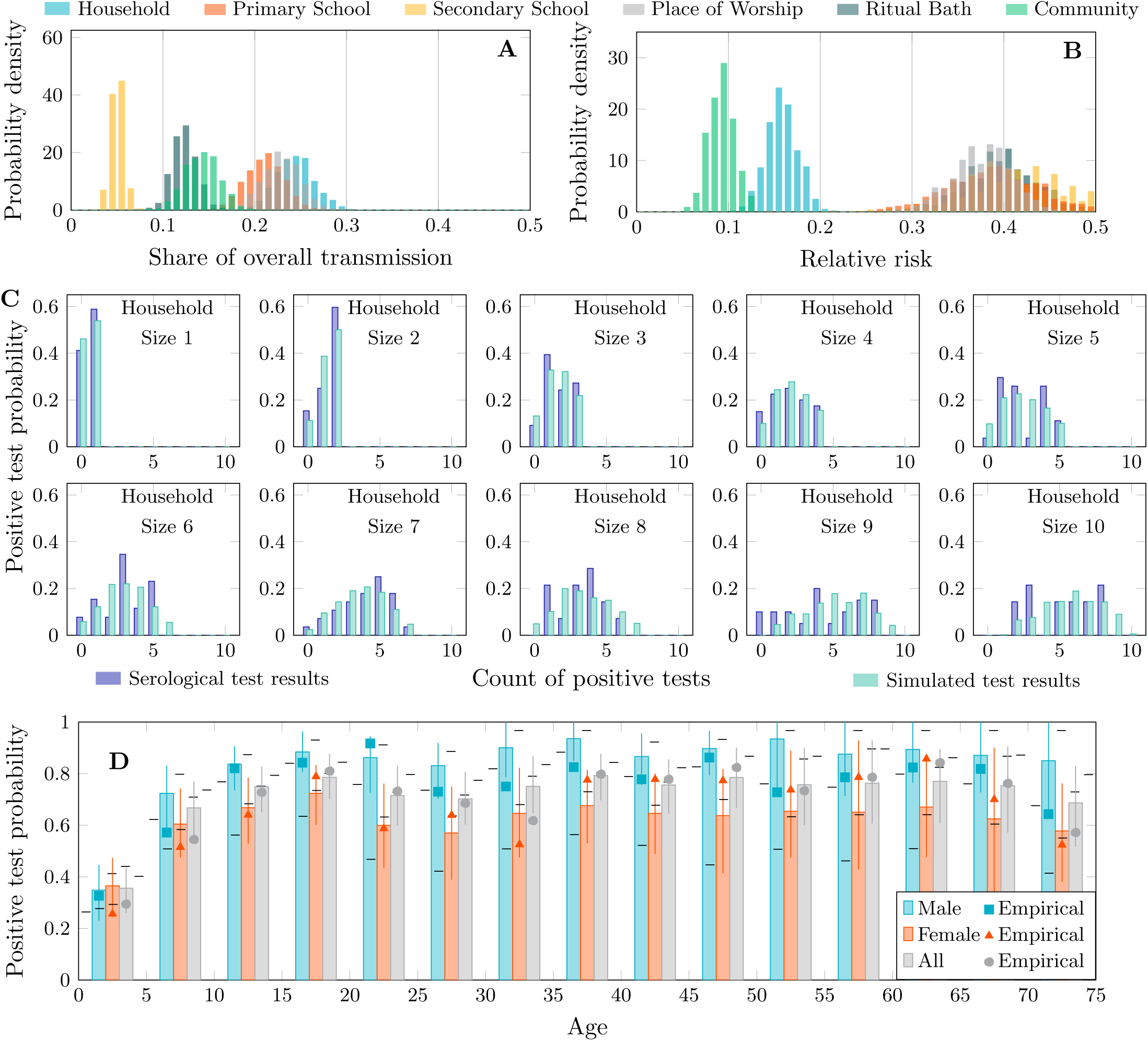
Transmission activity and positive test distributions. **A**. Share of transmission attributable to different settings or location types according to the simulation. **B**. Relative risk of transmission in different settings. This is the amount of transmission that occurred in a given setting relative to the total amount of transmission that is possible in that setting. There is a clear separation between the general community, households, and all other kinds of place. **C**. Probability distributions of positive test results for households of size 1-10 after censoring. The observed distributions are in dark blue and simulations in light blue. **D**. Probability of positive test result by age and sex after censoring. Square, triangle and circle marks indicate the values measured by serosurvey, error bars belong to the simulated values. Note that the model is not explicitly fitted to these data.

Using the place-mediated transmission rate parameters, the model also freely (i.e. without fitting) reproduced the overall seroprevalence of 65% measured in the population as well as the age-specific sero-prevalence (Figure 2 D) though exhibits a male-female asymmetry that was not present in the empirical data, clearly visible in Figure S1. This asymmetry is due to a lack of explicit data about which places adult women have a connection with which cannot be captured by a place-mediated transmission process. To mitigate this, we used an additional well-mixed transmission process for adult women described in more detail in Methods. We additionally found that it was not possible to accurately reproduce prevalence in children assuming age-independent susceptibility Figure S1. When children were assumed to be on average 50% less susceptible than adults in line with other studies (34) then prevalence more closely matched the empirical estimates.

We found that the highest share of transmission within the surveyed population was attributable to households at 24% [95% CI: 20-28%] (Figure 2 A), and the lowest to secondary schools at 5.1% [3.4-7.0%]. The share of transmission outside of places, i.e. the background community rate was 14% [11-18%]. This result was stable: and the strongest estimates of transmission rate (e.g. the parameter with the lowest variance in its posterior distribution and the least sensitivity to changes in network topology) are for general community transmission (Figure S2). That households are identified as the largest share of transmission is perhaps not surprising as every individual belongs to a household but not necessarily to any other setting. Transmission in places of worship was comparable to but slightly lower than households at 23% [19-28%]. Finally, primary schools at 20% [17-25%], and ritual baths account for about 12% [9.5-15%] of total transmission.

The relative risk of transmission in each place is the amount of transmission in a place relative to the total amount of transmission possible in that setting (Figure 2 B). Thus, for households the distribution was centered at 16% [12-19%] which can be interpreted as the relative risk of being infected in the household setting. This quantity is related to the household secondary attack rate but without distinguishing between single and multiple infections acquired elsewhere introduced into the household. All other settings had a greater risk with considerable variance (primary schools 38% [28-47%], secondary schools 41% [31-49%], places of worship 38% [31-45%], and ritual baths 40% [32-47%]). The background risk of infection in the community was 9.3% [6.6-12%].

To facilitate comparisons with other studies of COVID-19 transmission in households, from the fitted transmission rate parameters we calculated the susceptible-infectious transmission probability (SITP) (35, 36). This is the probability that a single infectious individual transmits the virus to a single susceptible individual in a given place, when both are in the reference age group (adults) for each household size. These values are shown in Table 2. Similar quantities can be defined for other settings, but with much larger population sizes the per-pair transmission probabilities are much smaller and difficult to compare across settings. Therefore, we also report the average number of secondary cases a single initial infective in the reference age class would generate in each setting, if all other members were susceptible and in the reference age class. In households, we found this to be 0.52 [0.46-0.56]. For other defined settings we found 0.56 [0.50-0.59] for primary schools, 0.64 [0.58-0.67] for secondary schools, 0.58 [0.52-0.63] for places of worship, and 0.61 [0.56-0.64] for ritual baths.

**Table 2:** Household pairwise susceptible-infectious transmission probability for household sizes 2-10 with 95 % confidence intervals.

| Household size | 2 | 3 | 4 | 5 | 6 | 7 | 8 | 9 | 10 |
| --- | --- | --- | --- | --- | --- | --- | --- | --- | --- |
| $P_{2.5\%}$ | 0.46 | 0.30 | 0.22 | 0.18 | 0.15 | 0.13 | 0.11 | 0.10 | 0.09 |
| Mean | 0.52 | 0.35 | 0.26 | 0.21 | 0.18 | 0.15 | 0.13 | 0.12 | 0.11 |
| $P_{97.5\%}$ | 0.56 | 0.39 | 0.30 | 0.24 | 0.20 | 0.17 | 0.15 | 0.14 | 0.12 |

### 2.3 Multiple household introductions

To determine the most frequent route of introduction of infection into a household we calculated the route of introduction for each household size from our simulations. We found that a minority of transmission events occurred within households, stabilising at 24% [0-62%] for the larger households (Figure 3). Examining the distribution of transmission events by source for each household size in more detail (SI Figure 5) we found that the most common situation was a single introduction from any given source, the exception being primary schools and to a lesser extent places of worship which commonly produced multiple infections for households of size greater than 6. This is consistent with an epidemic with an appreciable amount of transmission pressure from outside households.

**Figure 3:**
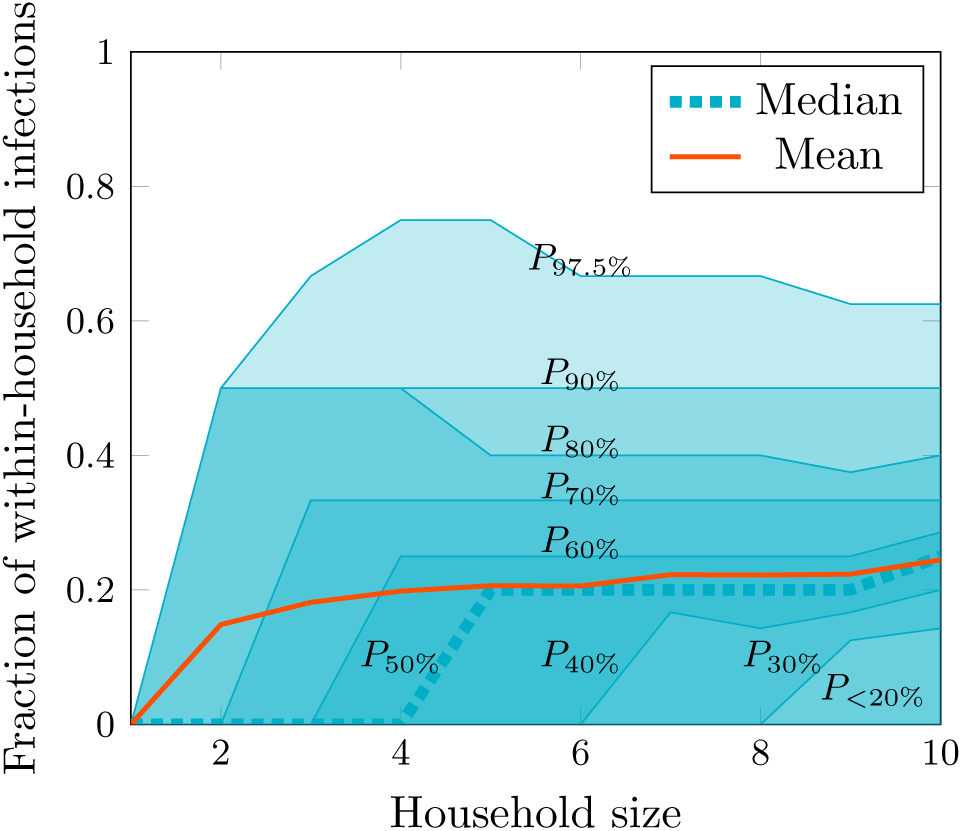
Fraction of within-household transmission events for households of size 1-10. Mean and percentile contours are shown for the fraction of transmission events within households. The maximal contours represent the 95% confidence interval computed as the region contained between *P*_2.5%_ (coincident with the horizontal axis) and *P*_97.5%_.

### 2.4 The role of network structure

Our model is characterised by six transmission mechanisms corresponding to the five types of place and the general community. The degree distributions for some settings, primary schools and places of worship in particular, were very skewed, with a small number of very large places. We hypothesised that large places may have internal structure such as classrooms in schools, or prayer groups during religious observance at places of worship and that it may be inaccurate to represent them as uniformly mixed environments. To determine sensitivity to place size, we split these large places into several smaller places to capture internal structure within those places. The splitting was done such that the total number of edges to each type of place were preserved, and transmission rate parameters were unchanged; only the number of places was modified.

We found that there was an influence of fine structure but this did not substantially affect the distribution of positive tests (Figure 4). If places of worship are constrained to be no larger than the 50th percentile, a reduction of approximately 20% of the peak epidemic size as well as the overall attack rate can be achieved, however sizes above this value have minimal effect. A further reduction by 40-60% can be further achieved if places of worship are made very small, though below the 30th percentile this essentially corresponds to closing them. The effect of smaller school sizes is similar but less pronounced. A 20% reduction in both peak and overall epidemic size can be achieved by closing schools and, short of such a drastic step, smaller school sizes yields approximately a 10% reduction. The percentile differed between places of worship and schools, where the 50th percentile was critical in the former (Supplementary section D, Figure S32-33) and the 10th percentile (Figure S23-24) in the latter. We did find an effect on the relative share of transmission attributable to different settings as well as the relative risk. The uncertainty in these observations is relatively high and at the most optimistic, smaller school sizes could produce as much as a 30% improvement only splitting at the 50th percentile and still be within the 95% confidence intervals. Correspondingly, splitting places of worship at the 50th percentile could yield as much as a 40% improvement in the epidemic trajectory.

**Figure 4:**
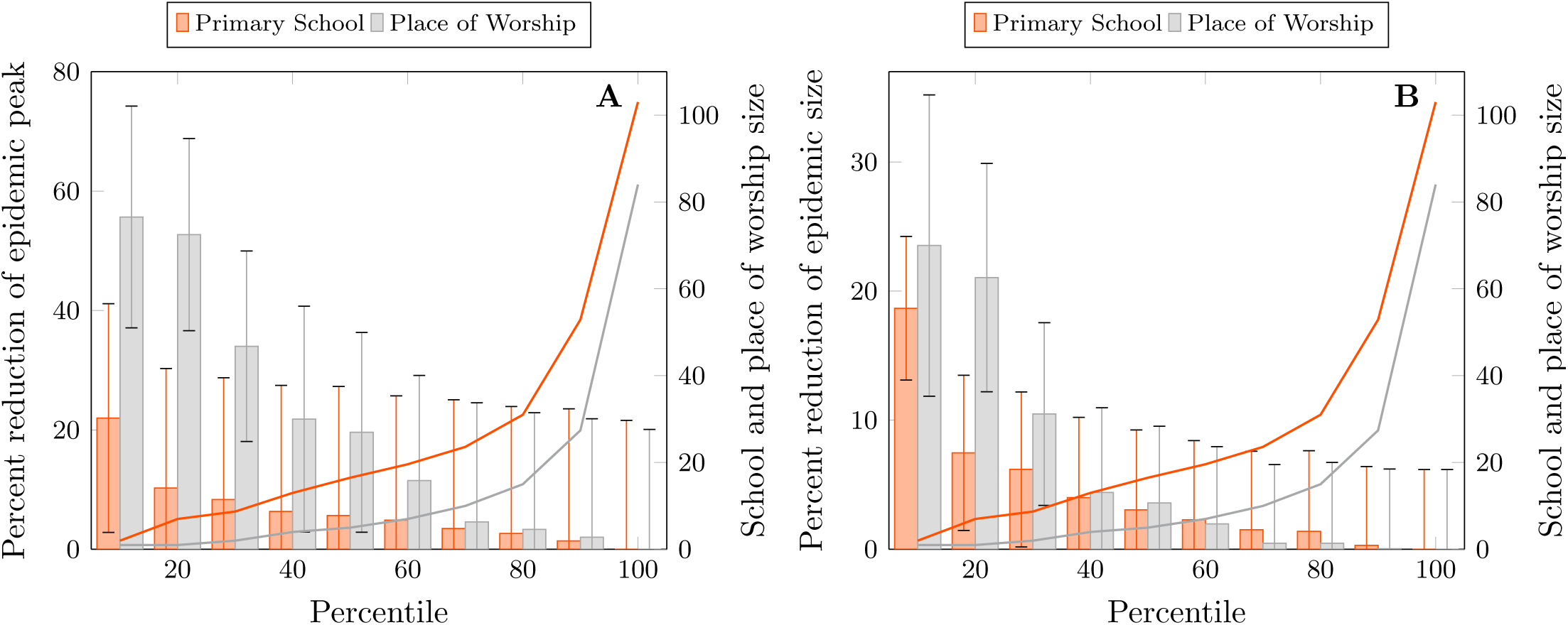
Comparison of epidemic sizes with altered network structure. These figures show estimates of epidemic sizes under conditions where primary schools and places of worship have separately been split such that no institution is larger than the percentile size indicated on the horizontal axis. **A**. shows the peak size of the epidemic and **B**. shows the final size, indicated with bars and the left-hand axis scale. The solid lines and the right-hand axis scale show the percentile institution size.

We also tested the sensitivity of the results to network size, and found that removing up to 20% of households or adding up to 20% has little effect on the household distributions of positive tests or the setting-specific share of overall transmission or relative risk. This suggests that the sampled population was sufficiently large for the present analysis to generalise to the whole population. However, increasing the network size does increase the share of transmission from general well-mixed activity in the community (see Supplementary Section E). It is well-known that a well-mixed dynamic overestimates transmission (32) and the addition of more individuals emphasises the role of this mechanism.

## 3 Discussion

We have estimated the relative contribution of different settings to transmission using explicit people- and place-data in this community. We found that the highest share of transmission within the community was within households, followed by places of worship and primary schools in approximately equal measure. By calculating the relative risk of attending each of these places, we found that primary and secondary schools as well as ritual baths and places of worship had very similar risks. Households have been identified as an important source of transmission since the beginning of the pandemic (37), potentially becoming a higher fraction of transmission during physical-distancing interventions, which could increase the amount of time spent within households.

The relative risk of other settings was higher where adults and children congregate separately. The importance of schools within total community transmission varied between primary and secondary schools. However, only 155 individuals reported an association with secondary schools compared with 686 for primary schools. We found that the relative transmission risk for primary and secondary schools was approximately equal, so the share of total transmission attributable to secondary schools would be comparable if the number of individuals attending were similar. Given the young population, the large proportion of transmission in schools is not surprising and this result may not be generalisable to older populations with a smaller proportion of people in school or similar settings.

We found that by limiting the effective size of gatherings in places of worship to be smaller than the median size, that the peak size of a COVID-19 epidemic in such a tightly knit religious community can be significantly reduced even as the overall size of the epidemic is less affected due to the presence of other transmission pathways. A similar, though less pronounced effect is visible with primary schools; the effect is smaller due to lower susceptibility of children (34). Nearly 200 years ago, Rabbi Eiger wrote of interventions against epidemic cholera:

> *“In my view, it is true that gathering in a small space is inappropriate, but it is possible to pray in groups, each one very small – about 15 people altogether. The groups should begin with first light and then another group, and each one should have a designated time to come and pray there*.*”*

– Letter from Rabbi Akiva Eiger, Posen, 1831

Though this would likely have had little effect for limiting the spread of cholera the overall message is strikingly similar for COVID-19, and likely for similarly-transmitted infections.

These results also emphasise the importance of structure in the contact patterns of the population. In the case of this study, this is evident with the larger schools and places of worship but the principle could reasonably be extended to workplaces. Schools are divided into classes, group lessons take place at places of worship, and worship is conducted in groups that do not necessarily include the entire congregation. It is reasonable to expect some mixing between classes but the majority of time spent in proximity is within a class group. We found that accounting for this effect reduced the overall contribution of these settings which indicates caution in interpreting studies involving transmission in large settings that do not consider heterogeneity within them.

Strictly-Orthodox Judaism consists of diverse groups but can be characterised by stringent interpretations of Jewish law (halachah), which governs almost every aspect of daily life and most relevant for this study, social roles of and relations between men and women (38, 39). Activities unique to men in this population, as compared to the wider UK population, include three-times daily collective prayer, religious study, and daily immersion in ritual baths. Moreover, strictly-Orthodox Jews often maintain some social and spatial separation from the wider metropolitan population that has implications for education and adult employment patterns. Strictly-Orthodox communities have larger youth populations and due to larger household sizes, within metropolitan areas, there is often household overcrowding (40, 41). These factors may potentially affect the generalisability of the transmission rates to the entire population: for example, the contributions of places of worship and ritual baths in this subpopulation are far higher than they would be in a population that does not attend these places. In other populations there may be settings such as certain kinds of workplaces that play a similar role in transmission (42). We did find a substantial contribution of school settings to transmission in this high-prevalence setting, consistent with some findings of the contributions of schools to transmission in moderate prevalence settings (22). The techniques that we developed to conduct this analysis do generalise and can usefully be applied to other populations where appropriate data is available. Cognate data would include households and schools and could sensibly include workplaces for which we do not have explicit information from this survey.

The probability of transmission in each setting, as measured by the SITP, was comparable for households to other studies in the UK (36) based on prevalence, but higher than those calculated for traced contacts (43, 44). The estimates for community transmission are higher than those found for other populations (43), which is not surprising given that strictly-Orthodox communities have practices and requirements that may increase the chance of repeated contacts, compared with the wider population.

We did not have explicit information about all possible places that community members attend, for example workplaces, and some respondents indicated non-specific responses for attendance at places of worship. To address this, we augmented the setting-specific transmission model with general community transmission and this improved the fit to the seroprevalence data. This indicates that transmission outside of those places that are explicitly represented is relevant. The lack of information was particularly marked for adult women about whom we have the least place-based information. In strictly-Orthodox Jewish communities women do not, as a rule, regularly attend places of worship and when attending ritual baths they do so alone (apart, perhaps from a shared waiting area) whereas for men it is a group activity. We know from the serological data that women became infected at approximately the same rate as men. We compensated for this lack of information in the survey data with an additional well-mixed transmission mechanism by which adult women may become infected and added a fitting penalty for sex asymmetry. These mechanisms necessarily mean that a smaller proportion of transmission events were simulated to occur households. Nevertheless, we find that the overall amount of transmission outside of explicitly known places was relatively small, accounting for about 14% of the total. We can therefore conclude that the majority of transmission is captured by the setting-specific part of the model.

At the time of data collection in November and early December 2020, the second wave of the COVID-19 epidemic was beginning. At various times from March 2020 through to the time of the survey, UK government interventions such as closure and reopening of schools and places of worship, and stay at home orders had changed. We have anecdotal evidence from communal leaders of significant effort among residents to adhere to these measures and strong motivation on their part as leaders to find culturally appropriate solutions to reduce local transmission rates. Indeed the project that produced our previous article (27) and the present one was initiated from within the community. However, we have little specific data about timing, both when and for how long interventions were implemented in the community or how widespread adherence was and how it changed over time. The common strategy of estimating this for populations from mobility data, for example, is not available here. We therefore took a time-homogeneous approach to estimate the overall effect of interventions and measures over this time-period to reproduce the cross-sectional data that was available.

We used an extension of the *κ*-calculus (29), in *κ* language for this study, which is a rule-based stochastic process calculus that is best understood as a graph rewriting system (30, 31) for labelled site graphs. It has been extensively used in its original form in molecular biology to study the dynamics of interacting large molecules or polymers, and has been shown to be applicable to population biology and epidemics (45). Here we have applied the method to epidemics on networks in a real-world setting for the first time. We exploited the rule-based formulations to include several distinct processes: infection in different settings, infection of subpopulations within the community, and disease progression. To implement the setting-specific transmission processes we extended the *κ*-calculus from site graphs to generalise to places with an unlimited number of edges connecting them to individuals, and rules parameterized by graph nodes. These generalisations are a small step in the direction envisioned by the theory of stochastic bigraphs (46).

We have constructed a transmission model suited to this kind of network, but there is ample scope for further development for example, a dynamic network that removes edges to capture changing practices over the course of the epidemic, household- or self-isolation, or incorporating within-host immune response models and the interaction of these phenomena with the epidemic at a population level.

We found an important role played by different settings of transmission in a strictly-Orthodox Jewish community in the UK. This study underlines the influence of structure within institutions for understanding transmission, and it follows that altering this structure for example with cohorting strategies, smaller groups and class sizes can be a useful measure to reduce transmission. This study also shows the benefit of analysis of a real network representing connectivity between people and the places that they frequent, in order to allow detailed understanding of transmission dynamics which can inform public health interventions.

## Supporting information

Technical Supplement

## Data Availability

All data produced in the present study are available upon reasonable request to the authors

https://github.com/wwaites/stamford-hill

## Author contributions

WW and RME designed and WW implemented the mathematical model. WW conducted the numerical experiments with assistance from CABP, TH and LP. WW and RME drafted the main text. KG, MM, RME, ChR, SL, NRHS designed the household survey. MJ, VG, AH and DG conducted serological assays in the laboratory. All authors contributed to analysis and interpretation of the survey results and results of the numerical experiments.

## Acknowledgements

The authors would like to thank David Manheim for insightful discussion. This work was jointly funded by UKRI and NIHR [COV0335; MR/V027956/1], a donation from the LSHTM Alumni COVID-19 response fund, HDR UK (MR/S003975/1), the MRC and the Wellcome Trust. The funders had no role in the design, conduct or analysis of the study or the decision to publish.

## Ethics

The study was approved by the London School of Hygiene & Tropical Medicine Ethics Committee (Ref 22532). The study was performed in accordance with all relevant guidelines and regulations. Verbal informed consent was given during the telephone survey and written consent provided prior to phlebotomy. Parents provided written consent for children.

## 4 Materials and Methods

### 4.1 Data

We surveyed 1,942 people from 374 households from a community of approximately 20,000 individuals. Households were chosen in two ways: 346 were randomly sampled from a community-maintained telephone book. The remaining 28 households were chosen from a list with at least one confirmed or suspected COVID-19 case in the household (“enriched households”). The survey methodology is described in detail in our original paper on the descriptive epidemiology of this community (27). We asked each household survey respondent about the membership and composition of their household: which individuals are part of it and their basic demographic details, and about the schools, places of worship, ritual baths (men only) that each household member attended.

We conducted a serological survey for IgG antibodies with affinity to trimeric spike, receptor binding domain and nucleocapsid protein targets from individuals in 309 households of which 24 were in the enriched group, and defined a cut-off value for seropositivity. The seropositivity in this population, 64.3% (95% CI 61.6-67.0%) in total, and 74% (70-77.6%) in adults was greater than estimates for the surrounding metropolitan area at that time, estimated to be 10.8% (9.3-12.5%) by the ONS38. There is some censoring in the serosurvey: not every individual surveyed provided a blood sample, and the distribution of censorship for different household sizes is shown in Figure S2. We excluded serology from the enriched subset of households.

### 4.2 Network generation

We used the information from the household survey to construct a bipartite network as substrate for a transmission model. Vertices or nodes in the network are partitioned into two groups representing people and places. Places furthermore have a label indicating their kind: primary or secondary school, place of worship, ritual bath, or household. The distinction between primary and secondary school is imputed based on age with a culturally-appropriate division at 13 years of age. There is an undirected edge joining a person and a place if that person has reported an association with that place in the survey. If a person is a member of a household, then they have an edge connecting them to that household. If they go to a particular school, they have an edge connecting them to that school. Person nodes are additionally annotated with age and sex as well as lifecycle stage (pre-school, primary school, secondary school and adult).

### 4.3 Transmission model

We used a susceptible-exposed-infectious-removed (SEIR) model for SARS-CoV-2 transmission formulated as a stochastic graph rewriting system (45). Dynamic simulation of this class of rewriting system is done with Gillespie’s algorithm with propensities for a rule given by the number of embeddings of its left-hand side in the underlying graph and its rate constant. This formulation allows us a refined representation of transmission dynamics with two classes of rewriting rule for transmission. The first class is place-mediated transmission where transmission is from a place to an individual connected to that place. It is expressed with rules of the form,

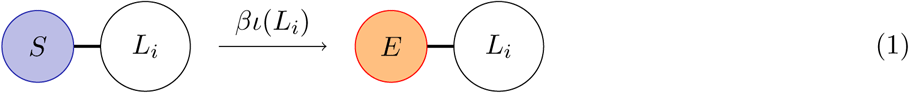

where the left-hand side of the rule is understood as a pattern to match in the underlying graph. The result of matching these patterns is a set of embeddings. The right-hand side is the replacement for a given embedding. The pattern contains an edge between the individual and a specific place *L*_*i*_ for this explicitly place-mediated transmission process. The individual is in the susceptible state, *S*, and becomes exposed indicated with the label *E*. The transmission rate in a place is proportional to the fraction, *ι* (*L*_*i*_) of infectious individuals connected to that place. This follows standard mass-action kinetics assuming that within-place interactions are well-mixed. We fit a rate constant, *β*, particular to each kind of place.

Reporting of places of each type was by characteristic demographic groups and therefore we did not need explicit demographic stratification - differing degrees of contact between demographic groups is implicit in the network topology itself and are a feature of the type of place and not of particular pairs of groups.

The second class of transmission rule occurs directly between individuals,

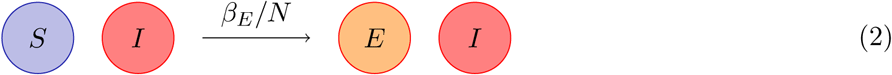

Here, the left-hand side of the rule matches any pair of individuals with the indicated disease progression states regardless of the network connectivity. There are five such well-mixed rules (Equations S13 and S14) to represent transmission at random within the community. The first four represent infection by lifecycle stage. This reflects lower susceptibility of children which we found was necessary to reproduce the probabilities of positive test results by age (Figure 2 D) comparable to the empirical data and as has been reported elsewhere (34). We take the mean estimate of 50% lower susceptibility of children32 and interpolate step-wise, assuming that pre-school children are 25%, primary school 50%, secondary school 75% as susceptible to infection as adults. The fifth is a mechanism for additional infection of adult women in order to compensate for lack of explicit information about the places that they frequent.

The complete model is given in the technical supplement both schematically (Section A) and in code (Section F).

### 4.4 Estimating transmission rate parameters

We fit the six transmission rate parameters to the distribution of positive serological tests in the subset of households that are randomly chosen. These would correspond to household attack rate distributions were there no censoring. In the presence of censoring, these distributions underestimate the attack rate because there is a non-zero probability that individuals who have been infected have not participated in the serological survey. We compare the distribution of simulated censored positive cases to the data using the Wasserstein metric (47, 48) which is designed to provide a well-defined distance between probability distributions. The measure for evaluating goodness of fit is the sum of the distances between distributions for each household size. To calibrate the well-mixed process introduced to compensate for the lack of information about the places frequented by adult women, we augment the fitting measure with a penalty term for asymmetric rates of infection between men and women in the population.

We inferred the transmission rate parameters using the sequential Monte-Carlo method of approximate Bayesian computation (ABC-SMC) starting with 10% of randomly chosen individuals in the infectious state. For efficiency, we fit in two stages. First, we used a uniform prior distribution on all transmission parameters for 10 generations consisting of a total of 2 *×* 10^5^ samples with an acceptance rate of 512 particles per generation to obtain a coarse estimate. Then, for 6 further generations with a total of 3 *×* 10^5^ samples also with an acceptance rate of 512 particles per generation, we used a prior normally distributed about the mean obtained from the first step, with standard deviation of 10%. The resulting kernel density estimates are provided in Supplementary figures 3 and 4.

### 4.5 Simulation

We present results from 1024 simulated epidemics with parameters randomly drawn from the fitted posterior distribution. As with fitting, 10% of individuals are randomly selected to be infectious at the start, and epidemics run for 90 days.

### 4.6 Sensitivity to network structure within places

To determine the effect of place-based heterogeneity in transmission risk, we split locations into parts. Given a percentile, each place with a degree greater than that percentile degree is split into the minimum number of approximately equal sized places that have degree smaller than the percentile degree. For example, if the 95th percentile of primary schools is used as the threshold, the largest primary school with 103 edges would become two primary schools with 51 and 52 edges each. We did this, holding all else the same, in 10 percentile increments from the 90th to the 10th percentile separately for both primary schools and places of worship.

### 4.7 Sensitivity to population size

The data, understood as a bipartite network, are asymmetric in the following sense. One partition, containing places, is complete meaning that all schools, places of worship Ȇc are represented in the data. The partition containing individuals, however, is incomplete; only about 10% of the population was sampled. Furthermore, the total sizes of most places is not known. This asymmetry means that special attention is required to the sensitivity of the results from a place-mediated transmission model on absolute population size. We address this by varying the size of the population. To decrease the population, we select households uniformly at random without replacement to remove, and remove those individuals who are members of the selected households. To increase the population, we select households uniformly at random with replacement and, for each, create a duplicate household whose members are connected to the same places as the role-model. This sensitivity analysis is then to simulate epidemics on these smaller or larger networks and check that the results hold.

### 4.8 Code and data availability

The simulator for the extended version of the *κ*-calculus that we use here is implemented as part of the NetABC package (https://git.sr.ht/~wwaites/netabc). The model and supporting functions for postprocessing the simulation data is available at https://github.com/wwaites/stamford-hill.

