## Supplementary material for "Transmission dynamics of SARS-CoV-2 in a strictly-Orthodox Jewish community in the UK": Technical Supplement

#### A Stochastic Rewriting of Labelled Bipartite Graphs

Our goal is to ascertain the relative contributions to disease transmission of different types of places frequented by individuals within a closed community. The following describes a substrate or state space describing the state of the system, a model that explains the possible transitions from one state to the next, an algorithm for conducting a stochastic simulation of this system, and a method for fitting to data to estimate the parameters governing transition rates. The relative contributions of different places are then estimated by comparing the activity of the infection transition rules corresponding to each place.

Consider a population of size $N$ in an environment that consists of several types, or kinds of place, $T = \{t_1, t_2, \ldots, t_n\}$, where types of place are understood to corresponds to households, schools, workplaces, community gathering places of one kind or another, and so forth. Suppose that we have complete knowledge of the set of places $P$ in this environment, and a morphism, $\tau : P \to T$, that assigns a type to each place. Suppose further that we only have knowledge a proper subset of $P$ that has been explicitly sampled and that we know which sampled individuals spend time in which places. The sampling activity is a morphism $\sigma : P \to V$ that produces the subset of sampled individuals, $V$. Let $L$ be a set of labels consisting of a finite alphabet and that there exists a morphism, $\lambda : V \to L^d$ that associates a tuple of labels to each individual. The $\lambda$ allows us to access demographic information such as age and sex collected during sampling as well as disease progression information that is used and producced in simulation. Finally, we can form a set of edges, $V \xleftarrow{p} E \xrightarrow{q} P$, where the morphisms $p$ and $q$ respectively describe which person and which place correspond to each edge. The structure $G = (V, P, E, T, \tau, L, \lambda, p, q)$ is a labelled bipartite graph. This structure is nothing more than a network of people and places where places have a type (or kind) and people can have various attributes, and is the substrate upon which we construct a transmission model.

We describe transmission dynamics in the form of rewriting rules. Two complementary approaches to are, *Rate equations for graphs* (1) and *Stochastic mechanics of graph rewriting* (2).

We will write the action of rule $a$ as $\mathcal{L}_a \to \mathcal{R}_a$ and the entire rule including its rate as $\mathcal{L}_a \xrightarrow{k_a} \mathcal{R}_a$. The left-hand side $\mathcal{L}_a$ is understood as a pattern, and the right-hand side $\mathcal{R}_a$ is a replacement. The action of a rule is to remove the an embedding of the pattern graph in the concrete graph and gluing in the corresponding replacement.

Particular to our formulation of rules here is the identification of vertices to compute embeddings. This can be understood in two ways. The first is to say that vertices can be identified if all of the labels in the pattern (or replacement) graphs match in the concrete graph. In particular, there may be labels in the concrete graph that do not appear in the pattern. Any labels that do not appear in the rule are preserved. Equivalently, we can consider an enriched graph in which labels are also vertices and an edge exists between a label vertex and a person vertex if that person has such a label. Rewriting is then done on an unlabelled graph. Because the matching operation is subgraph isomorphism, which is expensive in terms of the pattern graph size, we do not implement rewriting systems this way, nevertheless, the two representations of labelled graph or unlabelled property graph are isomorphic. This enables us to write rules that modify, for example, the disease progression state of an individual without explicitly considering their demographic characteristics. It is *possible* to write rules that consider such characteristics, but it is not necessary.

We wish to simulate dynamics where rules are applied with different rates. We consider rules to be applied according to a mass-action principle where the quantity corresponding to mass is the number of embeddings of the left-hand side, $\mathcal{L}_a$ of the rule. Alternatively, we formulate the continuous time Markov chain where transitions are the actions of the rules with the corresponding rates $k_a$. We can therefore construct the Hamiltonian for a system of rules as,

$$\mathcal{H} = \sum_a \mathcal{L}_a \xrightarrow{k_a} \mathcal{R}_a \tag{1}$$

This expression is well-formed because rules form an algebra (2). Denoting distributions over labelled bipartite graphs as $\mathcal{D}$, we can describe the dynamics of the system as,

$$\frac{d\psi}{dt} = \mathcal{H} \tag{2}$$

where $\psi : t \to \mathcal{D}$. This of course has the well-known formal solution,

$$\psi = \psi_0 e^{\mathcal{H}t} \tag{3}$$

We use Gillespie's algorithm to sample trajectories of the system.

### B Infectious Disease Transmission Dynamics

Though graph rewriting admits models where edges can be created or destroyed, we do not make use of that facility here. The graph that we have derived from survey data is fixed by the data and we simply rewrite labels. The general dynamic of the model is of type Susceptible-Exposed-Infectious-Removed (SEIR).

Regardless of the substrate, be it well-mixed with mass-action kinetics, individual-based or network based, the standard formulation of transmission models for infectious diseases transmitted between humans is for the infection process to be precisely that, an interaction between individuals. In this case, we do not know the number of individuals associated with any given place, apart from the households where, for those that are in the sample, we have a complete list of members. We therefore make three assumptions: (i) that interactions within a given place are well-mixed, (ii) that the baseline infection probability per unit time is the same for any two places of the same type, and (iii) that our sample is unbiased. The first assumption enables reasoning analogous to compartmental models, that the infection rate for a particular place is proportional to the fraction of infectious individuals associated with that place. The second assumption

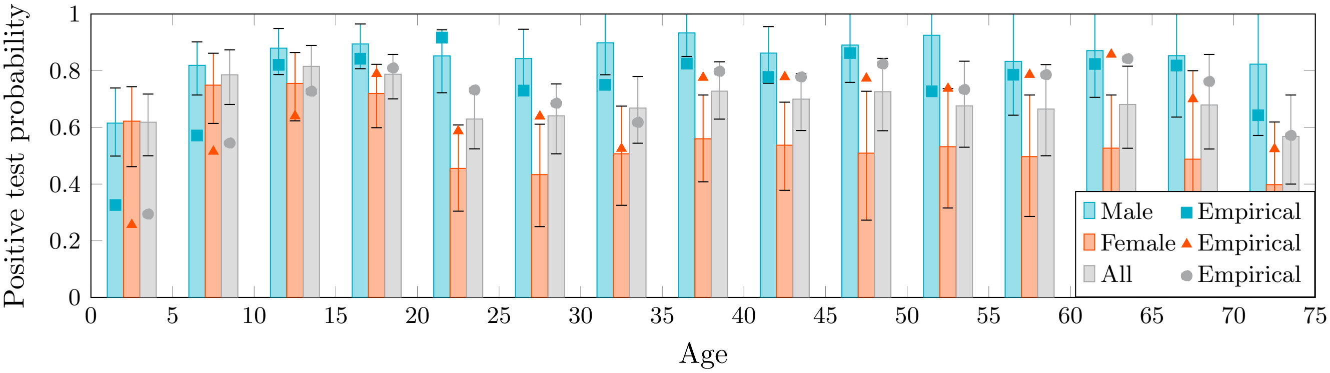


Figure 1: Uncorrected simulated test positivity. This is the result of a simulation where (1) all individuals are equally susceptible, and (2) there is no additional well-mixed infection process for adult women. Note that children become infected more frequently than would be expected (empirical data shown with square, triangle and circle marks) and as well the marked asymmetry between infection of males and females.

is about a kind of uniformity that, while unlikely to represent reality in a strict sense, is a necessary simplification in order to render the model tractible. The third assumption is that computing this fraction from the individuals in our sample gives the same result as computing it using the entire population if we had that information.

Accordingly, we write the location-specific infection process is described by the following rule,

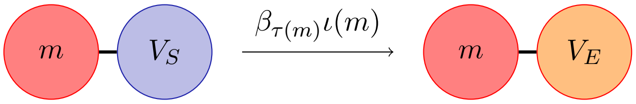


(4)

The upper vertex represents a place, $m$, whose type is given by $\tau(m)$. The lower vertex represents an individual with the label $S$. The individual may make the transition to the exposed state, acquiring the label $E$ at rate $\beta_{\tau(m)}\iota(m)$. This rate consists of a fixed transmission rate $\beta_{\tau(m)}$ characteristic of the type of place, and a scaling factor, $\iota(m)$. The scaling factor is the fraction of neighbours of $m$ that have the label indicating that they are infectious.

It would be unreasonable to believe that every possible infection pathway is accounted for in this model. An individual may be infected in a supermarket or at an event such as a wedding, for example. We therefore add a standard well-mixed transmission mechanism among individuals, unconstrained by the network,

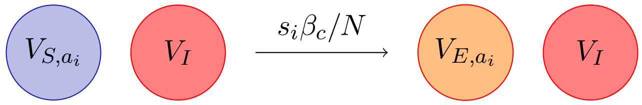


(5)

The semantics of the matching in the left-hand side are that the two individuals may or may not be connected by an edge. This rule captures standard well-mixed epidemic dynamics with a transmission rate $\beta_c$ and total population size $N$. We use separate instances of this rule with the rate scaled by $s_i \in \{0.25, 0.5, 0.75, 1.0\}$ for $a_i$ corresponding to pre-school, primary school, secondary school children and adults. Note that this scaling *only* affects susceptibility and not infectiousness. For reference, simulation with uniform susceptibility produces a result that significantly overestimates the likelihood of children to become infected as can be seen in Figure 1.

While the the data is rich with information about the connections of men and children with the various places, there is very little about women who are mainly connected to their families and rarely to a synagogue. This produces a marked asymmetry visible in Figure 1 in the simulated seropositivity when viewed stratified by sex that is not present in the empirical data. To partly address this limitation in the data, we add a fifth well-mixed rule for adult women with a separate $\beta_f$ that we fit. This does not entirely remove the asymmetry but significantly improves it.

Finally, disease progression is represented in the usual way,

$$V_E \xrightarrow{\alpha} V_I \tag{6}$$

$$V_R \xrightarrow{\gamma} V_R \tag{7}$$

where $\alpha$ and $\gamma$ have the conventional meaning as progression and removal rates.

The complete model is as follows,

$$V_S - H_i \xrightarrow{\beta_H \iota(H_i)} V_E - H_i \tag{8}$$

$$V_S - P_i \xrightarrow{\beta_H \iota(P_i)} V_E - P_i \tag{9}$$

$$V_S - S_i \xrightarrow{\beta_H \iota(S_i)} V_E - S_i \tag{10}$$

$$V_S - G_i \xrightarrow{\beta_H \iota(G_i)} V_E - G_i \tag{11}$$

$$V_S - M_i \xrightarrow{\beta_H \iota(M_i)} V_E - M_i \tag{12}$$

$$V_{S,a_i} \quad V_I \xrightarrow{s_i \beta_C / N} V_{E,a_i} \quad V_I \tag{13}$$

$$V_{S,F} \quad V_I \xrightarrow{\beta_F / N} V_{E,F} \quad V_I \tag{14}$$

$$V_E \xrightarrow{\alpha} V_I \tag{15}$$

$$V_I \xrightarrow{\gamma} V_R \tag{16}$$

Equations 8-12 are the specific forms of Equation 4 for households, primary schools, secondary schools, places of worship and ritual baths, respectively, and each has their characteristic rate parameter. Equation 13 is the set of well-mixed processes as described by Equation 5 in general form for the partition of the population $a_i$ with susceptibility $s_i$. Equation 14 is an additional well-mixed infection process for adult females. The remaining Equations 15 and 16 capture disease progression in the standard way. The rate parameters for progression from exposed to infectious is fixed at 0.2 days$^{-1}$ and infectious to removed at 0.16 days$^{-1}$ corresponding to a mean sojourn time in these states of 5 and 6.6 days respectively.

##### B.1 Susceptible-infectious transmission probability (SITP)

To enable comparison across studies, we report the susceptible-infectious transmission probability (SITP) (**?** ). This quantity is typically studied for households and with suitable assumptions has a closed form ex-

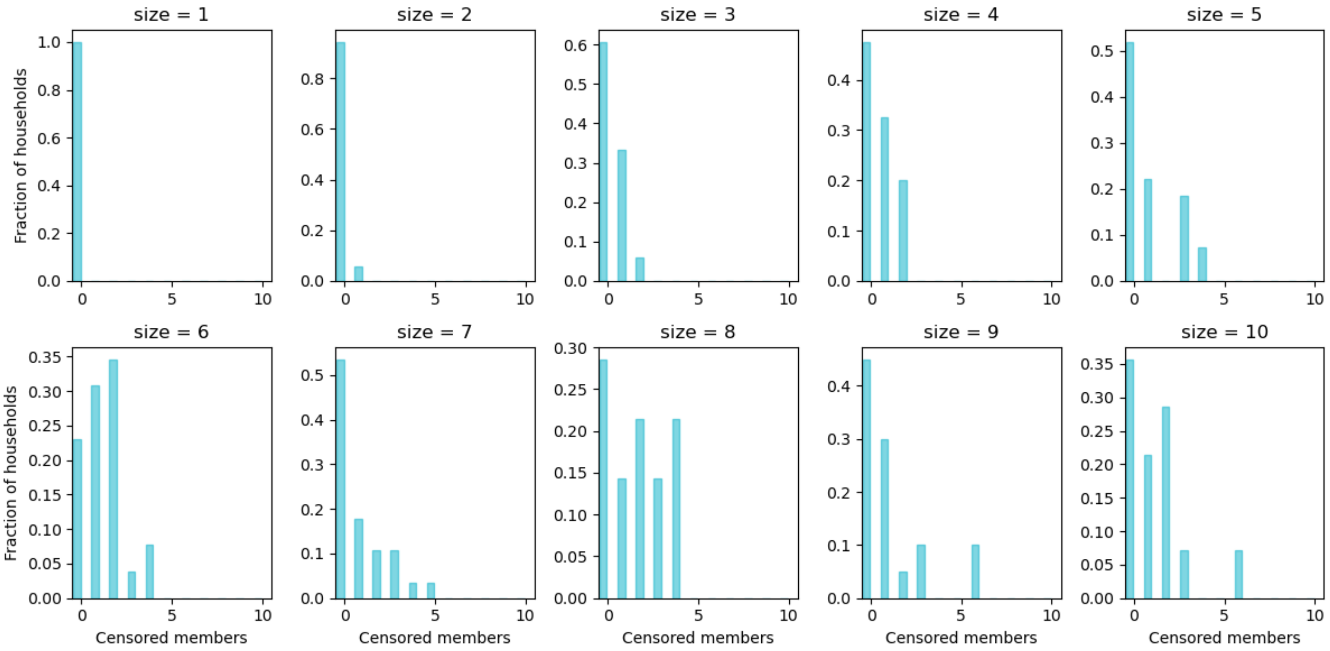


Figure 2: Censorship distributions $\zeta_k$ by household size. For each household with serology, this shows the number of tests actually performed.

pression,

$$\text{SITP} = \frac{\beta}{\beta + \gamma} \tag{17}$$

because we can calculate transmission rate parameters for other settings, we report those as well.

### C Fitting

The rates for transition from exposed to infectious and from infectious to removed we simply take from the literature. We assume that these are intrinsic to the biology of humans and the virus and are not specific to a particular population. The main values of interest are the transmission rates characteristic of the different settings which necessarily must account for varying behaviour in each.

We estimate these values using Approximate Bayesian Computation (ABC). This procedure requires the specification of a measure on model observables. We define the measure as follows. Let $\sigma_k$ be the distribution of positive serological tests in households of size $k$. Let $\zeta_k$ be the censorship distribution in the empirical data for households where at least one individual participated in the serosurvey (Figure 2). This gives the probability that, for households of size $k$ participating in the serosurvey, $i$ individuals participated. For the simulations, let $t_k(g)$ be the distribution of individuals in households of size $k$ that have become infected by the end of the simulation period, e.g. have disease state labels $E$, $I$, or $R$ in the graph $g$. The convolution of $t_k(g) \star \zeta_k$ gives the estimated distribution of positive tests that we would have had in such households with censorship. We compare this with the empirical distribution of positive tests to form to core of our measure.

The 1st Wasserstein metric or Earth Mover's Distance is a metric on the space of probability distributions; it defines a distance between probability distributions via the solution of an optimal transport problem. The intuition is that the difference between two distributions is the minimum work that must be done to rearrange probability mass from one distribution to make it coincide with the other. Writing $W(p, q)$ for the Wasserstein metric, the measure that we will minimise is the sum of the distances between the empirical and simulated distributions,

$$d_W(g) = \sum_k W(\sigma_k, t_k(g) \star \zeta_k) \tag{18}$$

where the sum is taken over all household sizes.

It is possible to define a similar measure using positive test distributions for other settings. The reason that we do not do this is that the data are sparse. For the households in our sample for which we have serology data, we have complete information about their composition (there is likely to be some distortion as we don't always have serology for every household member). We do not have such nearly complete information for the other settings, indeed we do not necessarily even know their absolute size so have no basis upon which to ascertain whether our sample is sufficiently large to accurately characterise the distributions of positive tests in those settings.

The measure $d_W$ is not sufficient to address the asymmetry in the network data for male and female individuals that can be clearly seen in Figure 1. Recall that we have far more information about the places that males frequent than we do for females. This means that the simulation is biased in that males have many more chances to become infected than females do. To correct for this bias, we introduce a penalty term,

$$d_a(g) = \frac{|m(g) - f(g)|}{m(g) + f(g)} \tag{19}$$

where $m(g)$ is the numer of males and $f(g)$ is the number of females who have a disease state label indicating infection in simulation.

The final form of the measure that we minimise for fitting is then,

$$d(g) = d_W(g) + d_a(g) \tag{20}$$

The fitting procedure is done in two stages. In the first stage, a uniform prior is assumed for all parameters. This is an extremely expensive operation and is used to obtain a rough estimate of parameter values. The pairwise kernel density estmates are shown in Figure 3. Once obtained, these estimates are used as informative gaussian priors on the parameter values in a second fitting stage. The mean values from the first stage are preserved, with a standard deviation of 10% of the mean. The pairwise kernel density estimates for the second stage are shown in Figure 4.

#### C.1 Kernel Density Estimates

These figures show the estimates for the posterior distributions of the fitted rate parameters. On the diagonal are the marginal posterior distributions for the parameters individually and below the diagonal are the pairwise joint posterior distributions. The tighter and more focused the distributions, the more constrained the parameter values are. Above the diagonal is the same joint data as an uninterpolated point cloud rather than a heat map.

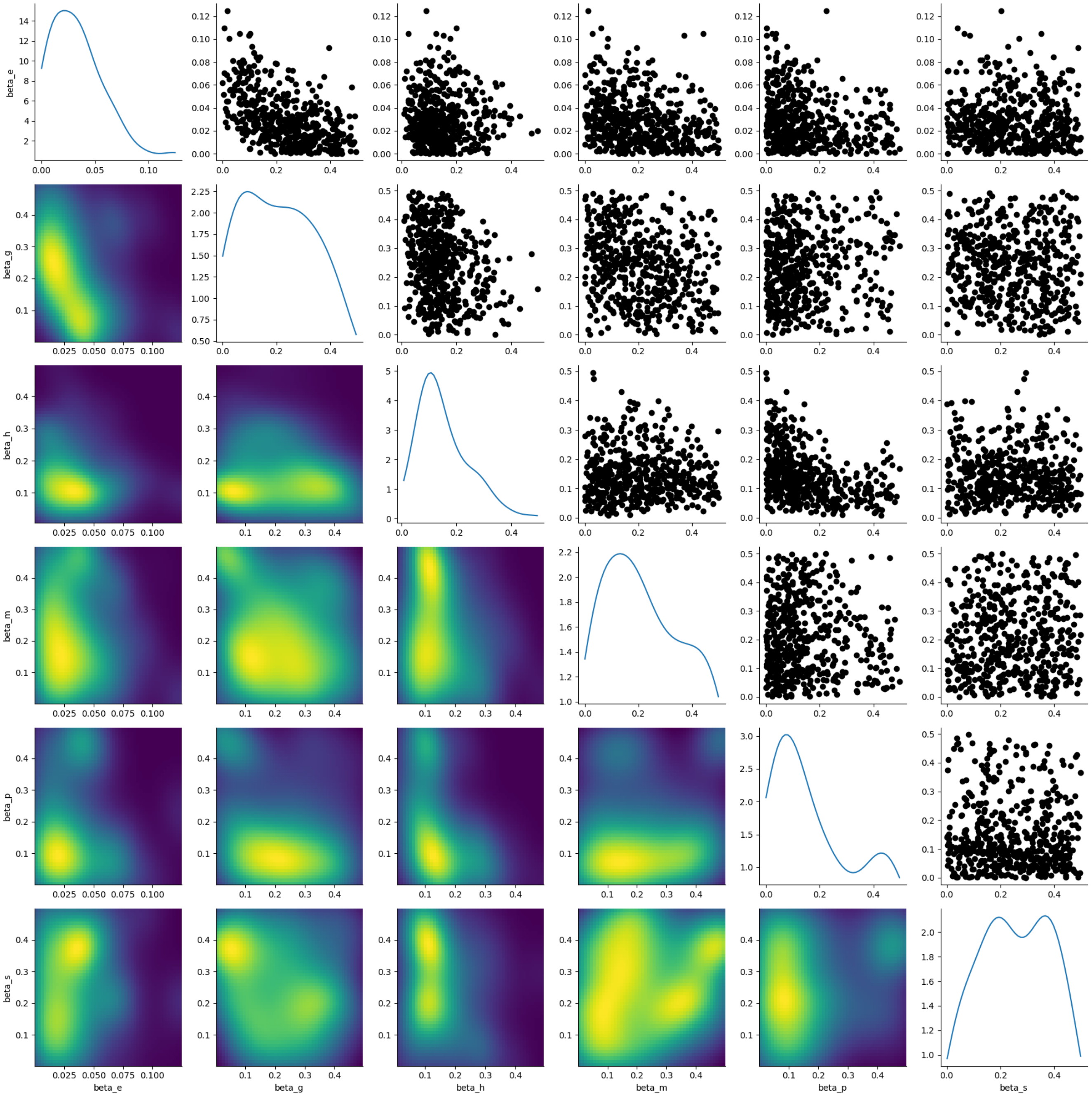


Figure 3: Stage 1 kernel density estimates for fitting from a uniform prior

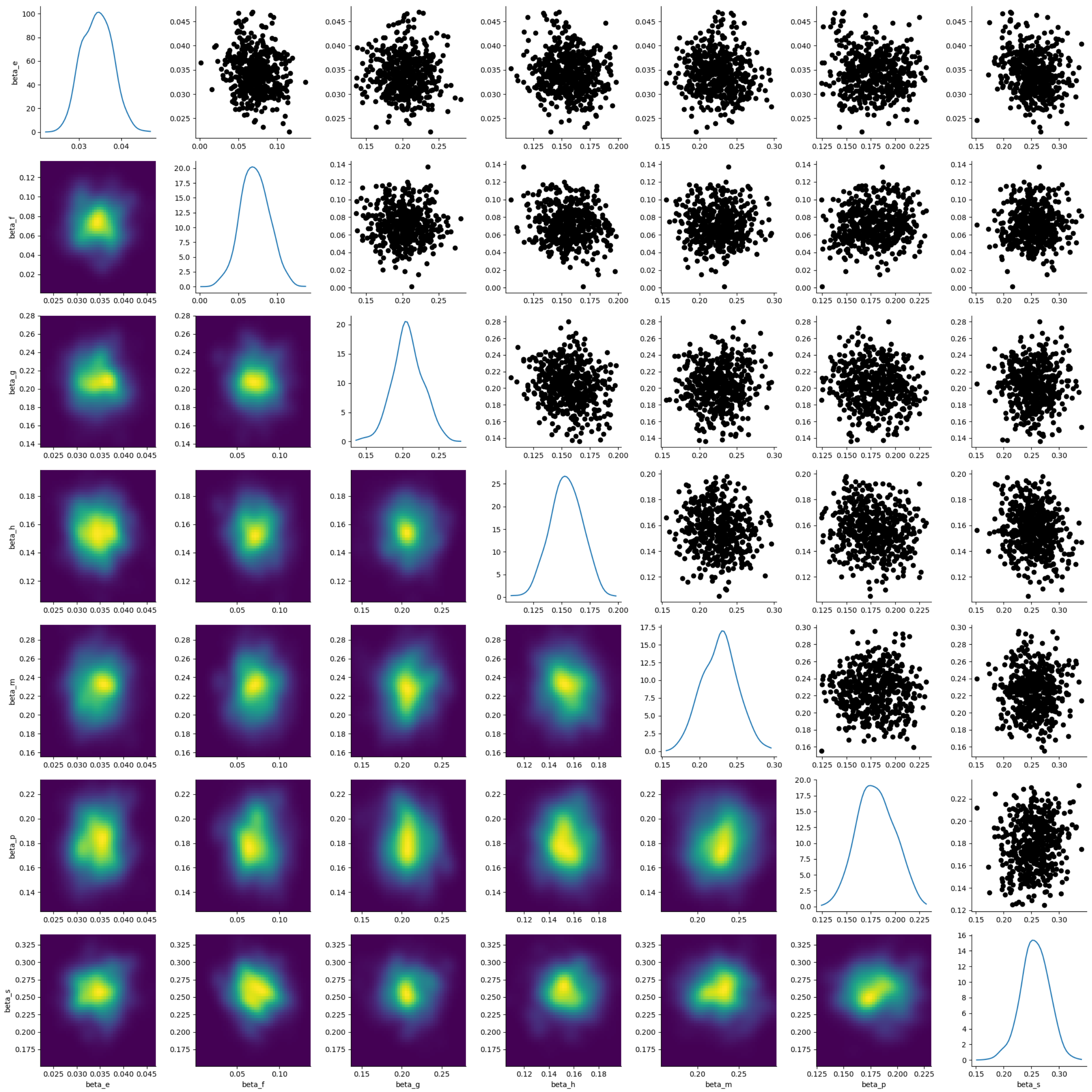


Figure 4: Stage 2 kernel density estimates for fitting from a gaussian prior with means given by Stage 1 fitting and standard deviations set at 10% of the means.

### D Sensitivity to Network Structure

This section shows the result of sensitivity analysis on the structure of the network with a focus on primary schools and places of worship. The procedure for modifying network structure is as follows. Given a percentile, only those places with degree greater than that percentile degree in the network are modified. A place with degree $u$, larger than that percentile degree, $r$, is replaced with the smallest number of approximately equally sized places with degree $v \leq r$. Individuals connected to that place are reallocated to the new places uniformly at random.

The following subsections contain the figures corresponding to Figure 2 in the main text for networks that have been modified by this prescription. Primary schools and places of worship are subject to the splitting procedure in 10 percentile increments from 100% (i.e. the unmodified network) to 50% (all institutions must not be larger than the original median size). Statistics depected in the figures are summarised in Tables 1 and 2.

| **Setting** | **Transmission activity** | | **Relative risk** | |
|---|---|---|---|---|
| Split primary schools at 90th percentile | | | | |
| household | 24.8% | [20.7-29.0%] | 16.6% | [13.3-20.0%] |
| primary school | 21.5% | [17.5-25.5%] | 40.6% | [31.5-48.4%] |
| secondary school | 5.0% | [3.1-6.8%] | 40.9% | [30.3-49.4%] |
| place of worship | 22.4% | [18.4-26.4%] | 37.9% | [31.2-43.4%] |
| ritual bath | 12.1% | [9.3-14.9%] | 39.9% | [32.0-47.3%] |
| community | 14.3% | [10.6-18.1%] | 9.6% | [6.7-12.5%] |
| Split primary schools at 80th percentile | | | | |
| household | 24.9% | [20.8-29.0%] | 16.5% | [13.2-20.0%] |
| primary school | 20.9% | [16.8-24.6%] | 39.1% | [29.6-47.4%] |
| secondary school | 5.0% | [3.3-6.8%] | 40.8% | [30.7-49.0%] |
| place of worship | 22.6% | [18.6-26.8%] | 37.8% | [31.2-43.7%] |
| ritual bath | 12.3% | [9.5-15.3%] | 40.0% | [32.3-47.5%] |
| community | 14.3% | [10.6-18.0%] | 9.5% | [6.6-12.4%] |
| Split primary schools at 70th percentile | | | | |
| household | 25.1% | [20.8-29.4%] | 16.6% | [13.1-20.1%] |
| primary school | 20.4% | [16.5-24.2%] | 38.2% | [29.6-46.4%] |
| secondary school | 5.1% | [3.2-7.0%] | 41.0% | [30.4-49.1%] |
| place of worship | 22.8% | [18.6-27.2%] | 38.1% | [31.1-44.3%] |
| ritual bath | 12.2% | [9.5-15.1%] | 39.9% | [32.1-47.1%] |
| community | 14.4% | [10.7-18.2%] | 9.6% | [6.7-12.4%] |
| Split primary schools at 60th percentile | | | | |
| household | 25.2% | [20.8-29.5%] | 16.5% | [13.1-20.2%] |
| primary school | 19.9% | [15.9-23.9%] | 37.0% | [28.2-45.9%] |
| secondary school | 5.1% | [3.3-7.1%] | 41.0% | [31.1-49.5%] |
| place of worship | 23.1% | [18.9-27.3%] | 38.1% | [31.7-44.2%] |
| ritual bath | 12.3% | [9.5-15.3%] | 39.9% | [32.0-47.2%] |
| community | 14.5% | [10.6-18.3%] | 9.5% | [6.6-12.4%] |
| Split primary schools at 50th percentile | | | | |
| household | 25.3% | [21.3-29.3%] | 16.4% | [13.2-20.0%] |
| primary school | 19.4% | [15.6-23.4%] | 35.7% | [27.5-44.5%] |
| secondary school | 5.1% | [3.5-7.0%] | 41.1% | [30.9-49.2%] |
| place of worship | 23.2% | [18.9-27.4%] | 38.1% | [31.2-44.2%] |
| ritual bath | 12.4% | [9.7-15.4%] | 39.9% | [32.2-47.2%] |
| community | 14.6% | [10.7-18.4%] | 9.5% | [6.6-12.4%] |

| | | | | |
|---|---|---|---|---|
| Split primary schools at 40th percentile | | | | |
| household | 25.4% | [21.4-29.9%] | 16.4% | [12.9-20.1%] |
| primary school | 18.7% | [15.0-22.4%] | 34.2% | [26.1-42.6%] |
| secondary school | 5.2% | [3.5-7.2%] | 41.2% | [31.1-49.4%] |
| place of worship | 23.5% | [19.4-27.6%] | 38.2% | [31.8-44.3%] |
| ritual bath | 12.6% | [9.7-15.4%] | 40.0% | [32.1-47.1%] |
| community | 14.6% | [10.6-18.3%] | 9.4% | [6.5-12.3%] |
| Split primary schools at 30th percentile | | | | |
| household | 25.8% | [21.7-30.0%] | 16.2% | [12.9-19.8%] |
| primary school | 17.0% | [13.5-20.5%] | 30.4% | [23.0-37.8%] |
| secondary school | 5.4% | [3.6-7.3%] | 41.4% | [31.2-49.4%] |
| place of worship | 24.2% | [19.7-28.4%] | 38.4% | [31.4-44.2%] |
| ritual bath | 12.9% | [9.8-16.1%] | 39.9% | [32.0-47.8%] |
| community | 14.8% | [11.0-18.5%] | 9.4% | [6.6-12.3%] |
| Split primary schools at 20th percentile | | | | |
| household | 26.0% | [21.9-30.3%] | 16.1% | [12.7-19.6%] |
| primary school | 15.9% | [12.7-19.3%] | 28.0% | [21.0-35.2%] |
| secondary school | 5.4% | [3.6-7.4%] | 41.3% | [30.3-49.5%] |
| place of worship | 24.6% | [20.2-28.9%] | 38.5% | [31.9-44.4%] |
| ritual bath | 13.1% | [10.3-16.2%] | 40.3% | [32.7-47.3%] |
| community | 15.0% | [10.9-18.8%] | 9.3% | [6.5-12.2%] |
| Split primary schools at 10th percentile | | | | |
| household | 27.5% | [23.1-32.3%] | 15.0% | [11.7-18.7%] |
| primary school | 6.6% | [4.7-8.5%] | 10.2% | [7.5-13.3%] |
| secondary school | 6.3% | [4.2-8.4%] | 41.8% | [31.6-49.5%] |
| place of worship | 28.4% | [23.6-33.2%] | 39.1% | [31.8-45.3%] |
| ritual bath | 15.2% | [11.9-18.5%] | 40.8% | [32.9-47.7%] |
| community | 15.9% | [11.6-20.2%] | 8.7% | [5.7-11.7%] |

Table 1: Sensitivity to network structure: transmission activity and relative risk for splitting of primary schools at different percentile degrees.

| **Setting** | **Transmission activity** | | **Relative risk** | |
|---|---|---|---|---|
| Split places of worship at 90th percentile | | | | |
| household | 25.1% | [21.4-29.6%] | 17.1% | [13.8-20.9%] |
| primary school | 21.8% | [17.9-25.8%] | 41.5% | [32.9-48.8%] |
| secondary school | 5.0% | [3.1-6.6%] | 41.6% | [30.8-49.5%] |
| place of worship | 21.5% | [18.1-25.3%] | 36.7% | [30.6-42.4%] |
| ritual bath | 12.1% | [8.9-15.0%] | 40.3% | [31.6-47.1%] |
| community | 14.6% | [10.8-19.1%] | 9.8% | [7.0-13.3%] |
| Split places of worship at 80th percentile | | | | |
| household | 25.2% | [21.4-29.4%] | 17.0% | [13.7-20.6%] |
| primary school | 21.8% | [17.9-25.8%] | 41.2% | [32.5-48.7%] |
| secondary school | 5.0% | [3.4-7.0%] | 41.4% | [31.4-49.2%] |
| place of worship | 21.0% | [17.1-25.1%] | 35.9% | [29.8-41.2%] |
| ritual bath | 12.2% | [9.6-15.1%] | 40.8% | [32.6-48.6%] |
| community | 14.8% | [11.4-18.3%] | 9.9% | [7.3-13.0%] |
| Split places of worship at 70th percentile | | | | |
| household | 25.7% | [21.6-29.6%] | 17.2% | [13.7-20.4%] |

| | | | | |
|---|---|---|---|---|
| primary school | 21.9% | [17.1-25.5%] | 41.2% | [30.8-48.9%] |
| secondary school | 5.1% | [3.2-7.2%] | 42.1% | [30.6-49.5%] |
| place of worship | 20.2% | [16.6-24.1%] | 34.1% | [28.3-40.5%] |
| ritual bath | 12.7% | [9.6-15.4%] | 41.9% | [33.3-48.5%] |
| community | 14.4% | [10.4-18.2%] | 9.6% | [6.7-12.9%] |
| Split places of worship at 60th percentile | | | | |
| household | 25.8% | [21.4-30.1%] | 17.1% | [13.7-20.9%] |
| primary school | 22.5% | [18.0-26.6%] | 41.8% | [30.6-49.1%] |
| secondary school | 5.4% | [3.6-7.3%] | 42.7% | [35.3-49.5%] |
| place of worship | 19.1% | [16.1-23.1%] | 31.9% | [27.4-37.9%] |
| ritual bath | 12.8% | [10.1-15.7%] | 41.7% | [35.0-48.6%] |
| community | 14.7% | [10.9-18.8%] | 9.7% | [6.8-12.5%] |
| Split places of worship at 50th percentile | | | | |
| household | 26.3% | [22.4-30.3%] | 17.2% | [14.1-20.6%] |
| primary school | 22.4% | [18.4-26.5%] | 41.1% | [32.5-48.8%] |
| secondary school | 5.3% | [3.4-7.4%] | 41.9% | [31.0-49.5%] |
| place of worship | 17.6% | [14.7-20.9%] | 29.0% | [24.1-33.8%] |
| ritual bath | 13.2% | [10.4-16.1%] | 42.5% | [35.5-49.1%] |
| community | 15.1% | [11.2-19.2%] | 9.9% | [7.0-13.1%] |
| Split places of worship at 40th percentile | | | | |
| household | 26.5% | [22.7-31.2%] | 16.9% | [13.5-20.9%] |
| primary school | 22.7% | [18.2-26.4%] | 40.9% | [30.3-48.8%] |
| secondary school | 5.4% | [3.6-7.3%] | 41.9% | [33.6-49.2%] |
| place of worship | 16.5% | [13.7-19.7%] | 26.7% | [22.2-31.3%] |
| ritual bath | 13.5% | [10.6-16.8%] | 42.4% | [34.0-49.1%] |
| community | 15.3% | [10.9-19.5%] | 9.8% | [6.7-13.4%] |
| Split places of worship at 30th percentile | | | | |
| household | 28.4% | [23.8-32.9%] | 17.2% | [12.7-21.4%] |
| primary school | 23.9% | [19.1-28.3%] | 40.3% | [29.8-48.8%] |
| secondary school | 5.9% | [3.7-8.2%] | 42.5% | [31.2-49.5%] |
| place of worship | 10.0% | [7.8-12.4%] | 15.3% | [12.5-18.4%] |
| ritual bath | 15.6% | [12.6-19.4%] | 44.9% | [36.5-49.5%] |
| community | 16.0% | [12.5-20.5%] | 9.7% | [6.5-13.2%] |
| Split places of worship at 20th percentile | | | | |
| household | 30.7% | [26.1-35.2%] | 16.3% | [11.9-20.1%] |
| primary school | 26.1% | [20.8-31.3%] | 38.8% | [26.9-49.2%] |
| secondary school | 6.9% | [4.6-9.6%] | 42.4% | [30.4-49.5%] |
| place of worship | 0.5% | [0.5-1.4%] | 0.5% | [0.5-1.4%] |
| ritual bath | 18.5% | [14.3-22.4%] | 45.7% | [35.7-49.5%] |
| community | 17.8% | [13.7-21.9%] | 9.5% | [6.5-13.1%] |
| Split places of worship at 10th percentile | | | | |
| household | 30.7% | [25.6-35.4%] | 16.6% | [12.6-20.8%] |
| primary school | 26.6% | [20.6-32.1%] | 39.8% | [27.2-48.8%] |
| secondary school | 6.7% | [4.6-9.4%] | 42.6% | [32.5-49.5%] |
| place of worship | 0.5% | [0.5-1.4%] | 0.5% | [0.5-1.4%] |
| ritual bath | 17.7% | [13.9-22.0%] | 44.9% | [36.3-49.5%] |
| community | 18.2% | [14.0-22.2%] | 9.9% | [6.7-12.6%] |

Table 2: Sensitivity to network structure: transmission activity and relative risk for splitting of places of worship at different percentile degrees.

#### D.1 Unmodified network

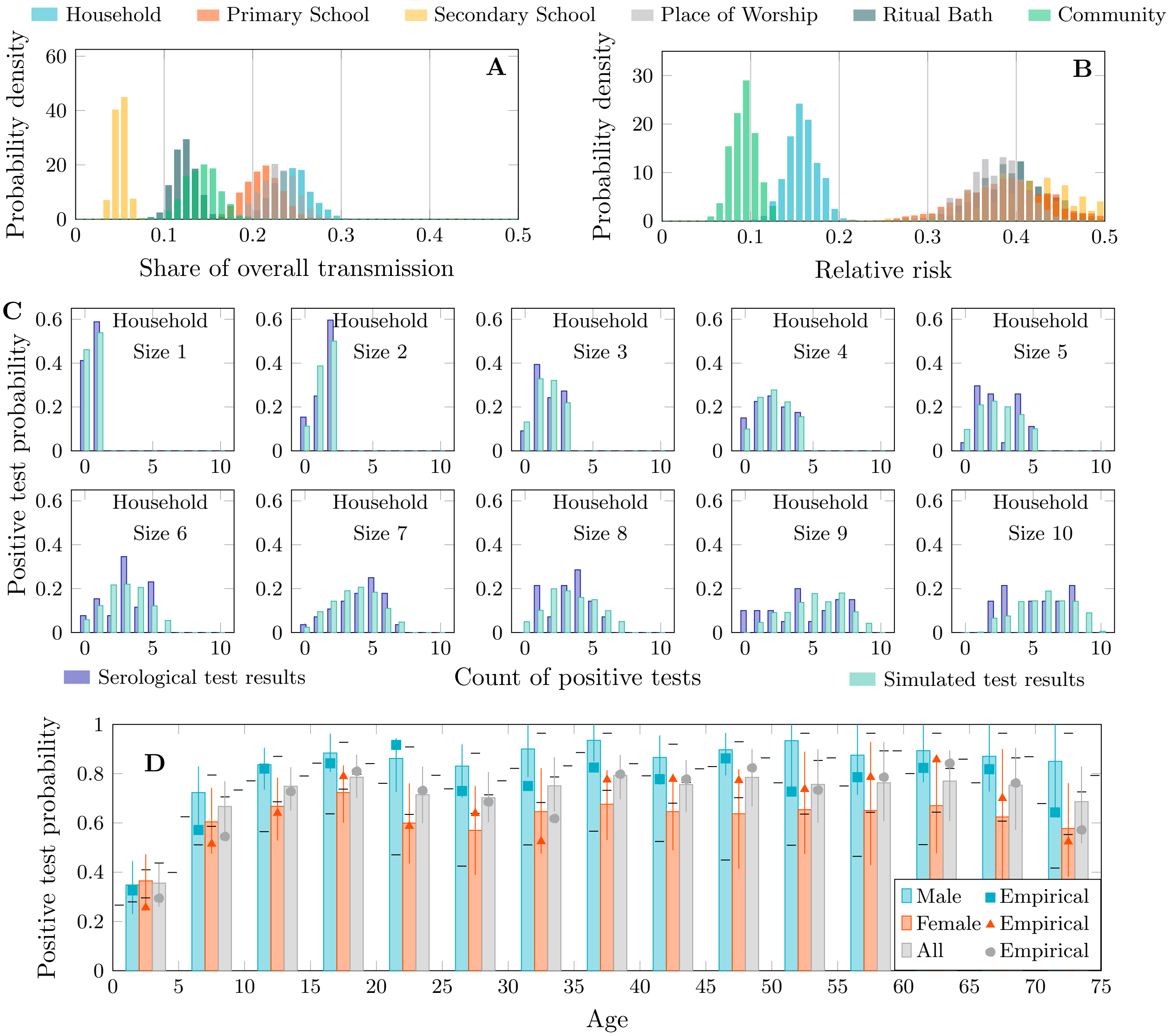


Figure 5: Transmission share, relative risk and postitive test distributions (Reproduction of Figure 2 from main text)

[H]

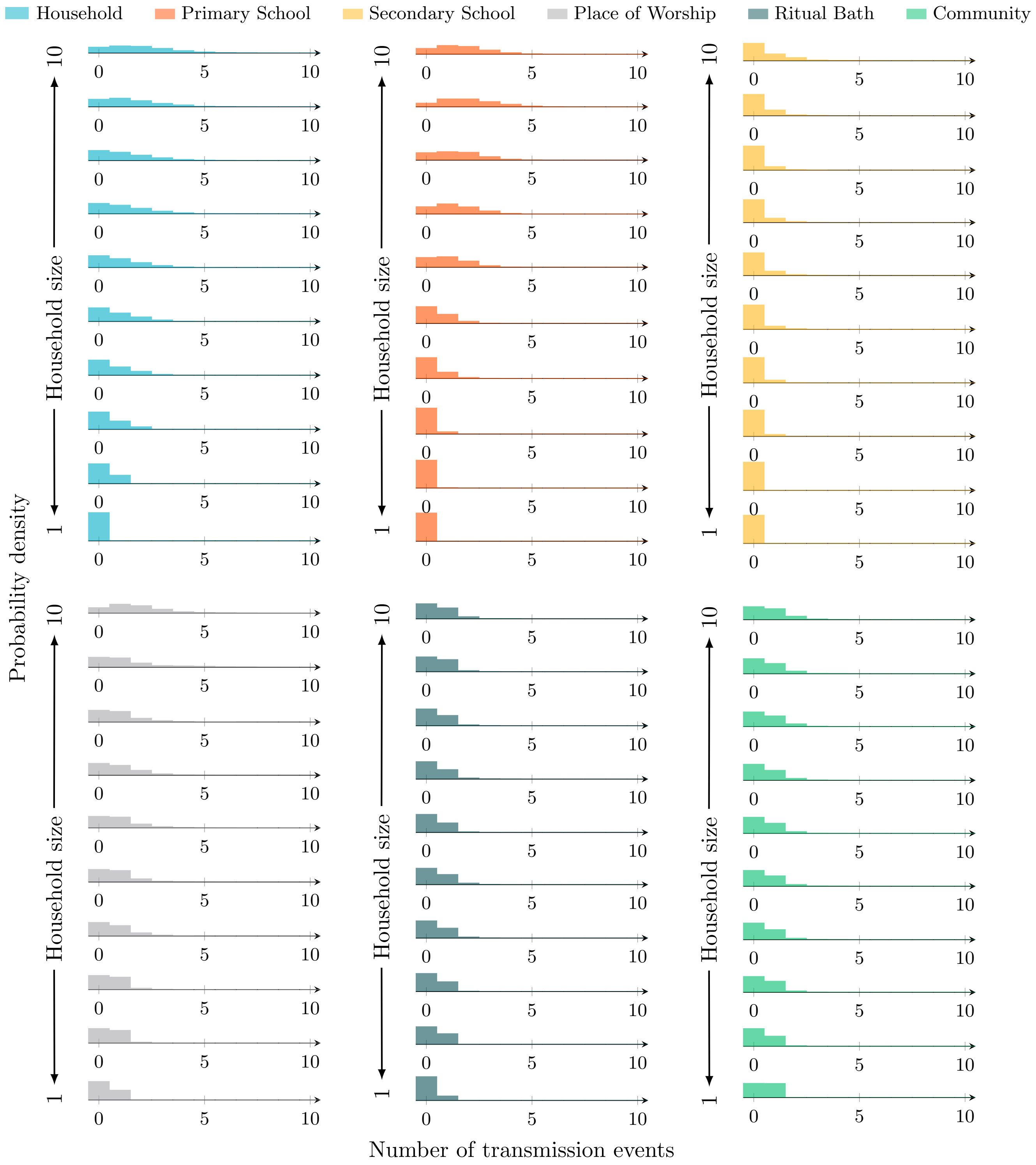


Figure 6: Transmission event distributions by source and household size

#### D.2 Splitting primary schools (90th percentile)

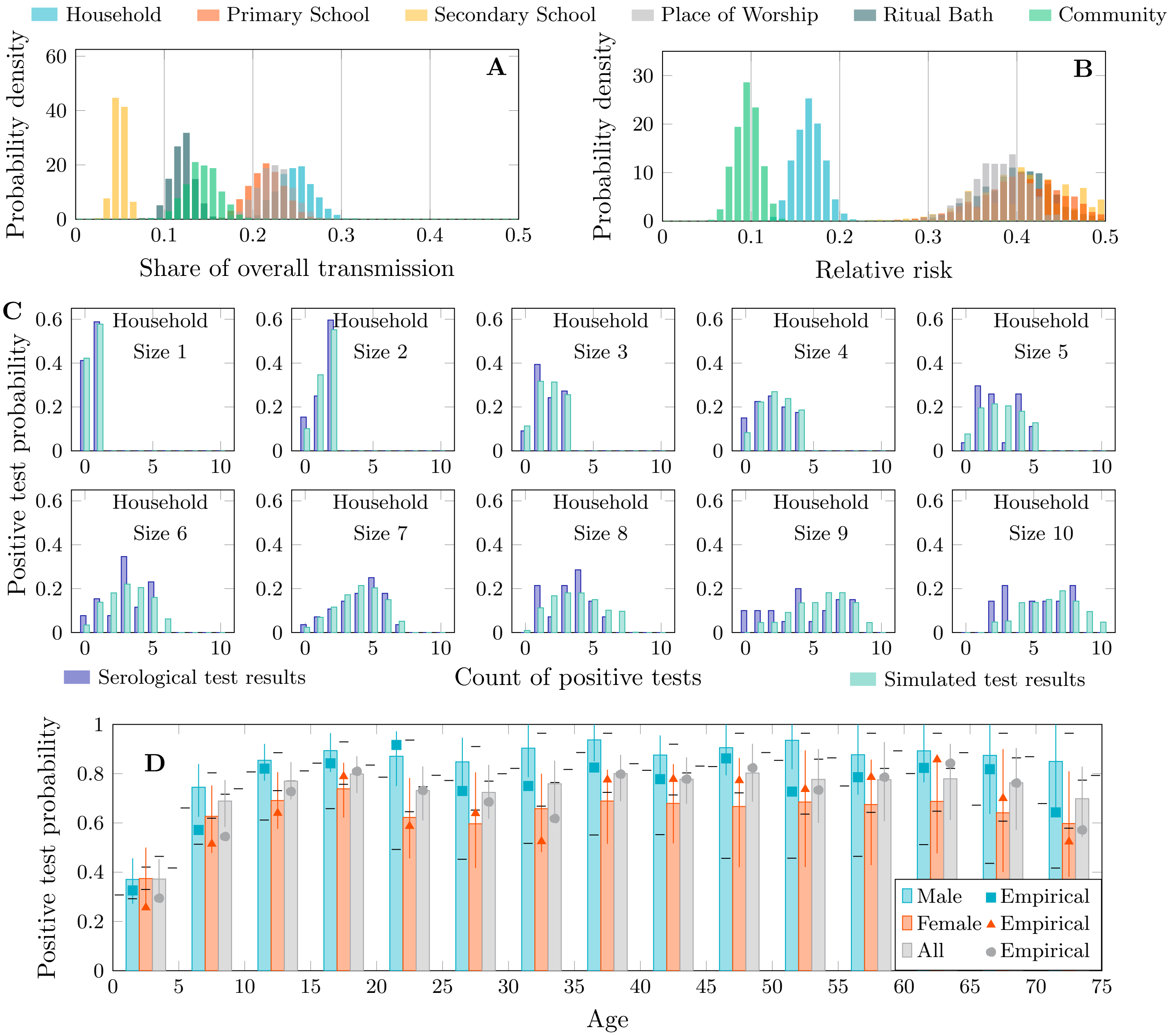


Figure 7: Transmission share, relative risk and postitive test distributions – Splitting primary schools (90th percentile)

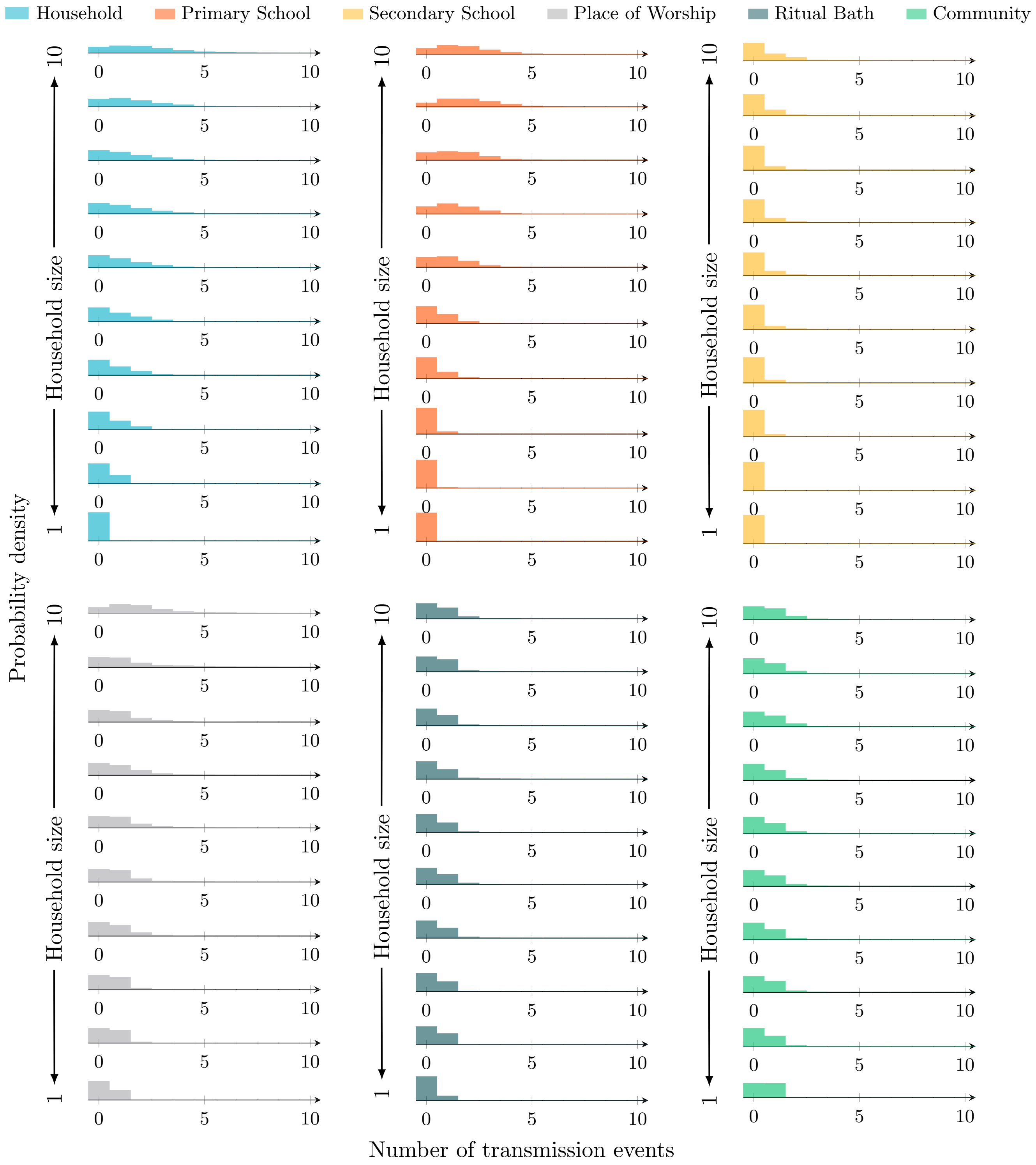


Figure 8: Transmission event distributions by source and household size – Splitting primary schools (90th percentile)

#### D.3 Splitting primary schools (80th percentile)

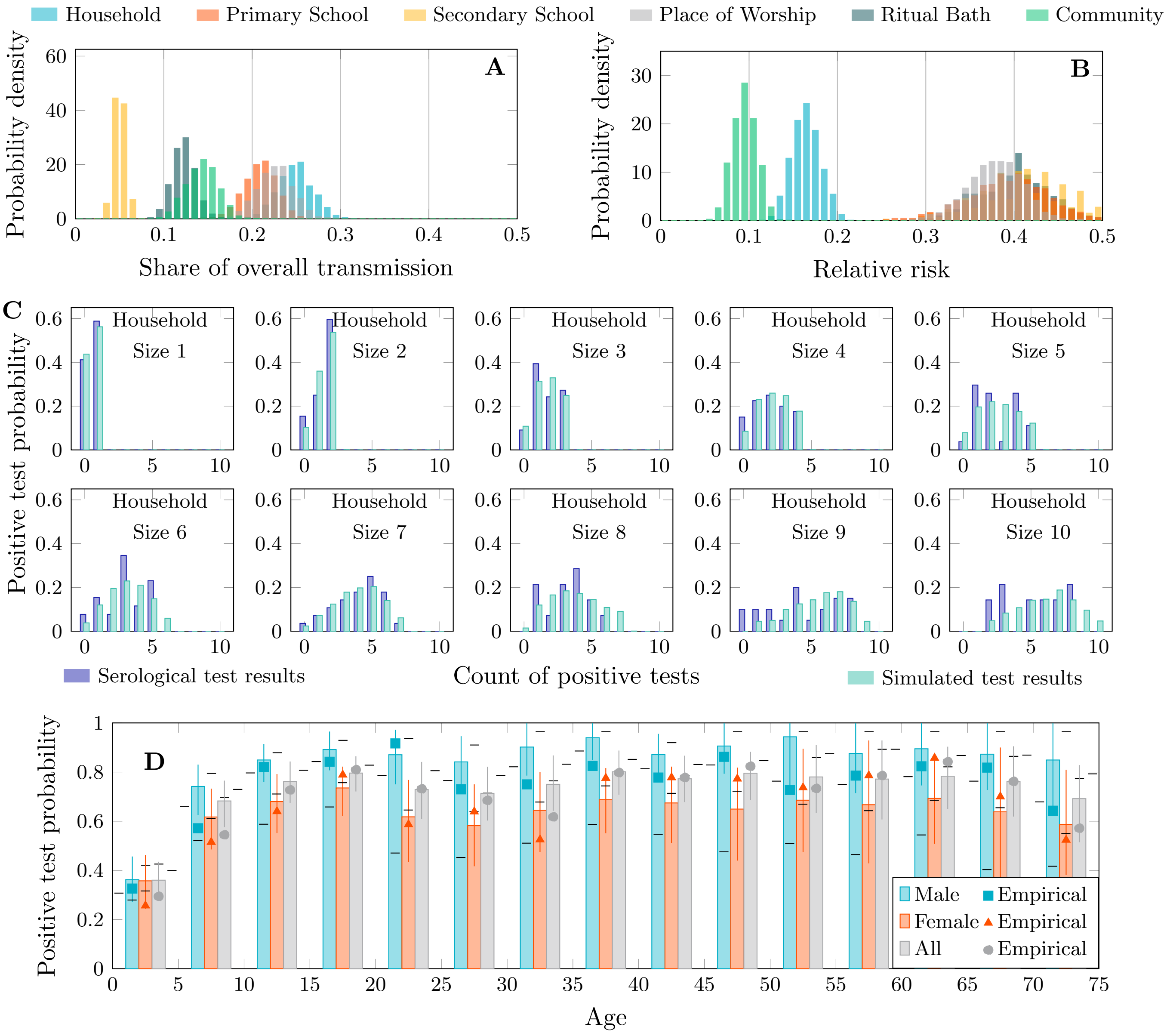


Figure 9: Transmission share, relative risk and postitive test distributions – Splitting primary schools (80th percentile)

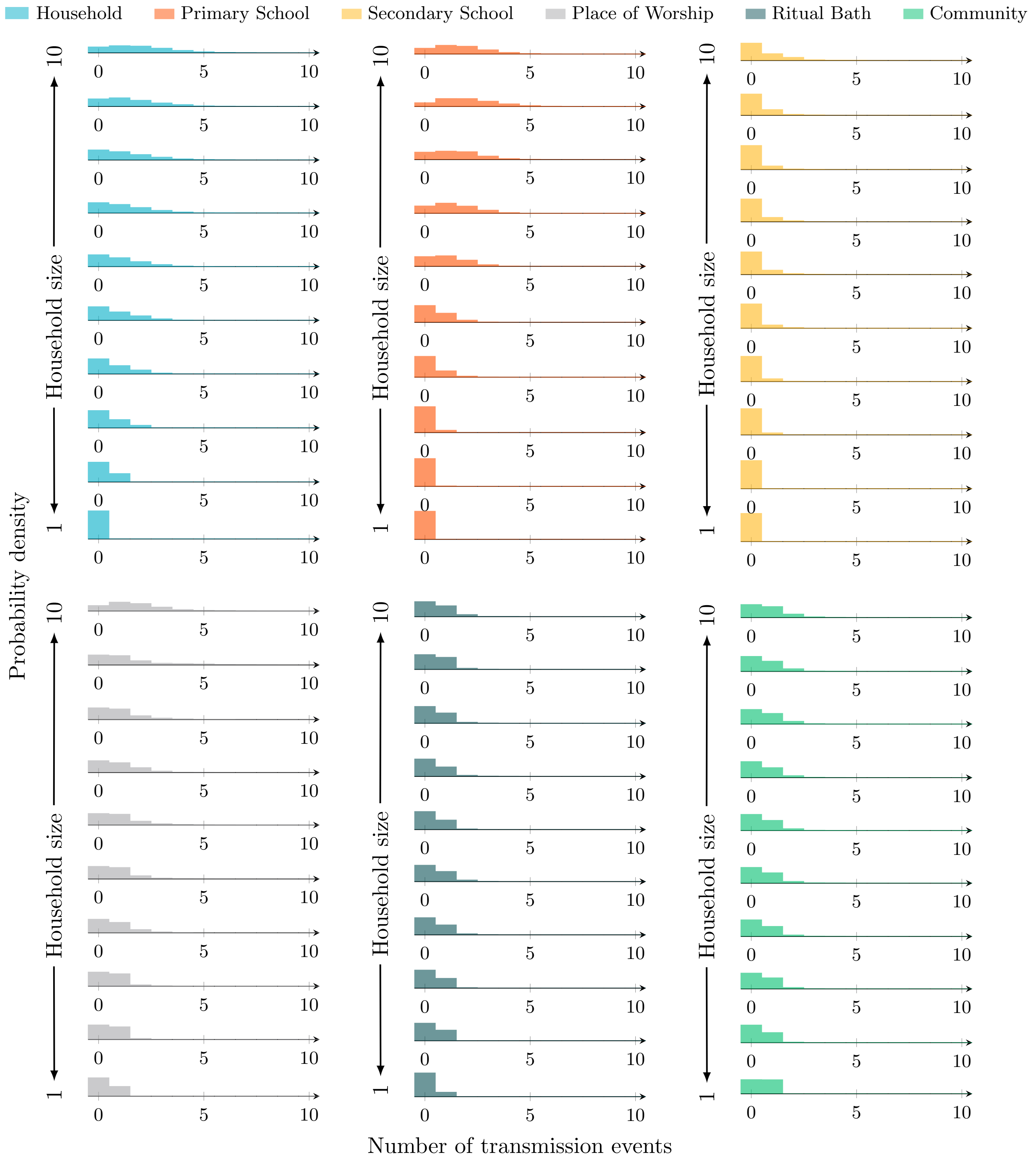


Figure 10: Transmission event distributions by source and household size – Splitting primary schools (80th percentile)

#### D.4 Splitting primary schools (70th percentile)

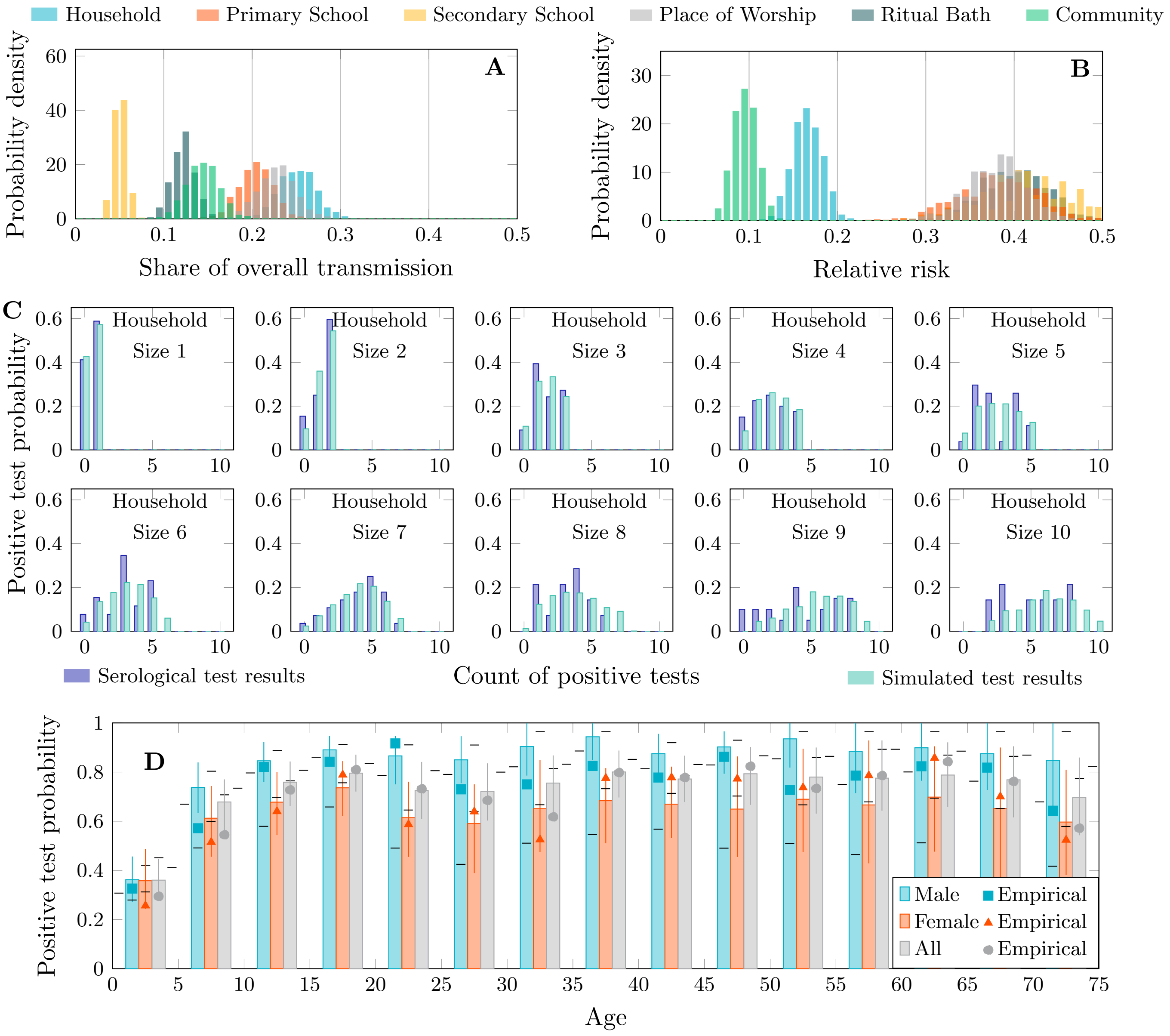


Figure 11: Transmission share, relative risk and postitive test distributions – Splitting primary schools (70th percentile)

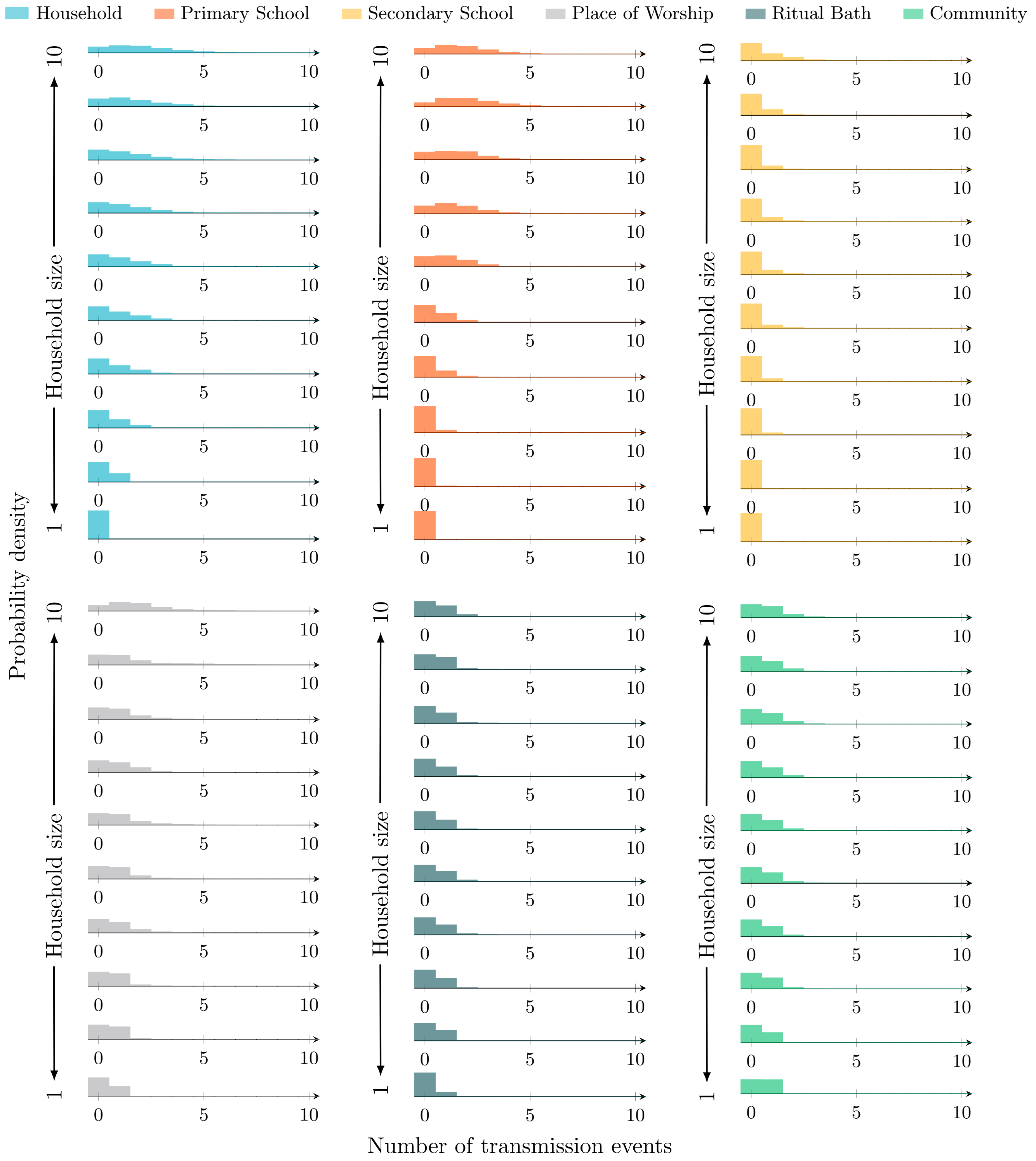


Figure 12: Transmission event distributions by source and household size – Splitting primary schools (70th percentile)

#### D.5 Splitting primary schools (60th percentile)

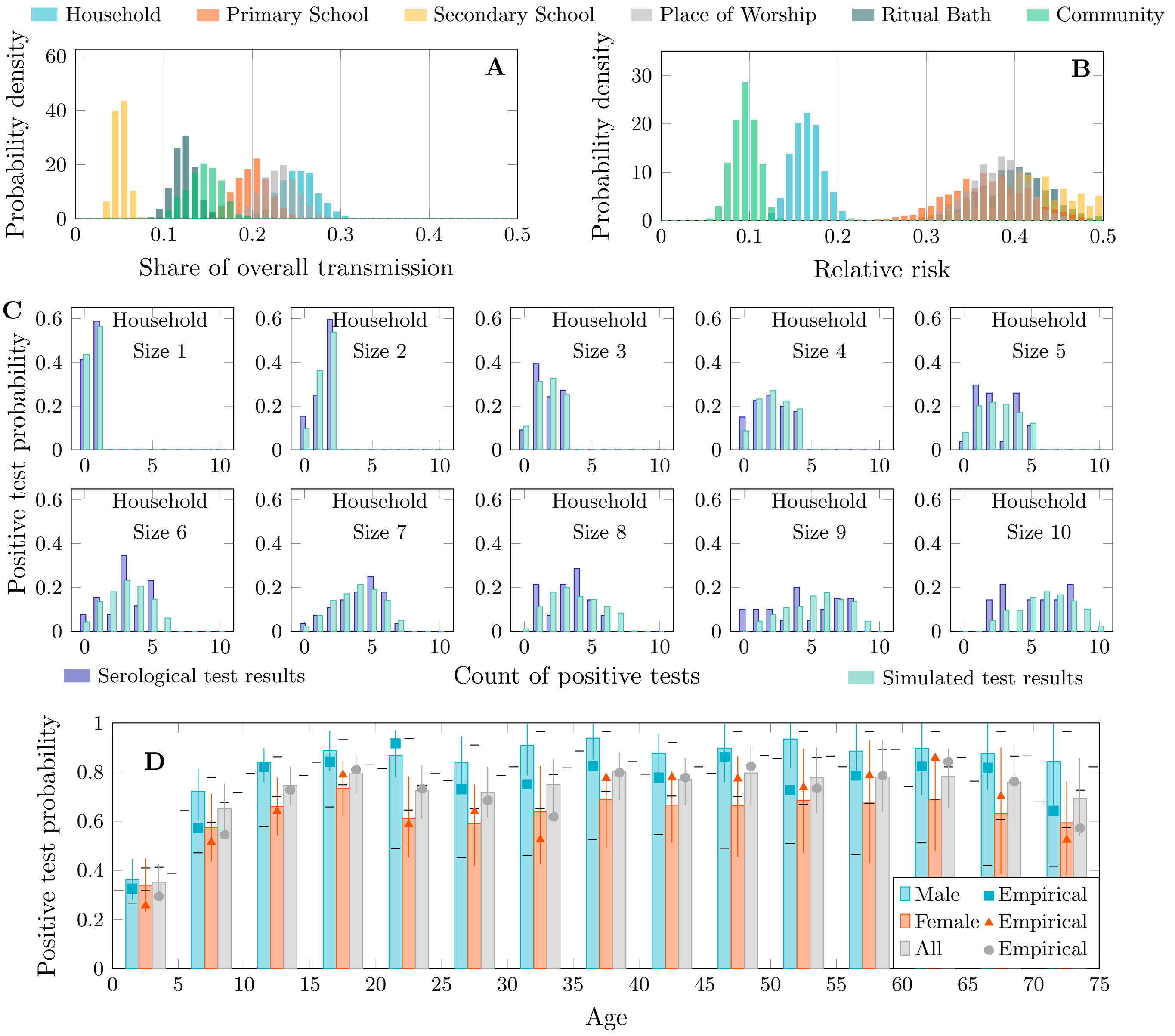


Figure 13: Transmission share, relative risk and postitive test distributions – Splitting primary schools (60th percentile)

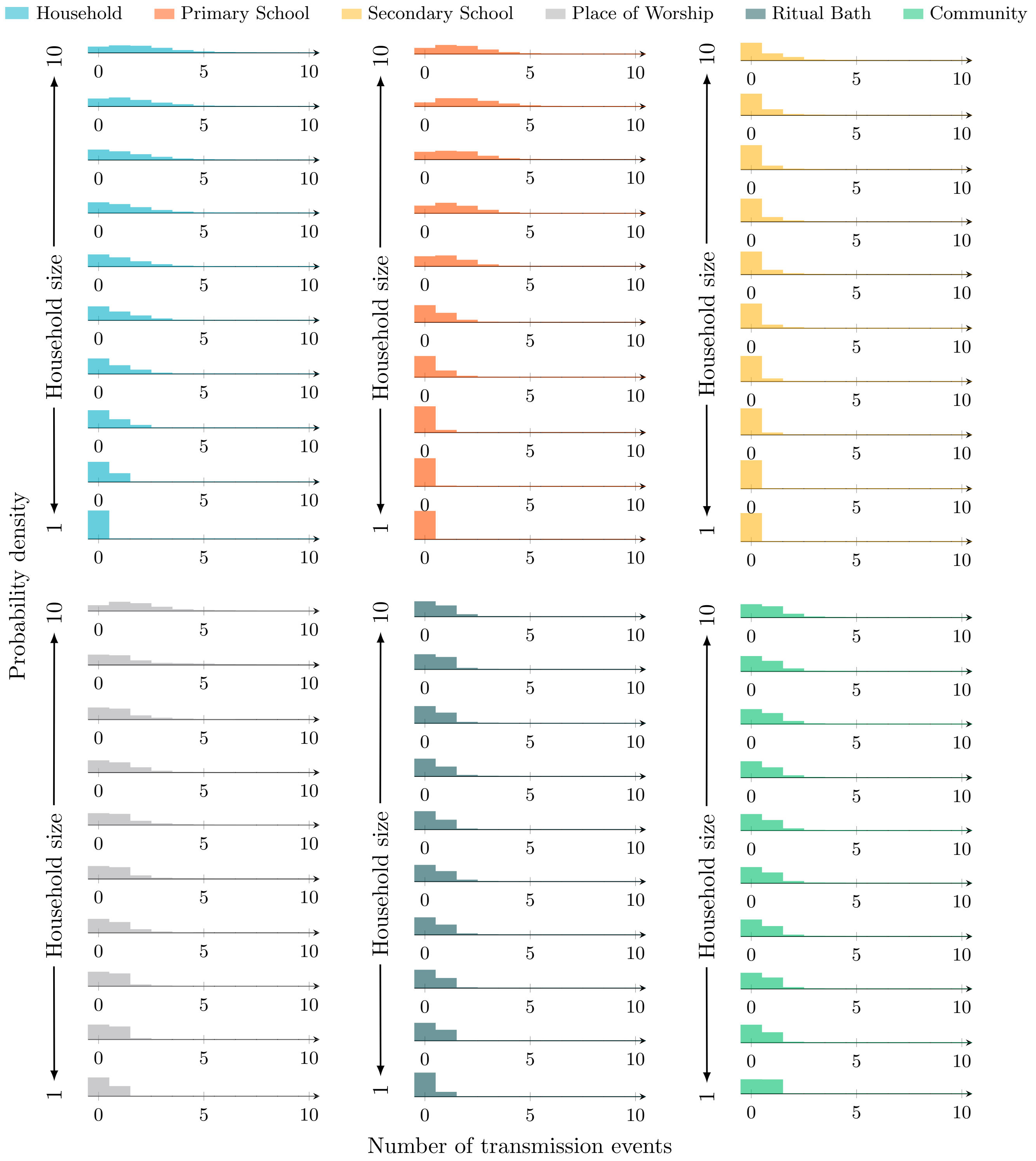


Figure 14: Transmission event distributions by source and household size – Splitting primary schools (60th percentile)

##### D.6 Splitting primary schools (50th percentile)

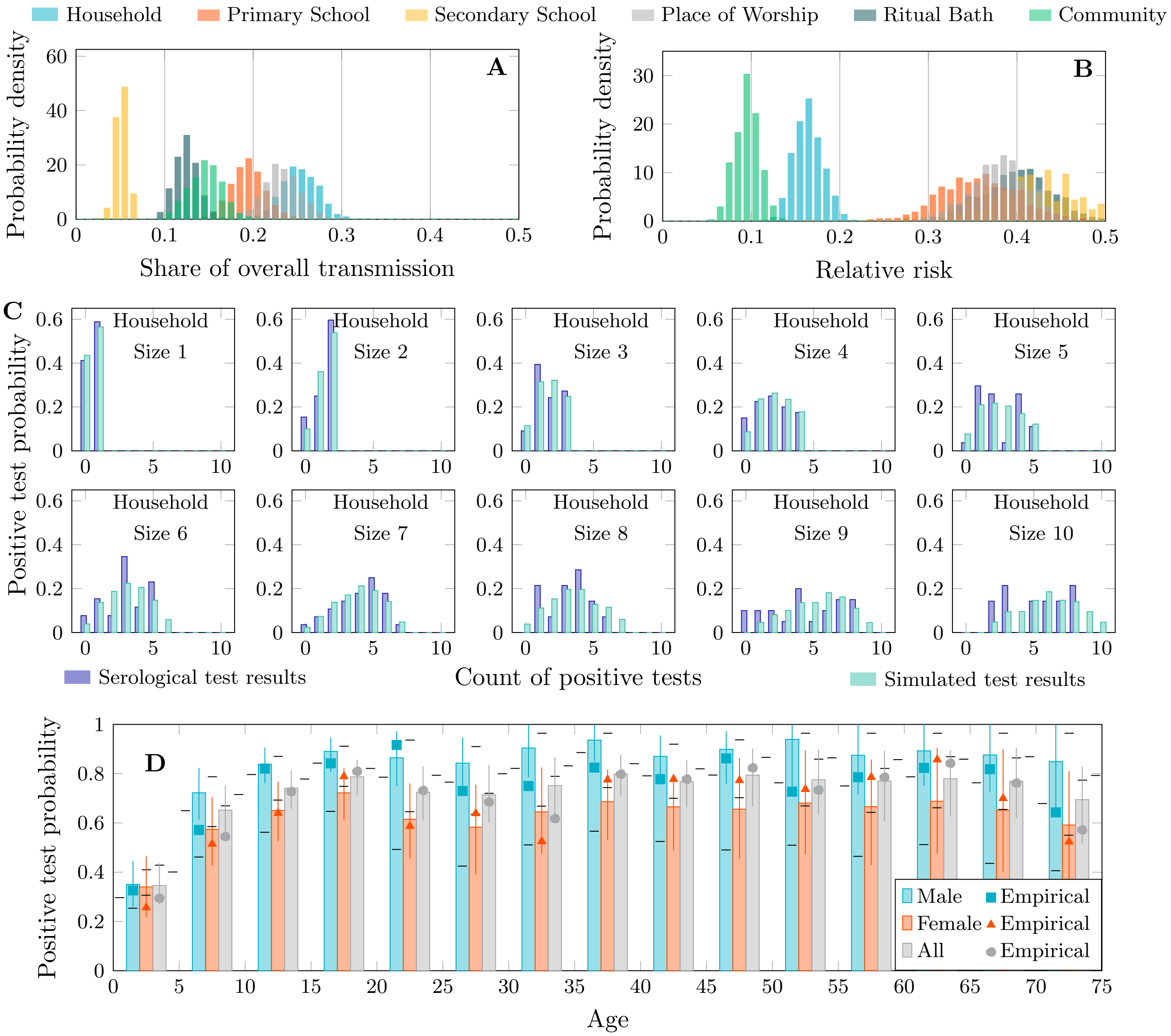


Figure 15: Transmission share, relative risk and postitive test distributions – Splitting primary schools (50th percentile)

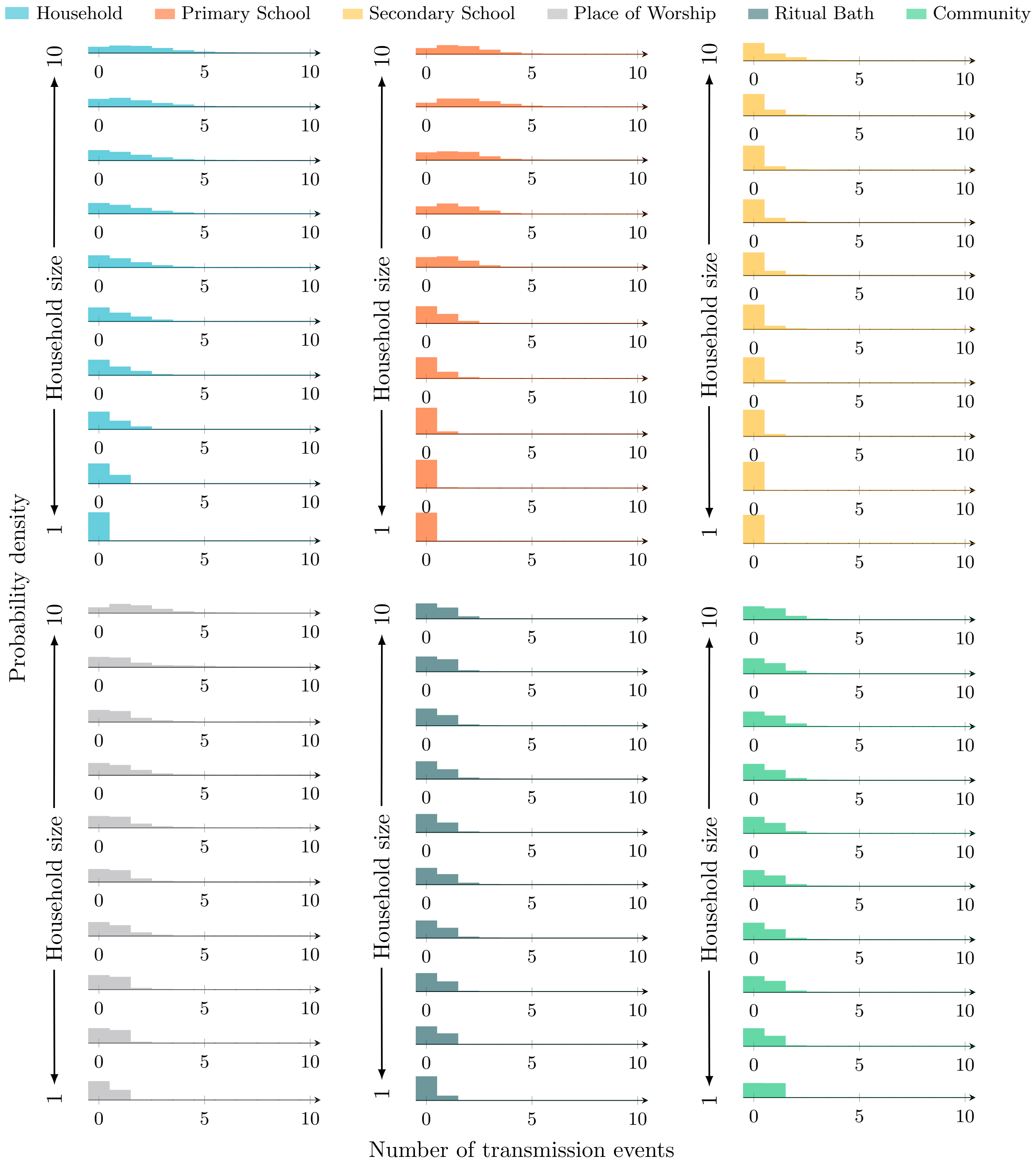


Figure 16: Transmission event distributions by source and household size – Splitting primary schools (50th percentile)

#### D.7 Splitting primary schools (40th percentile)

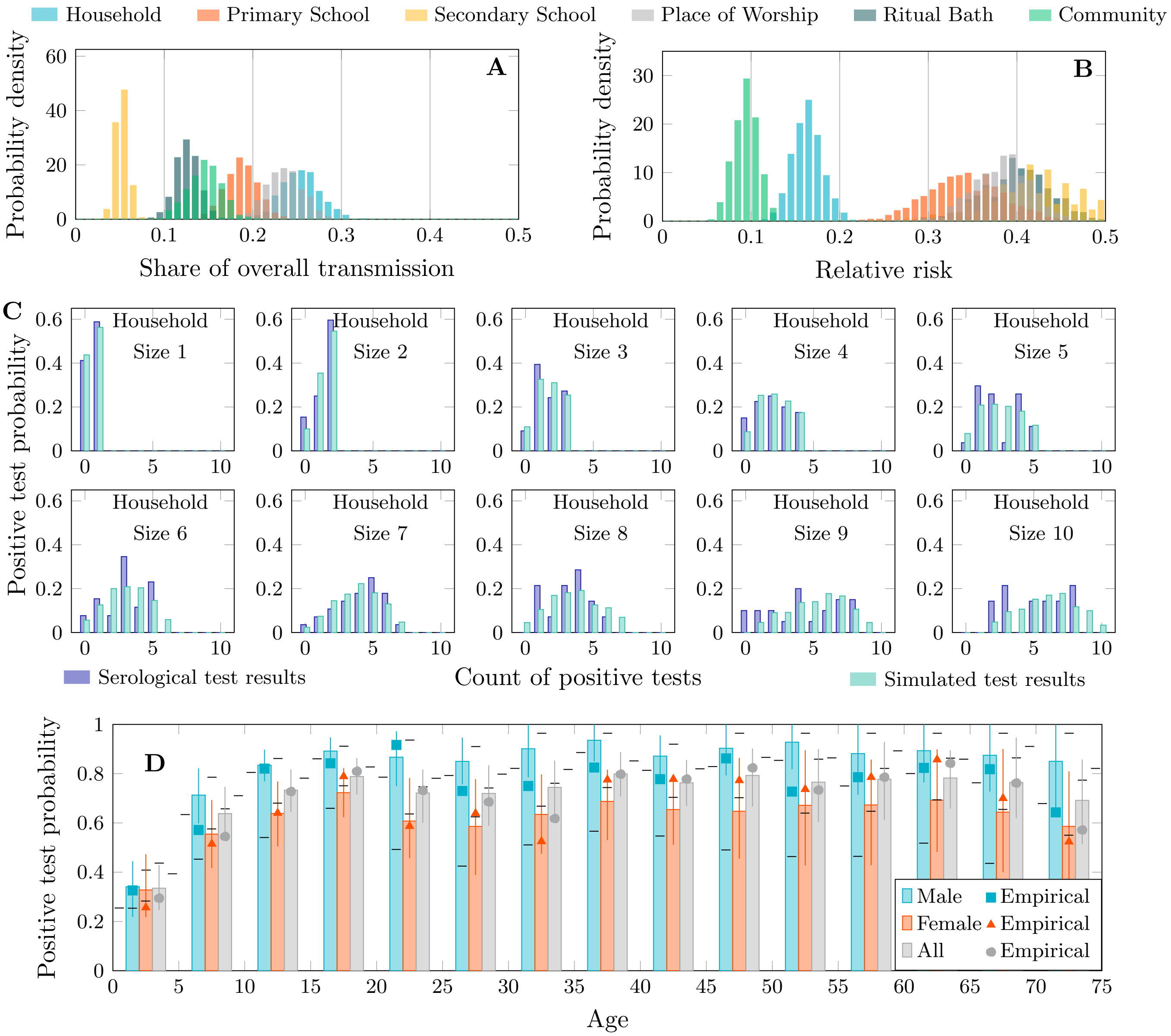


Figure 17: Transmission share, relative risk and postitive test distributions – Splitting primary schools (40th percentile)

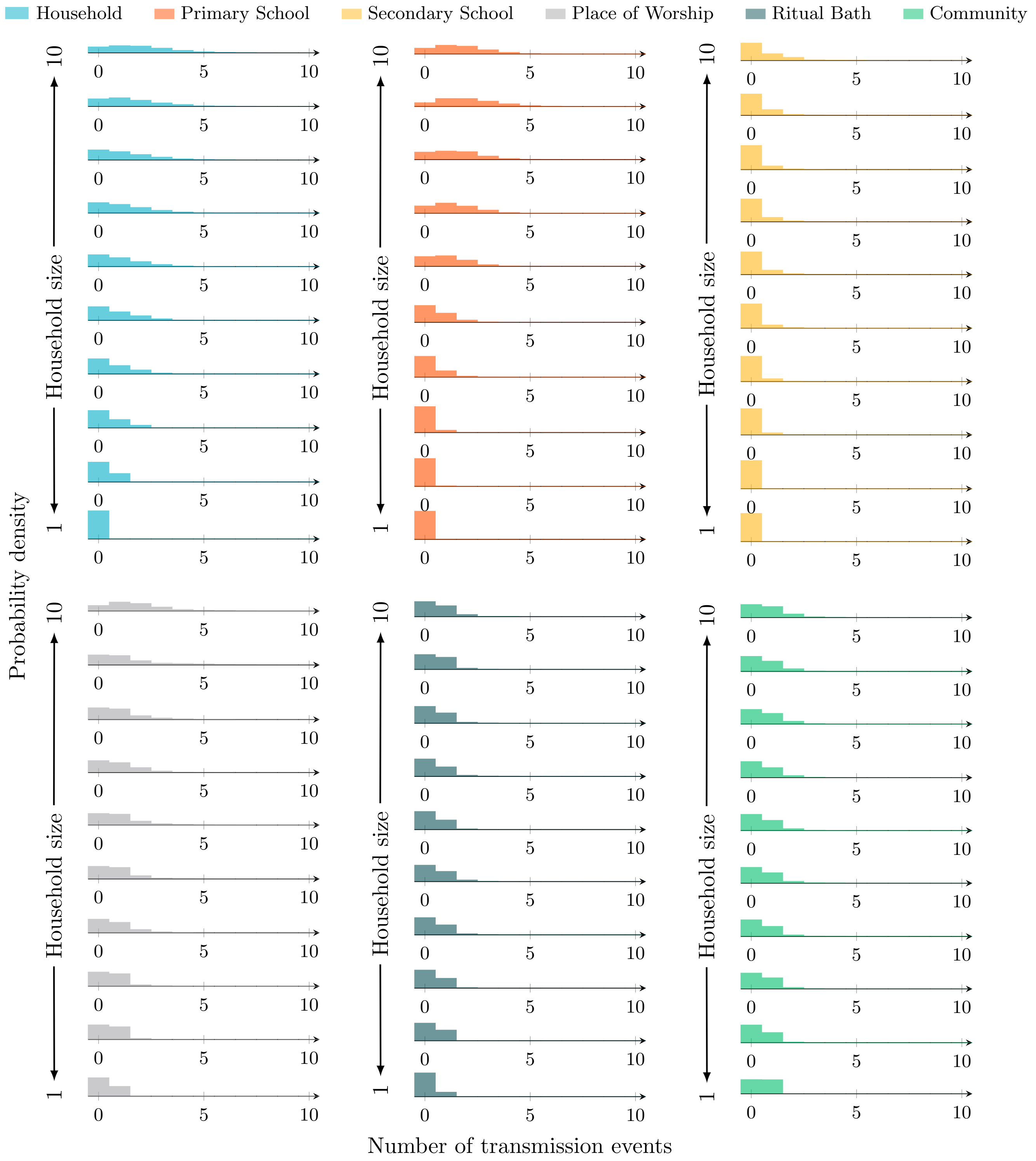


Figure 18: Transmission event distributions by source and household size – Splitting primary schools (40th percentile)

#### D.8 Splitting primary schools (30th percentile)

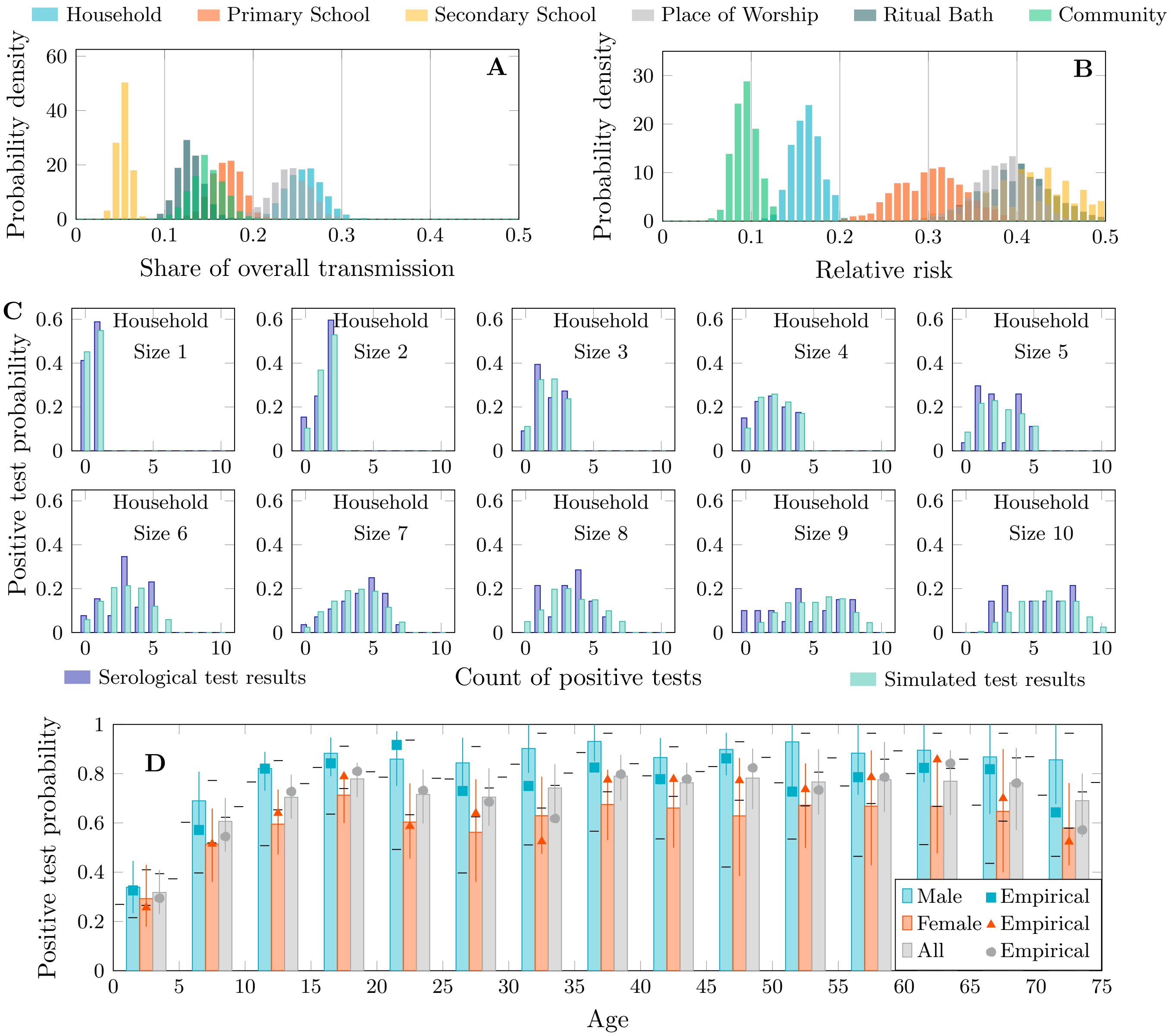


Figure 19: Transmission share, relative risk and postitive test distributions – Splitting primary schools (30th percentile)

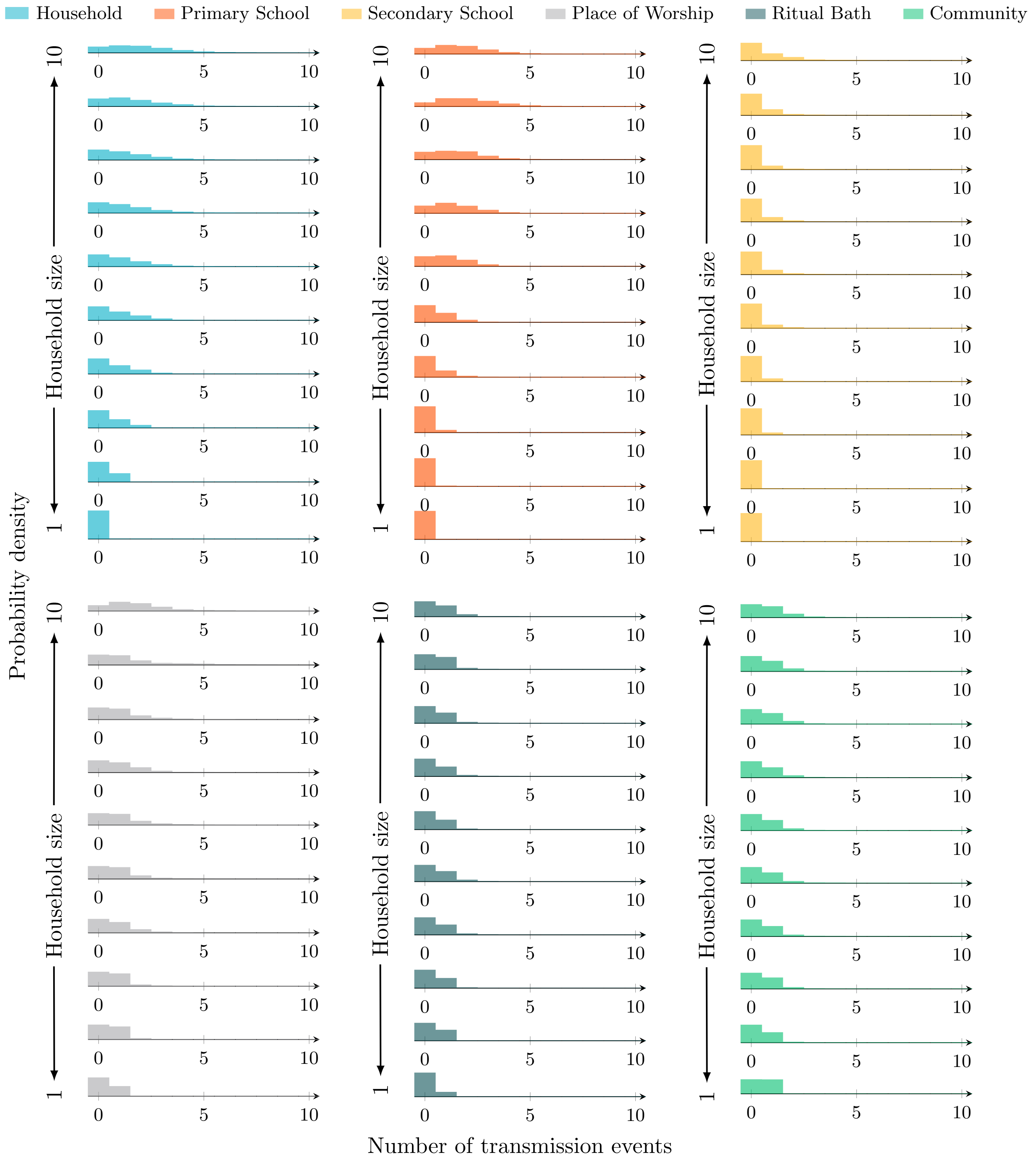


Figure 20: Transmission event distributions by source and household size – Splitting primary schools (30th percentile)

##### D.9 Splitting primary schools (20th percentile)

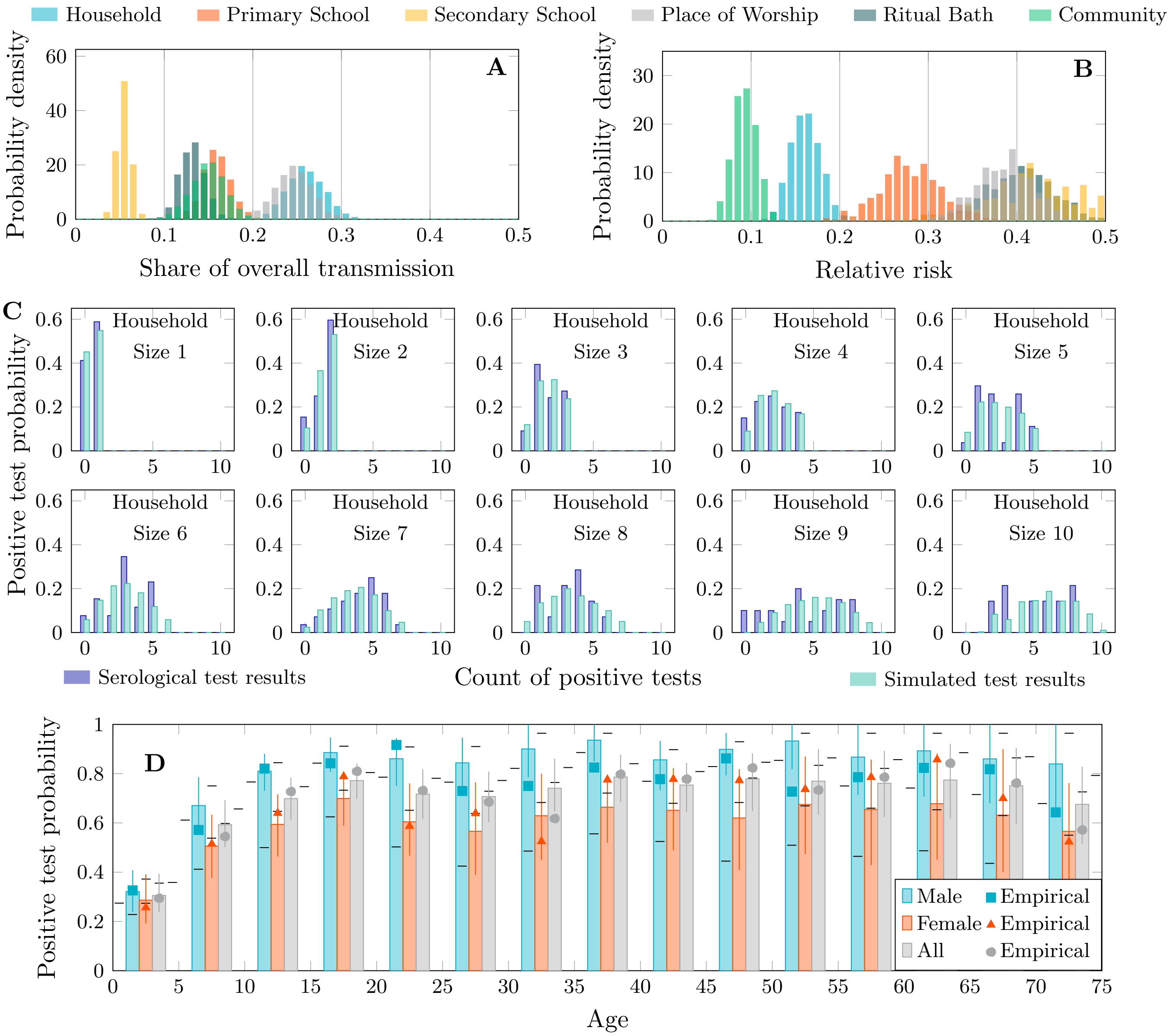


Figure 21: Transmission share, relative risk and postitive test distributions – Splitting primary schools (20th percentile)

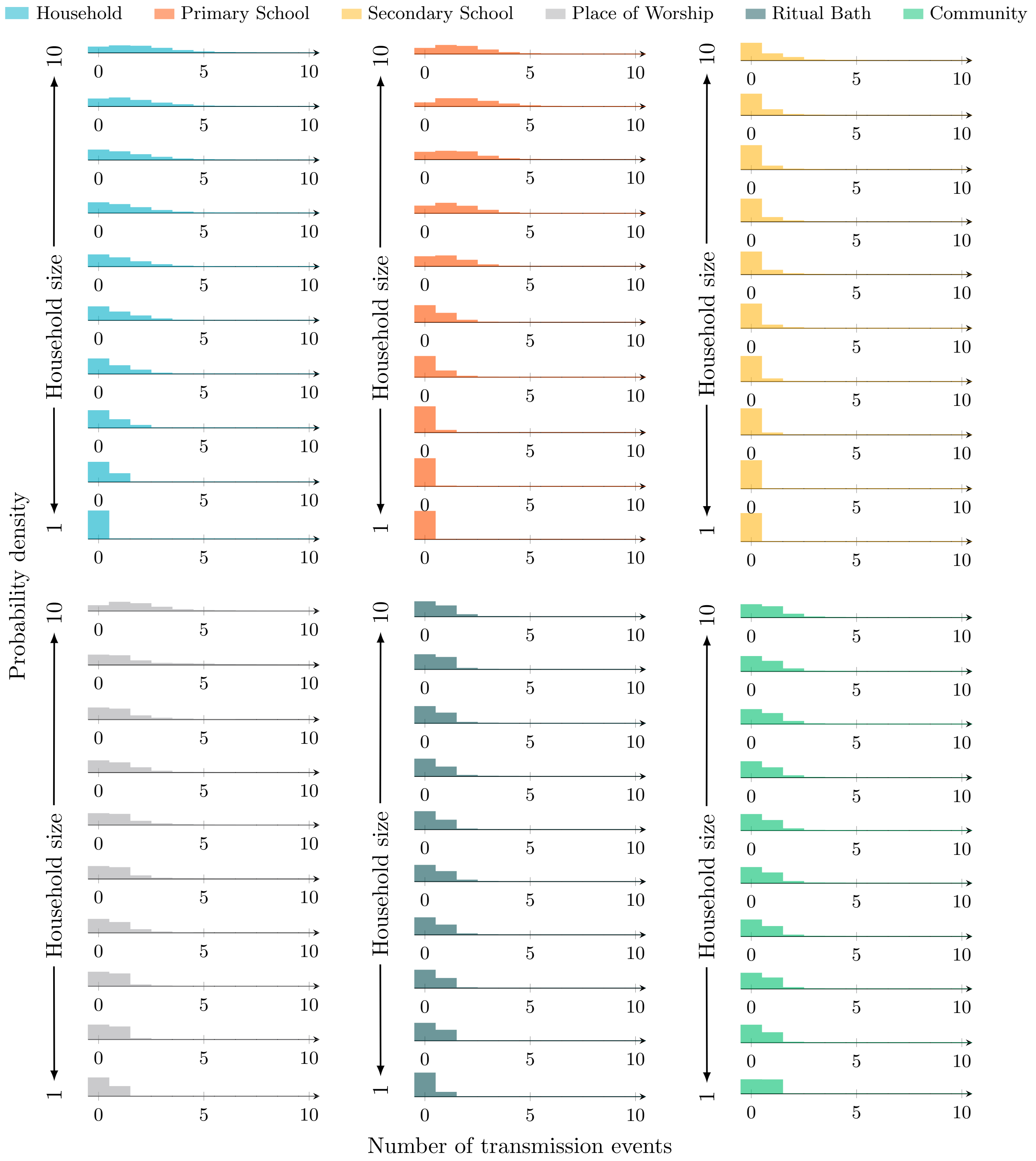


Figure 22: Transmission event distributions by source and household size – Splitting primary schools (20th percentile)

#### D.10 Splitting primary schools (10th percentile)

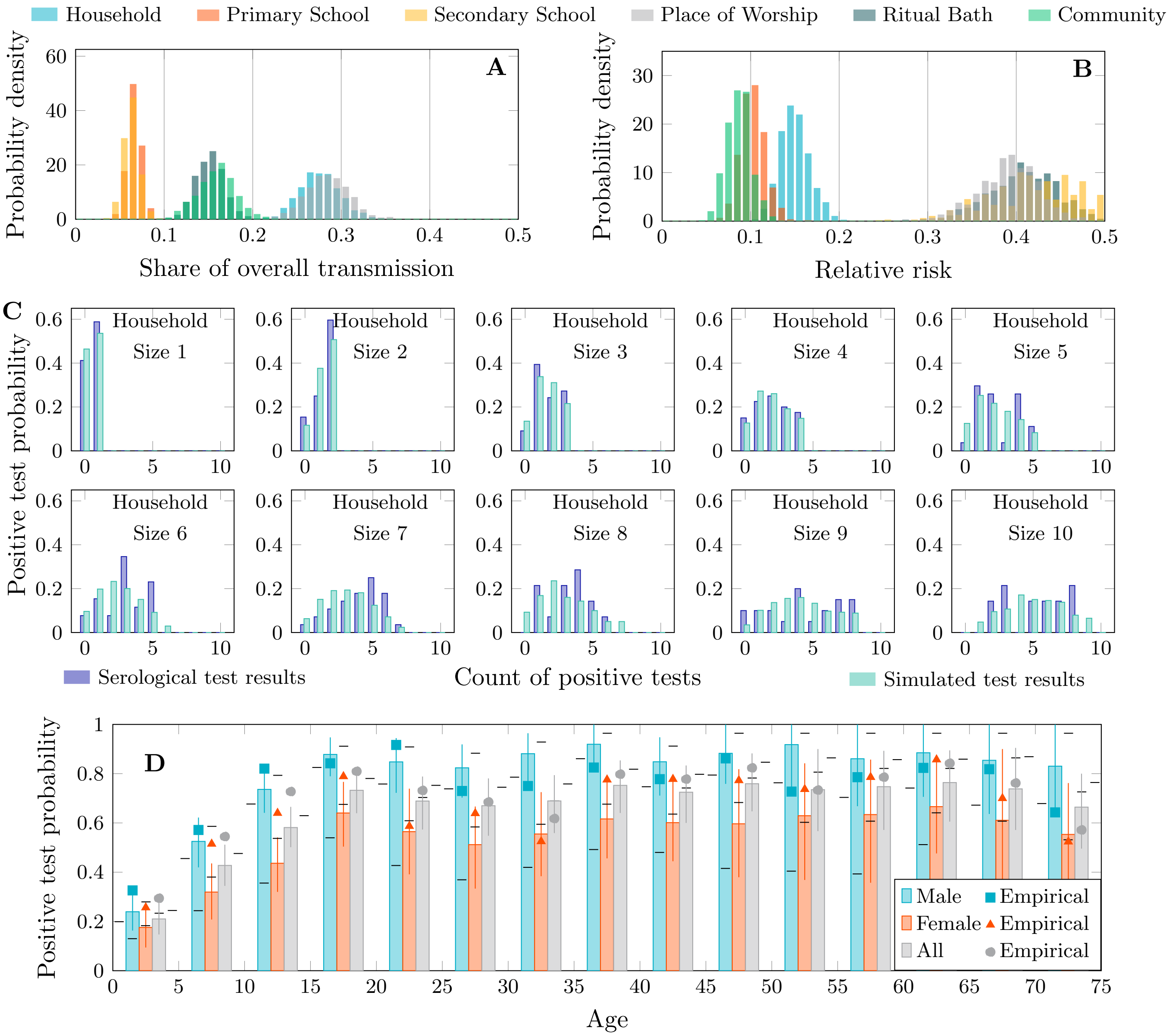


Figure 23: Transmission share, relative risk and postitive test distributions – Splitting primary schools (10th percentile)

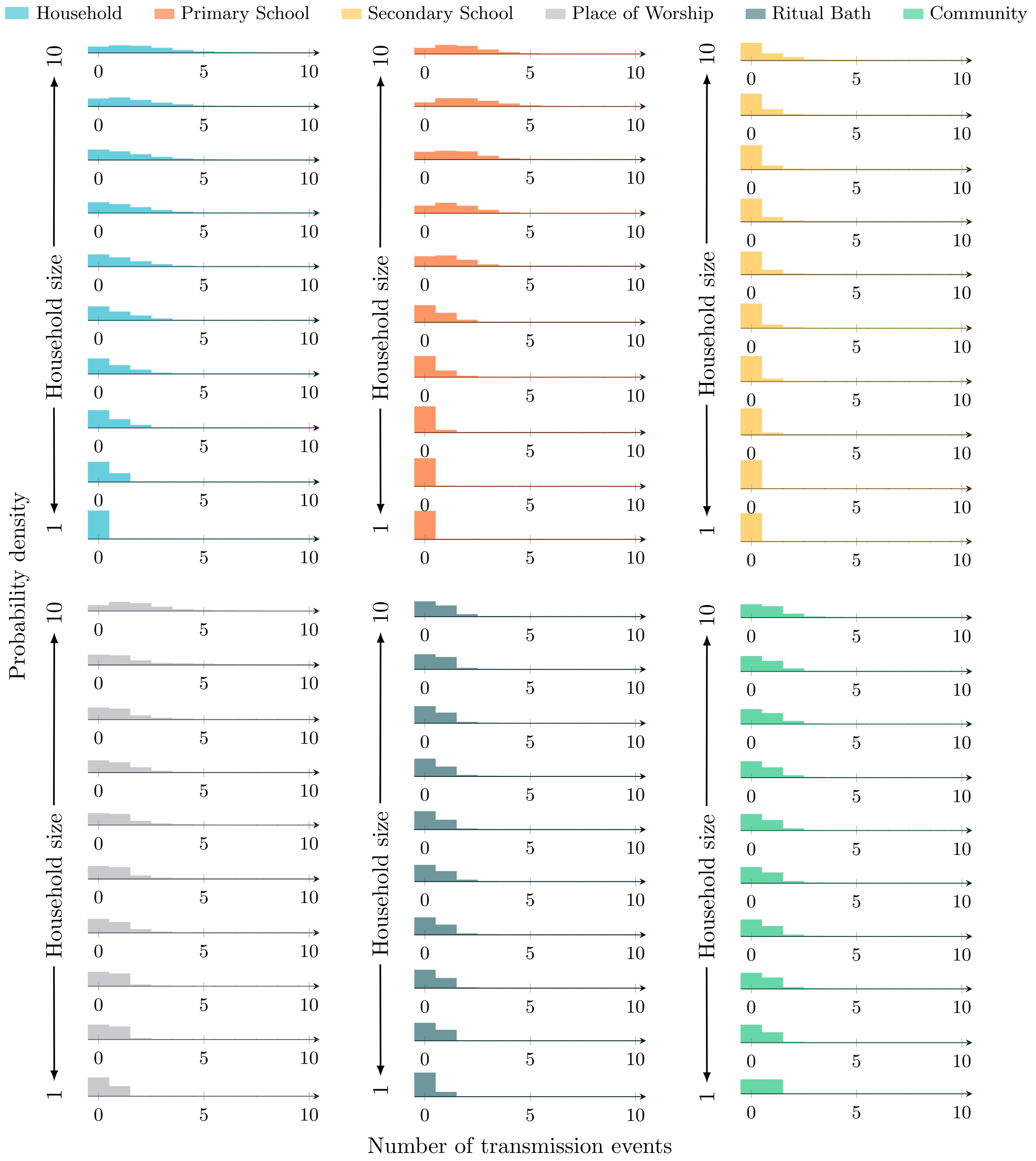


Figure 24: Transmission event distributions by source and household size – Splitting primary schools (10th percentile)

##### D.11 Splitting places of worship (90th percentile)

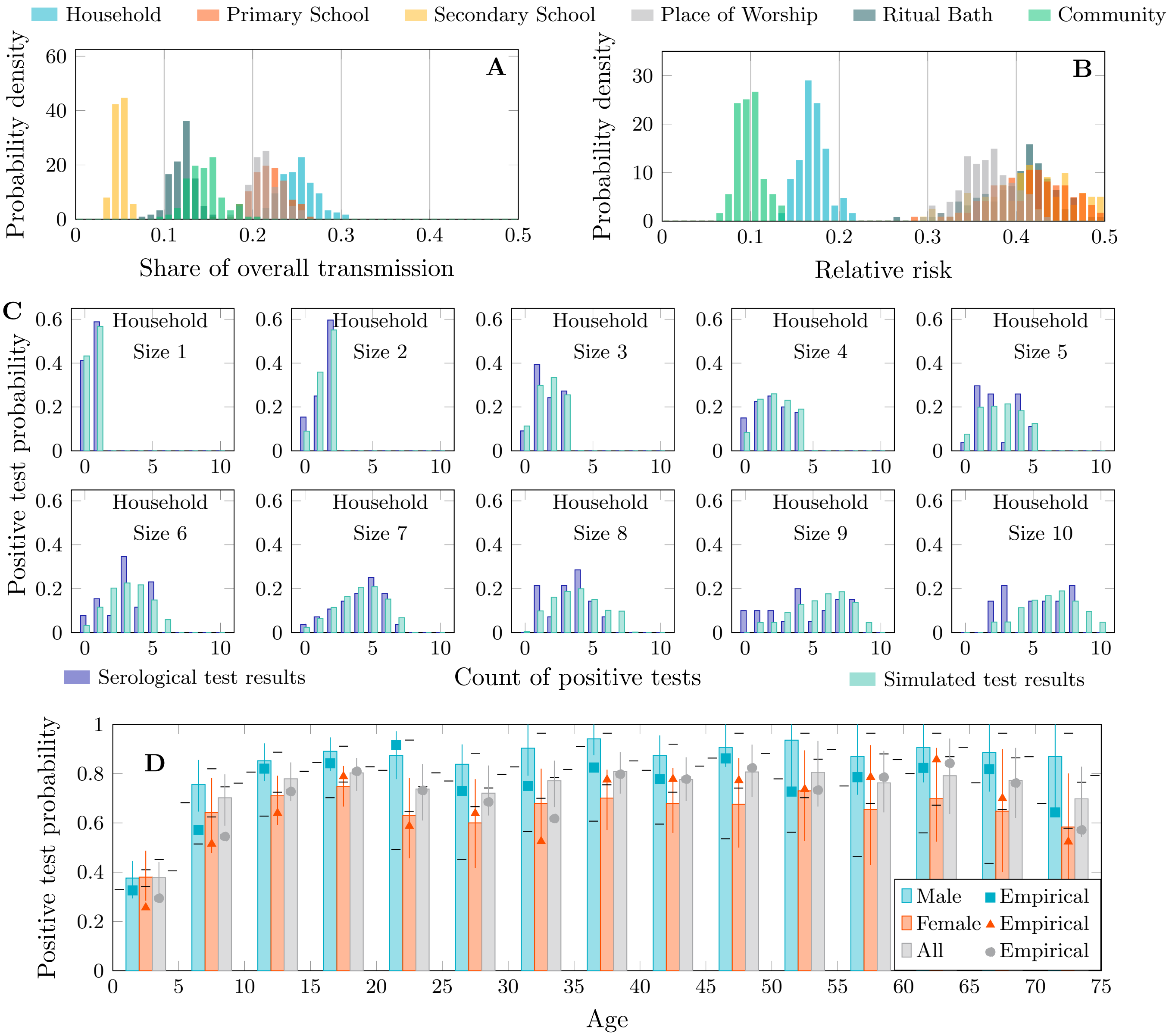


Figure 25: Transmission share, relative risk and postitive test distributions – Splitting places of worship (90th percentile)

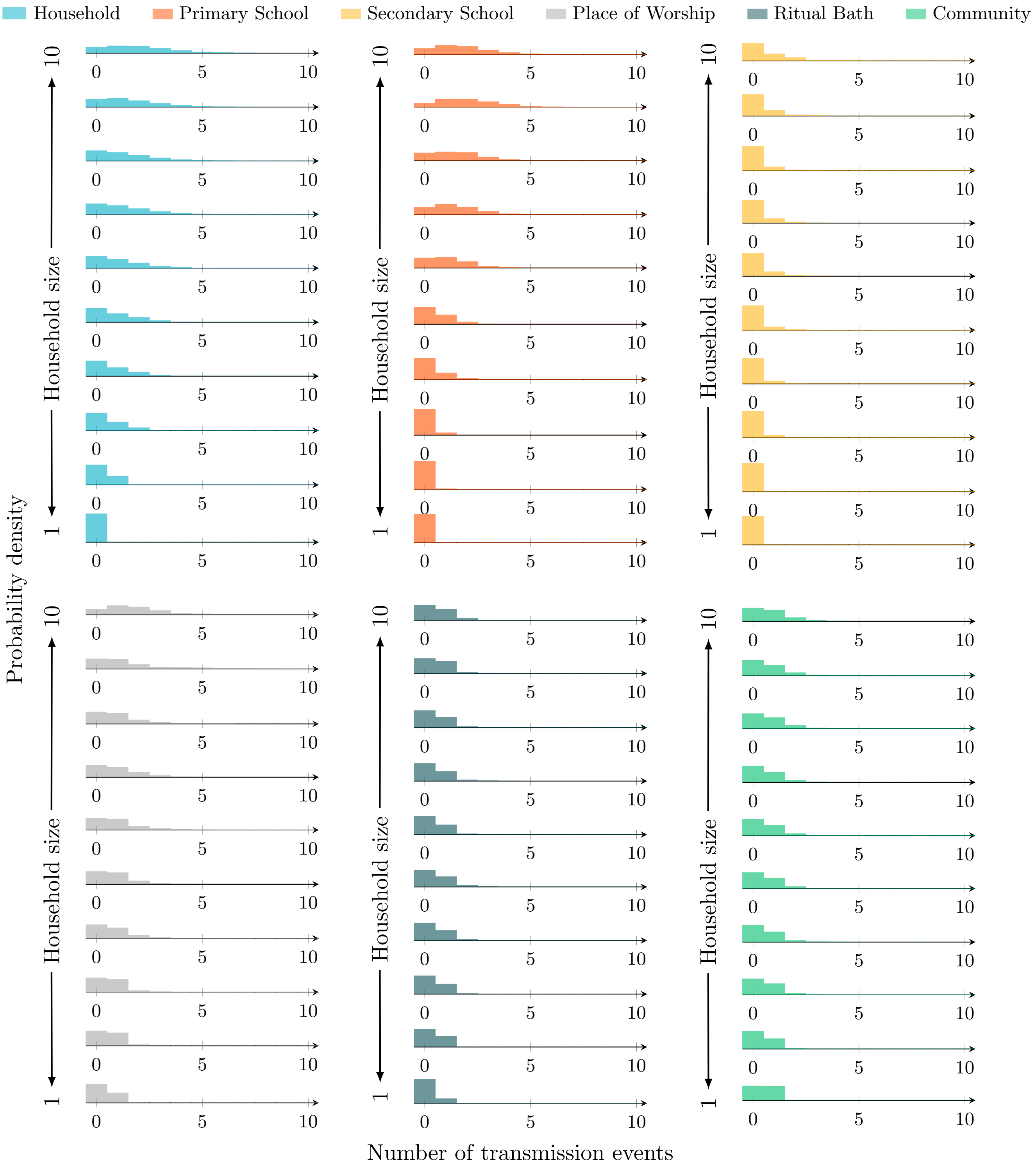


Figure 26: Transmission event distributions by source and household size – Splitting places of worship (90th percentile)

#### D.12 Splitting places of worship (80th percentile)

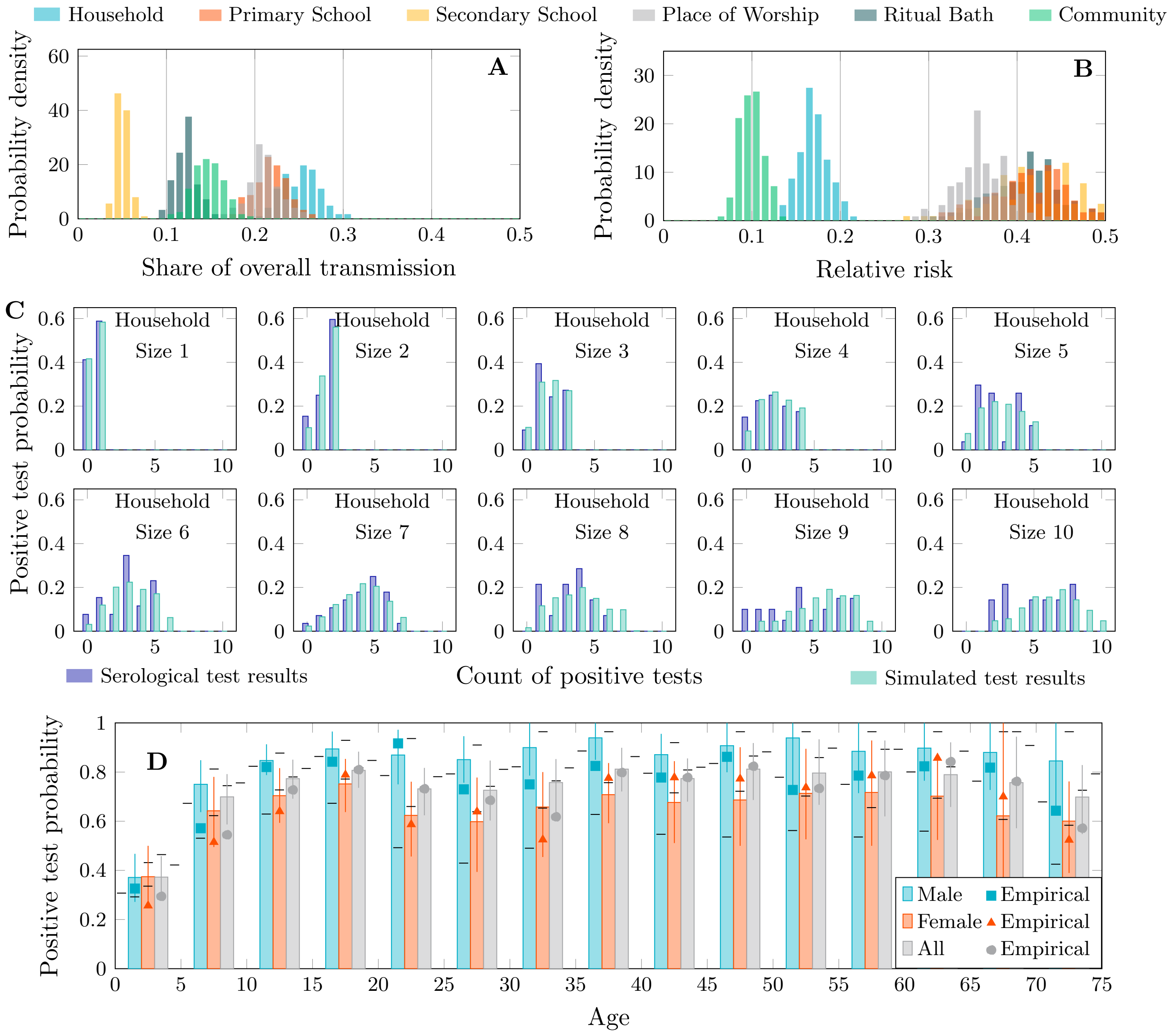


Figure 27: Transmission share, relative risk and postitive test distributions – Splitting places of worship (80th percentile)

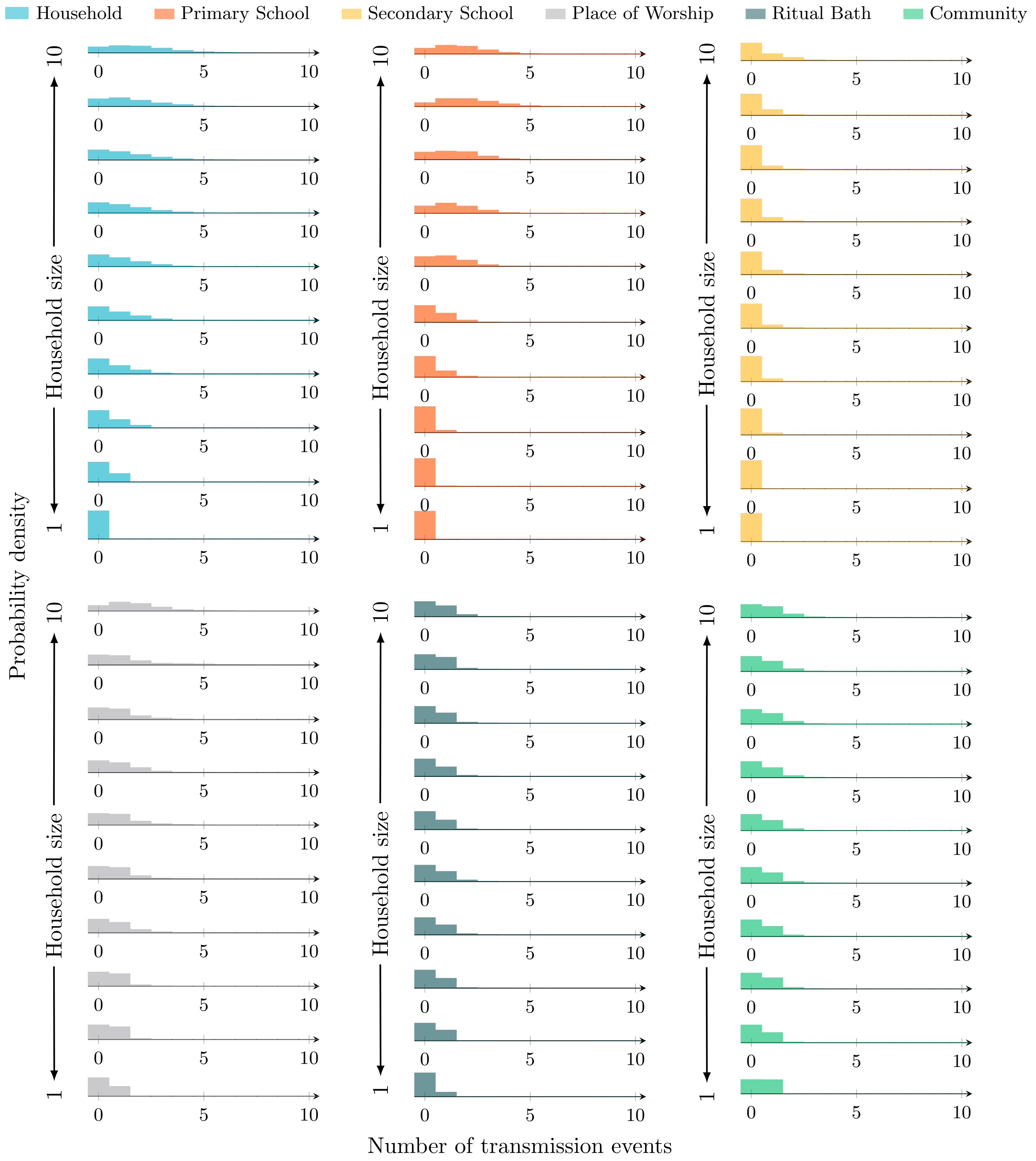


Figure 28: Transmission event distributions by source and household size – Splitting places of worship (80th percentile)

#### D.13 Splitting places of worship (70th percentile)

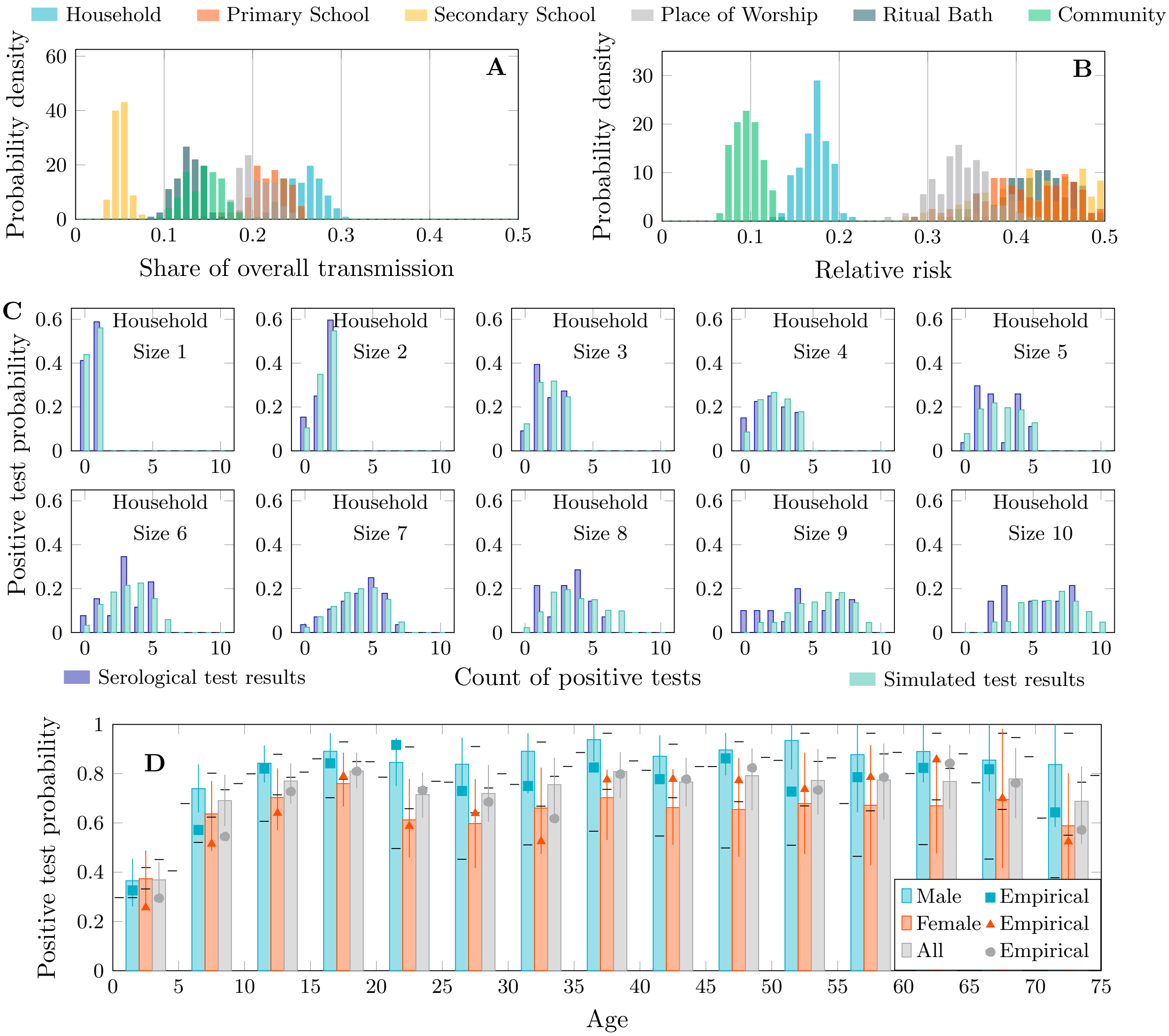


Figure 29: Transmission share, relative risk and postitive test distributions – Splitting places of worship (70th percentile)

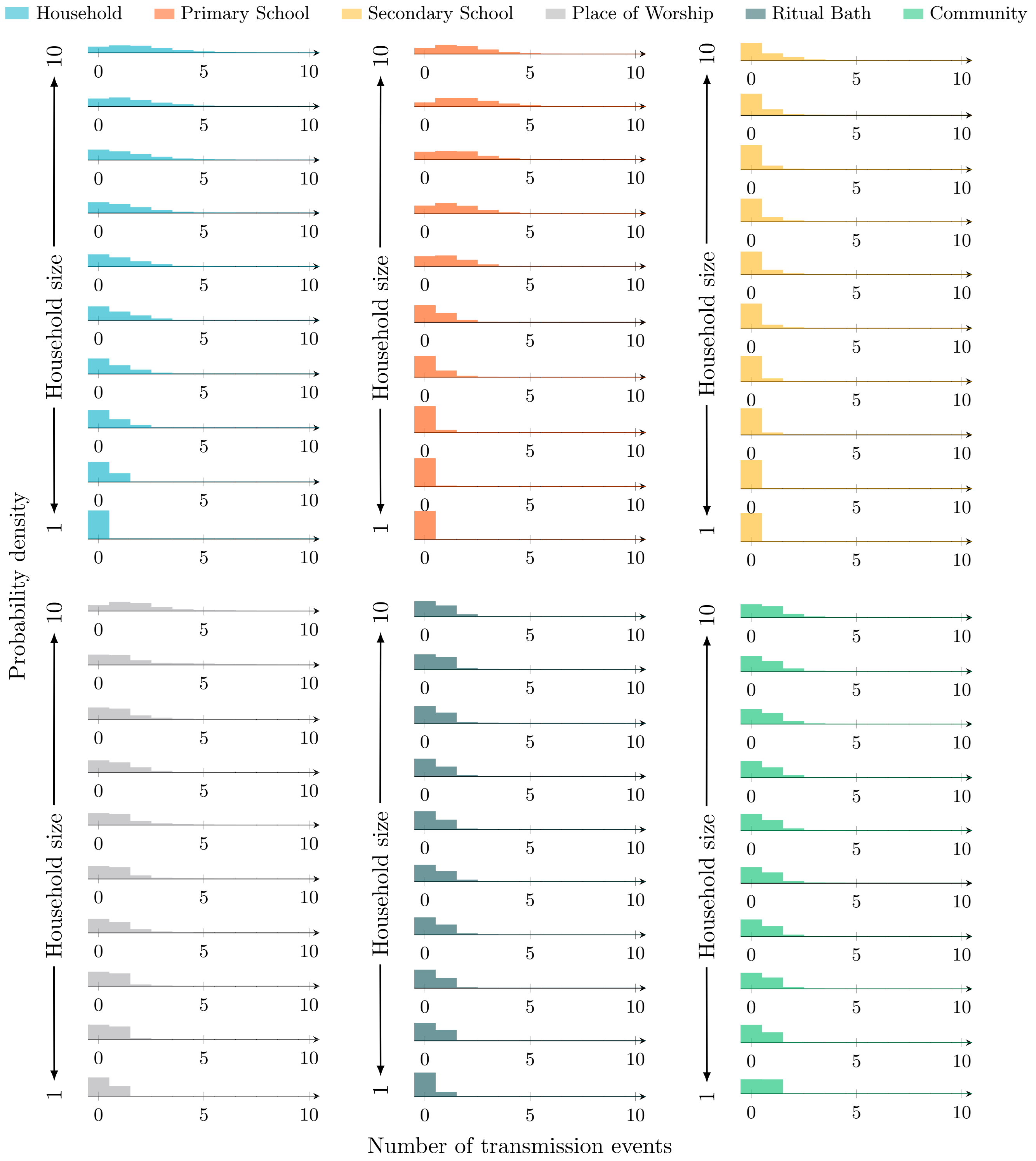


Figure 30: Transmission event distributions by source and household size – Splitting places of worship (70th percentile)

#### D.14 Splitting places of worship (60th percentile)

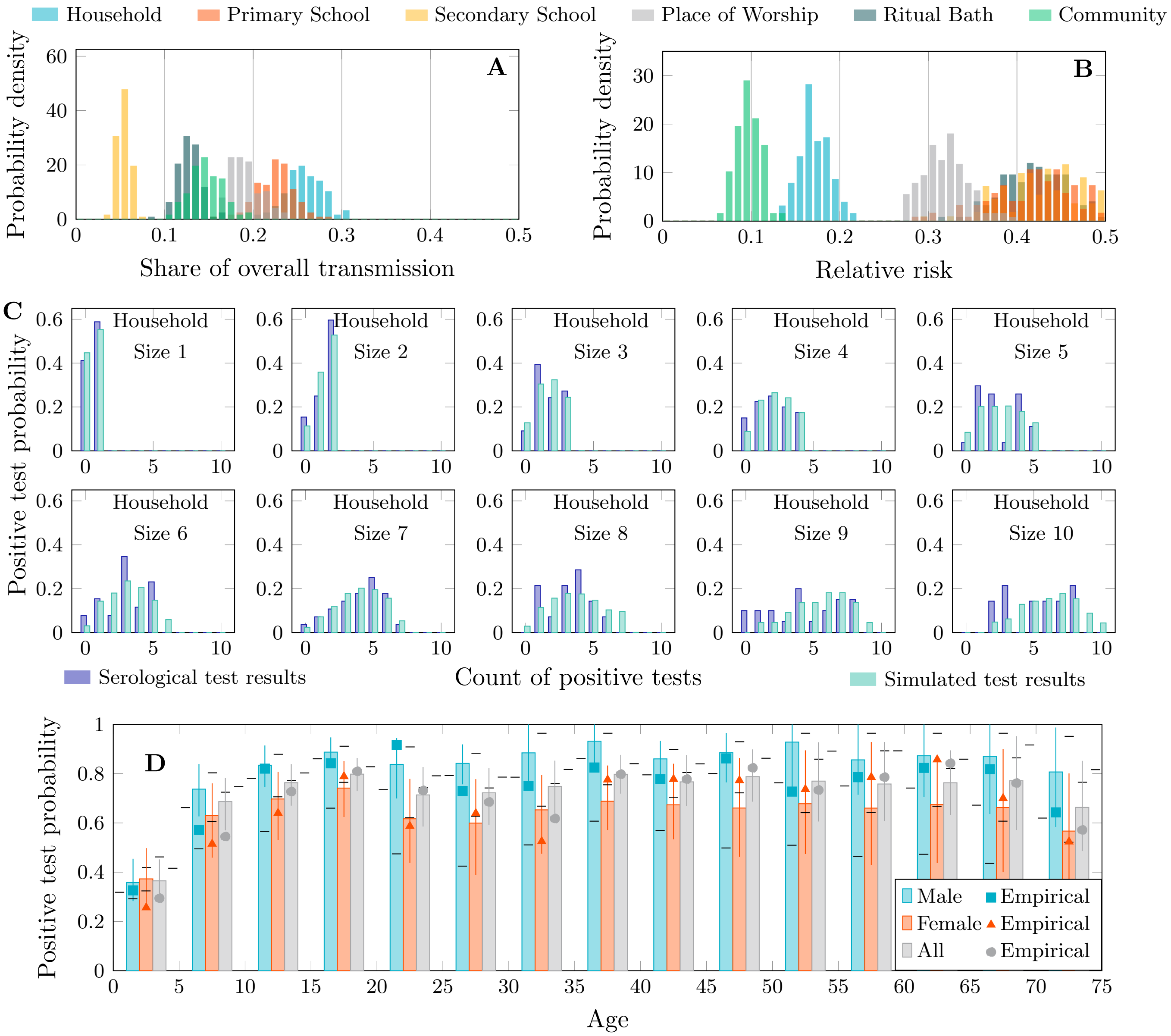


Figure 31: Transmission share, relative risk and postitive test distributions – Splitting places of worship (60th percentile)

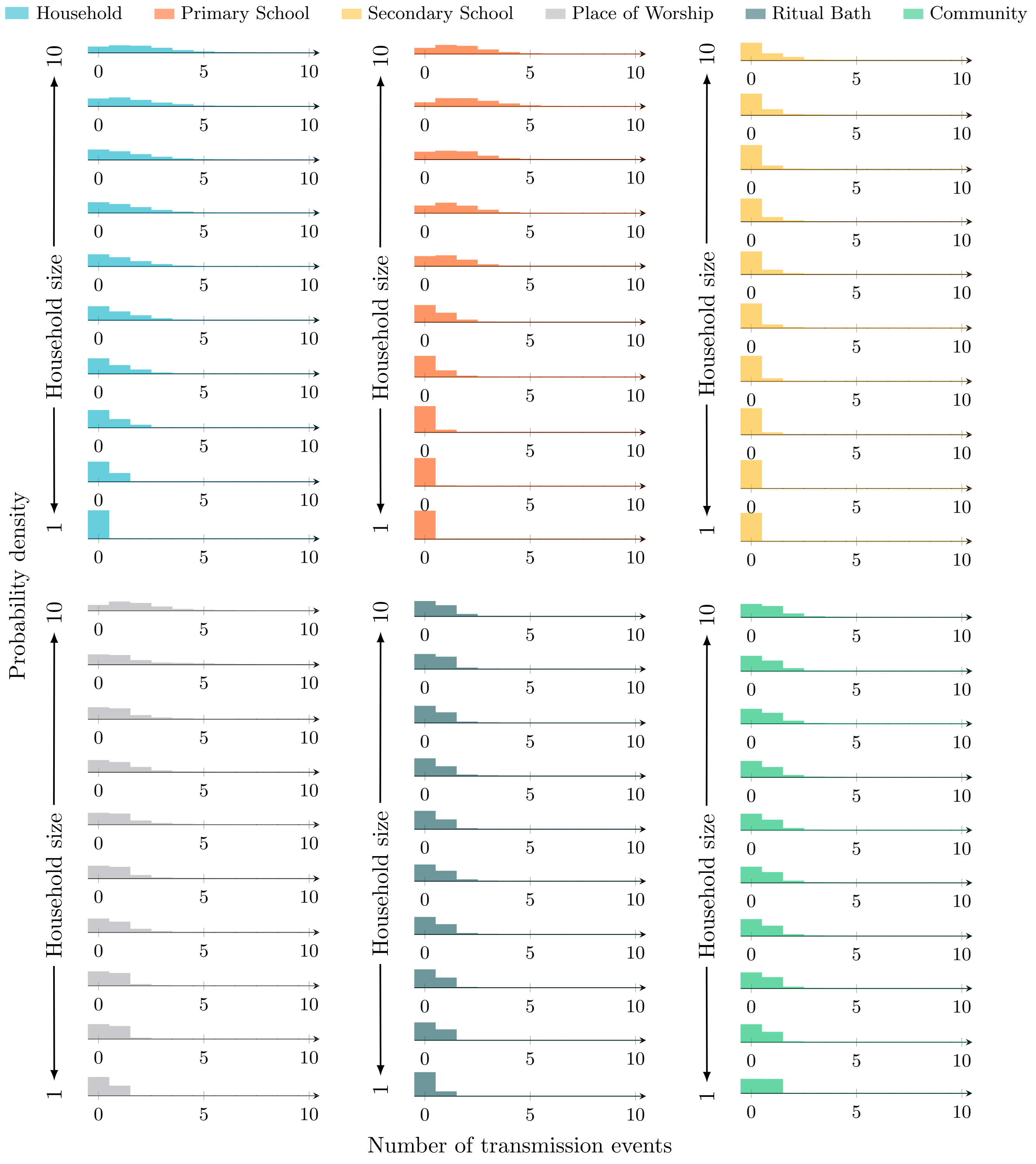


Figure 32: Transmission event distributions by source and household size – Splitting places of worship (60th percentile)

#### D.15 Splitting places of worship (50th percentile)

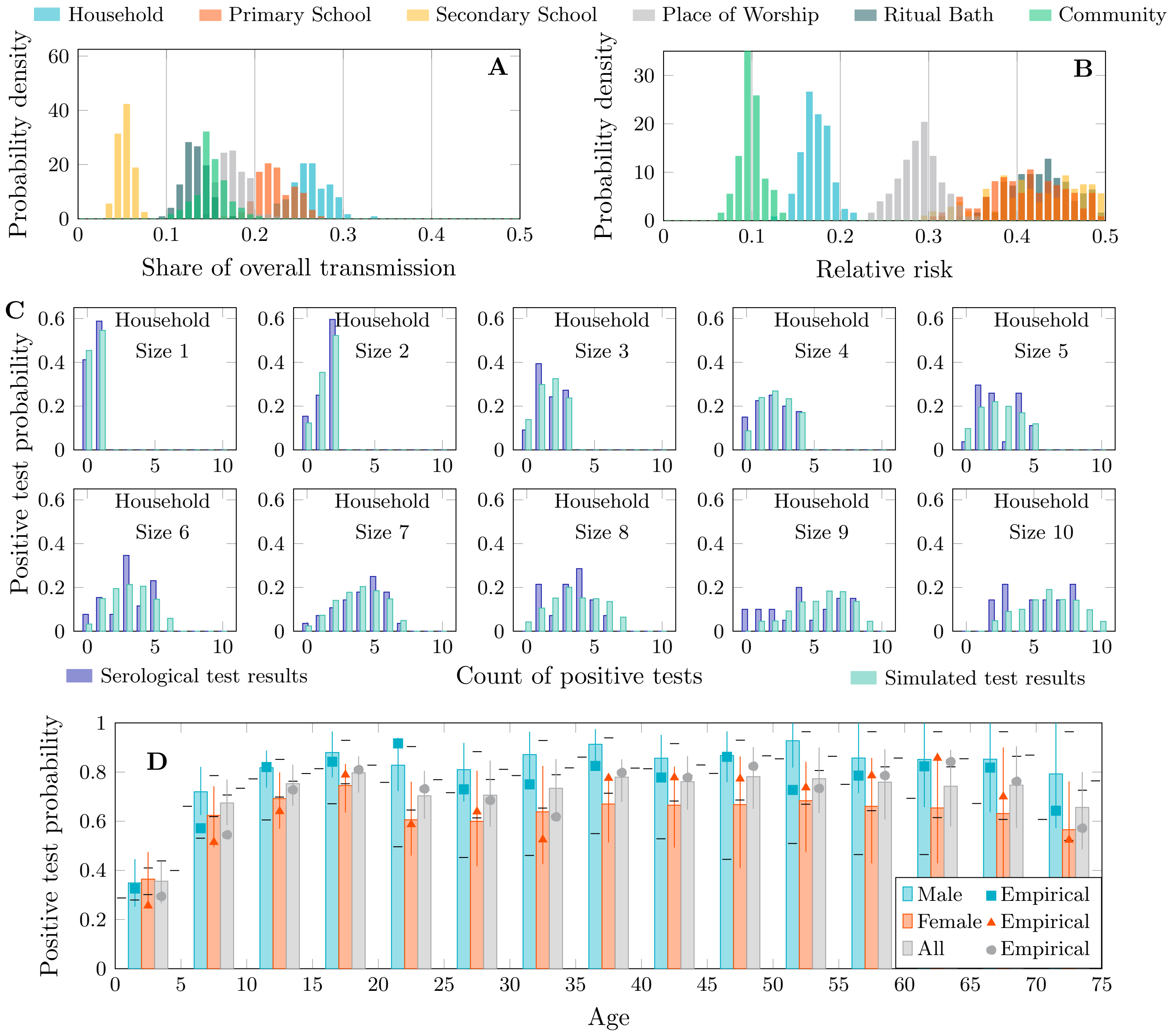


Figure 33: Transmission share, relative risk and postitive test distributions – Splitting places of worship (50th percentile)

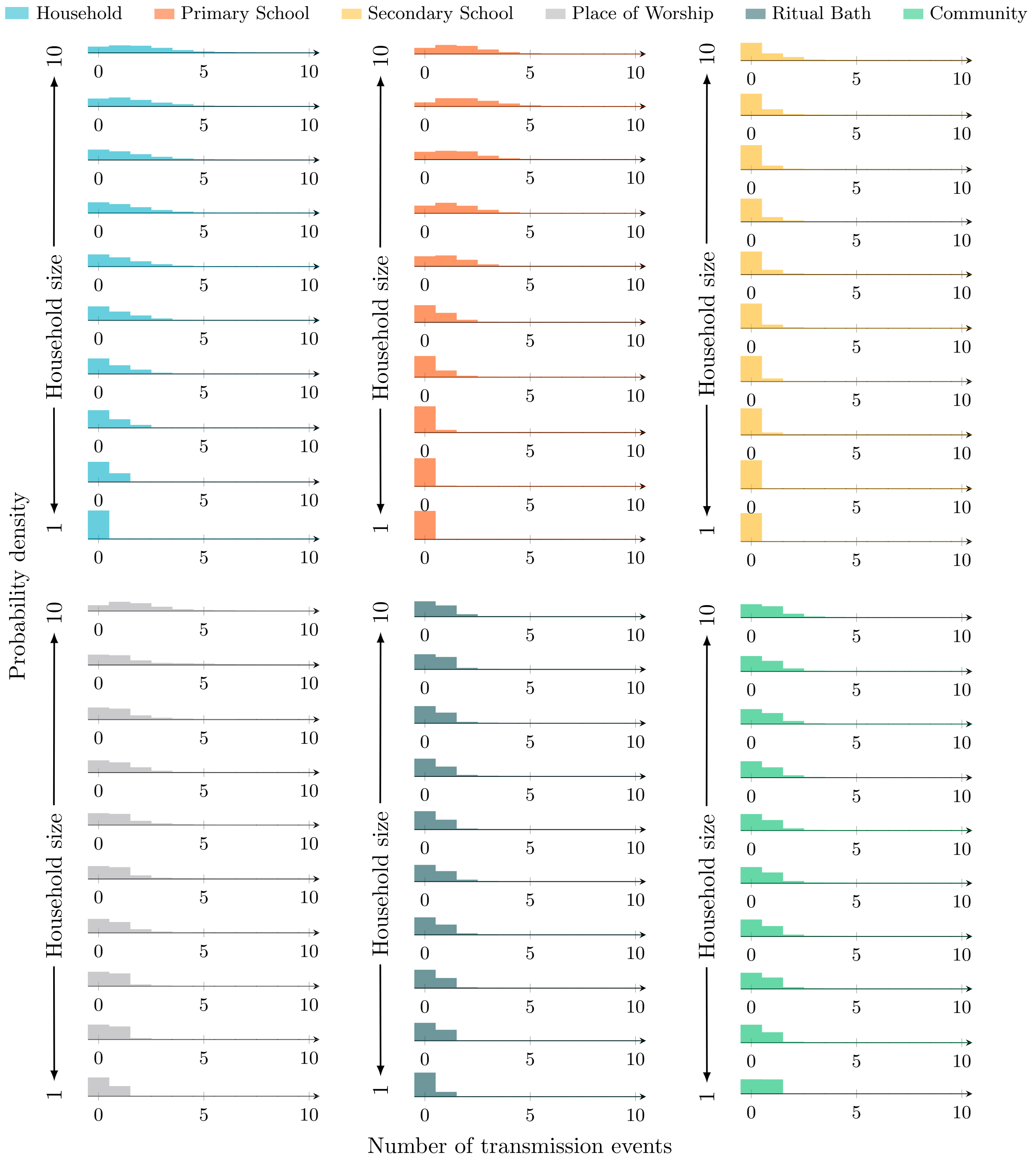


Figure 34: Transmission event distributions by source and household size – Splitting places of worship (50th percentile)

##### D.16 Splitting places of worship (40th percentile)

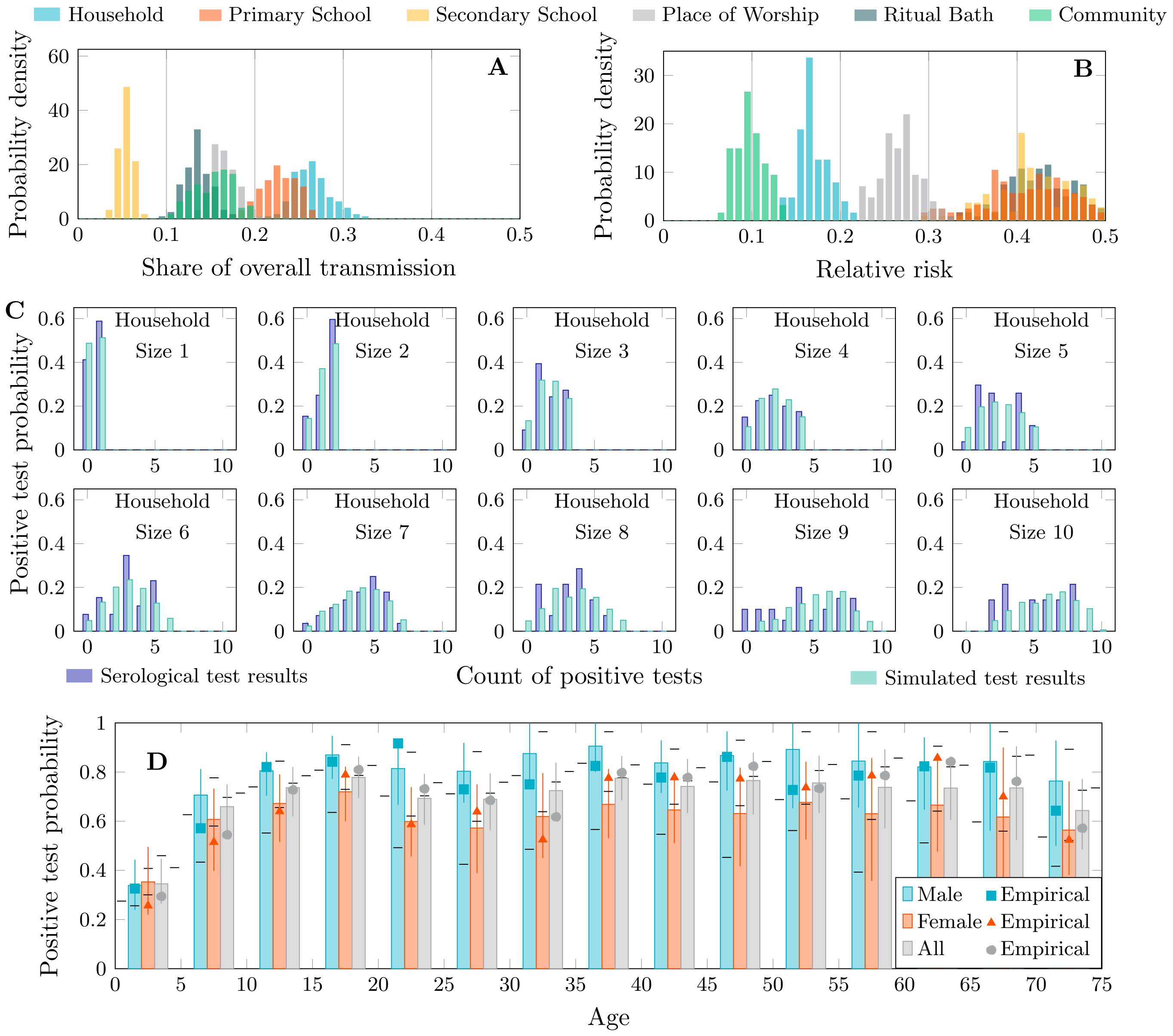


Figure 35: Transmission share, relative risk and postitive test distributions – Splitting places of worship (40th percentile)

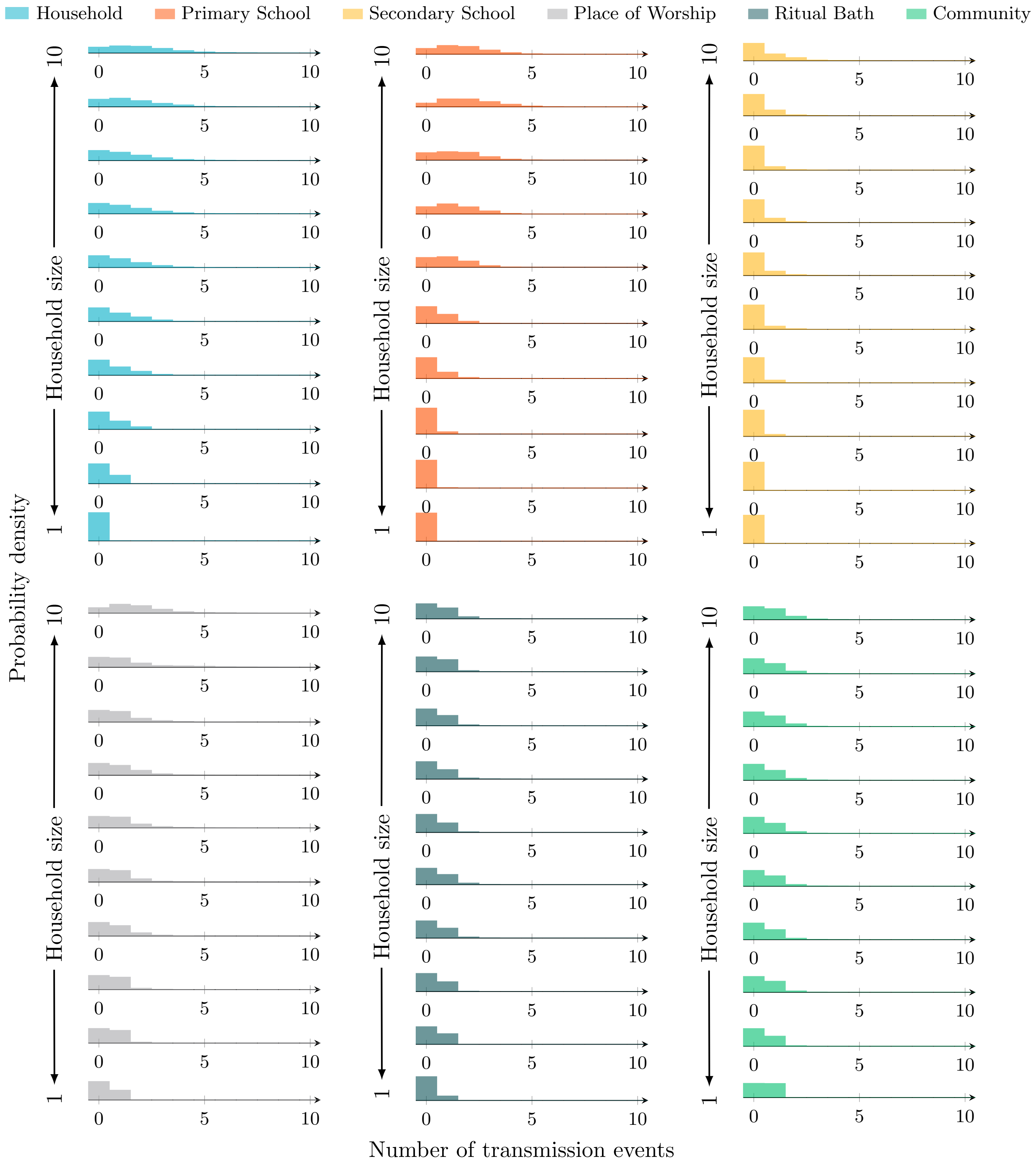


Figure 36: Transmission event distributions by source and household size – Splitting places of worship (40th percentile)

#### D.17 Splitting places of worship (30th percentile)

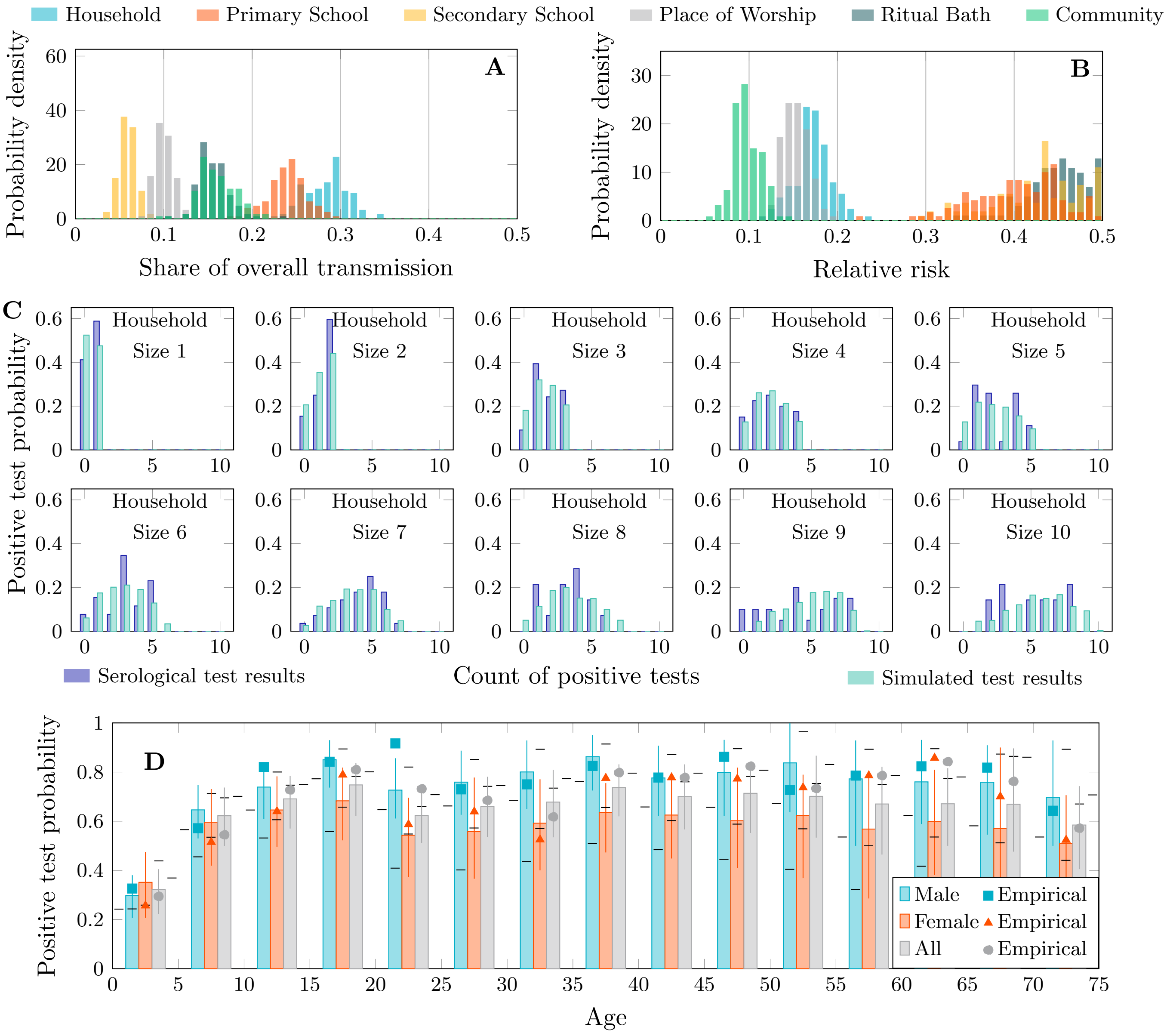


Figure 37: Transmission share, relative risk and postitive test distributions – Splitting places of worship (30th percentile)

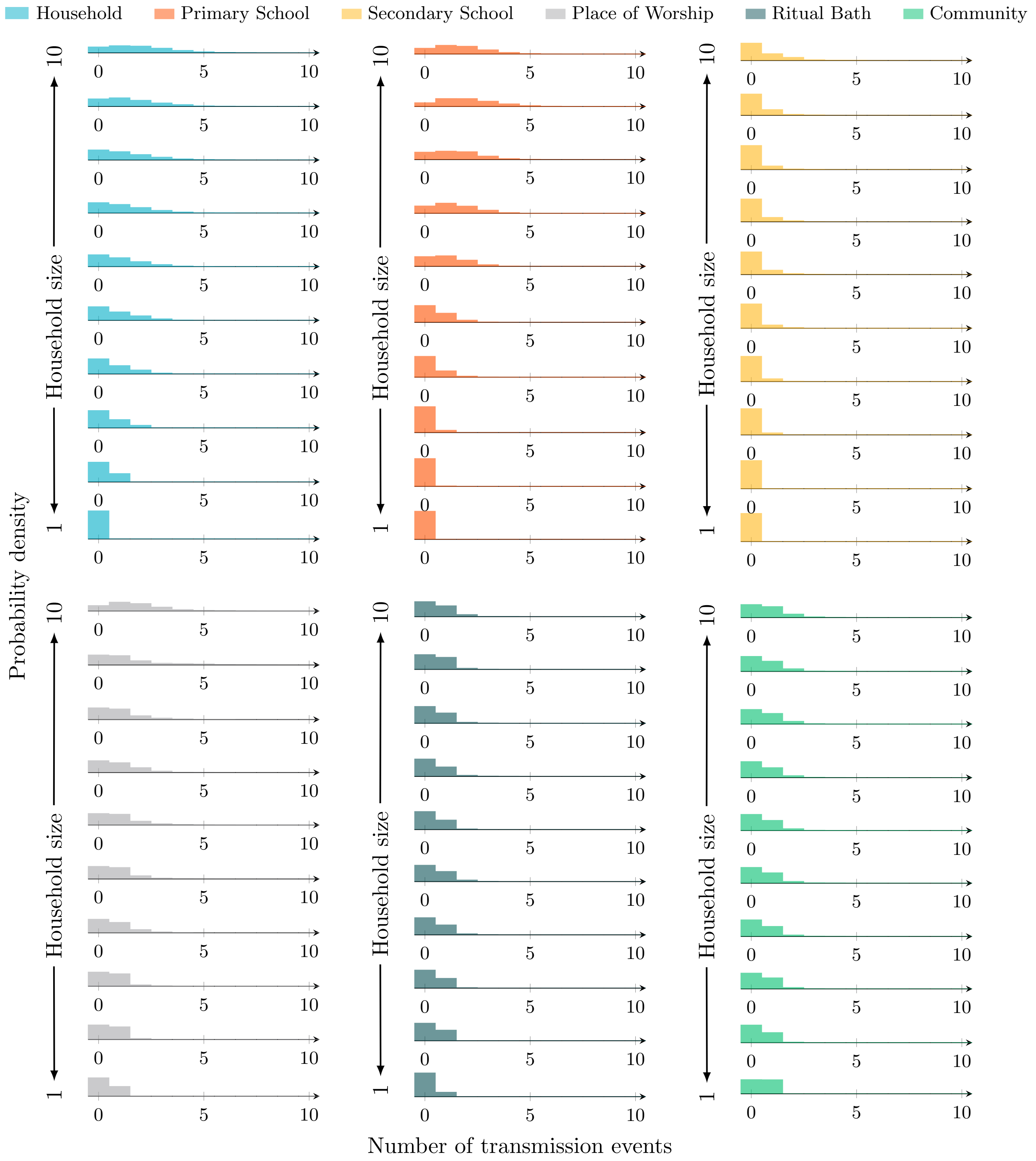


Figure 38: Transmission event distributions by source and household size – Splitting places of worship (30th percentile)

#### D.18 Splitting places of worship (20th percentile)

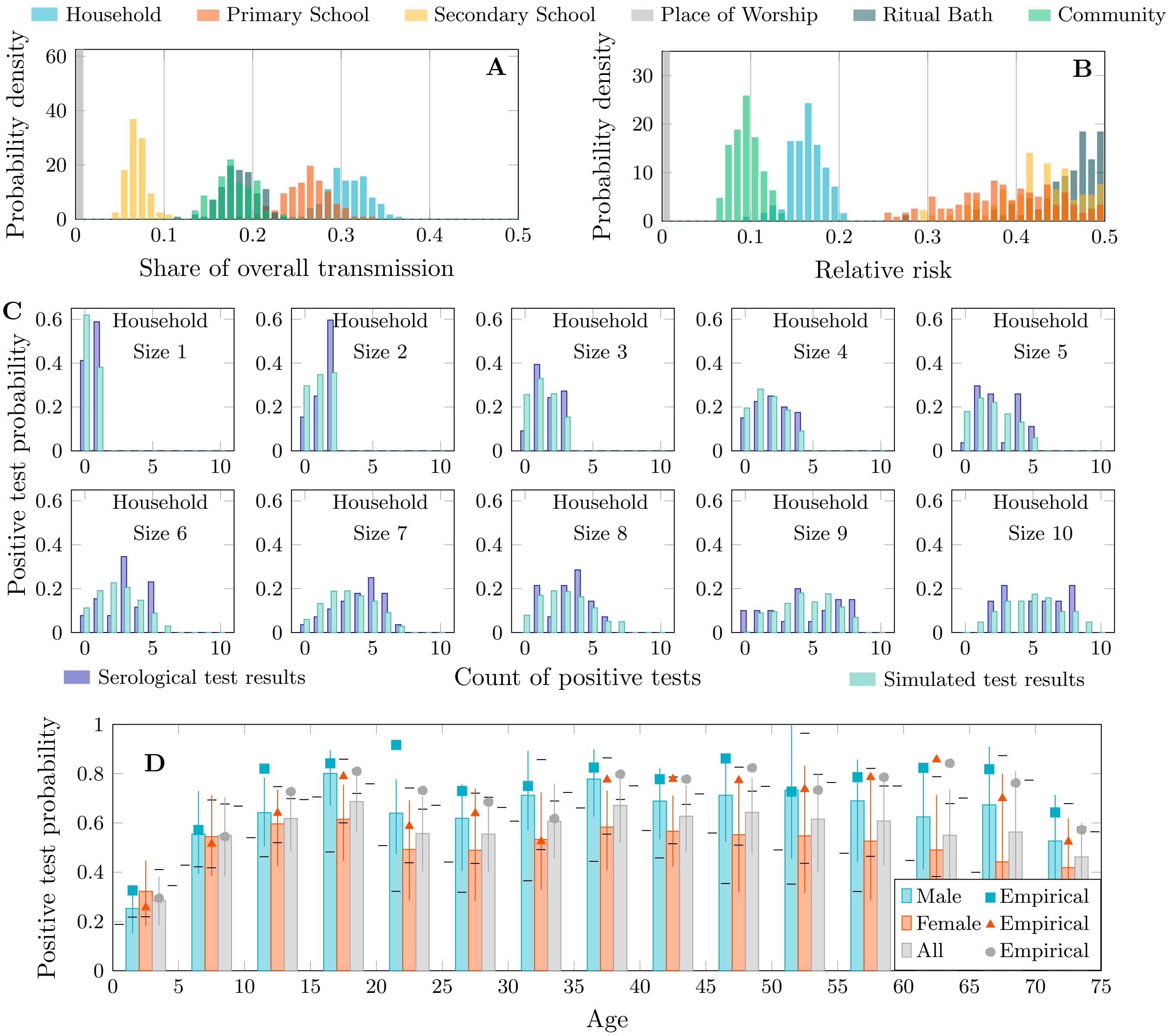


Figure 39: Transmission share, relative risk and postitive test distributions – Splitting places of worship (20th percentile)

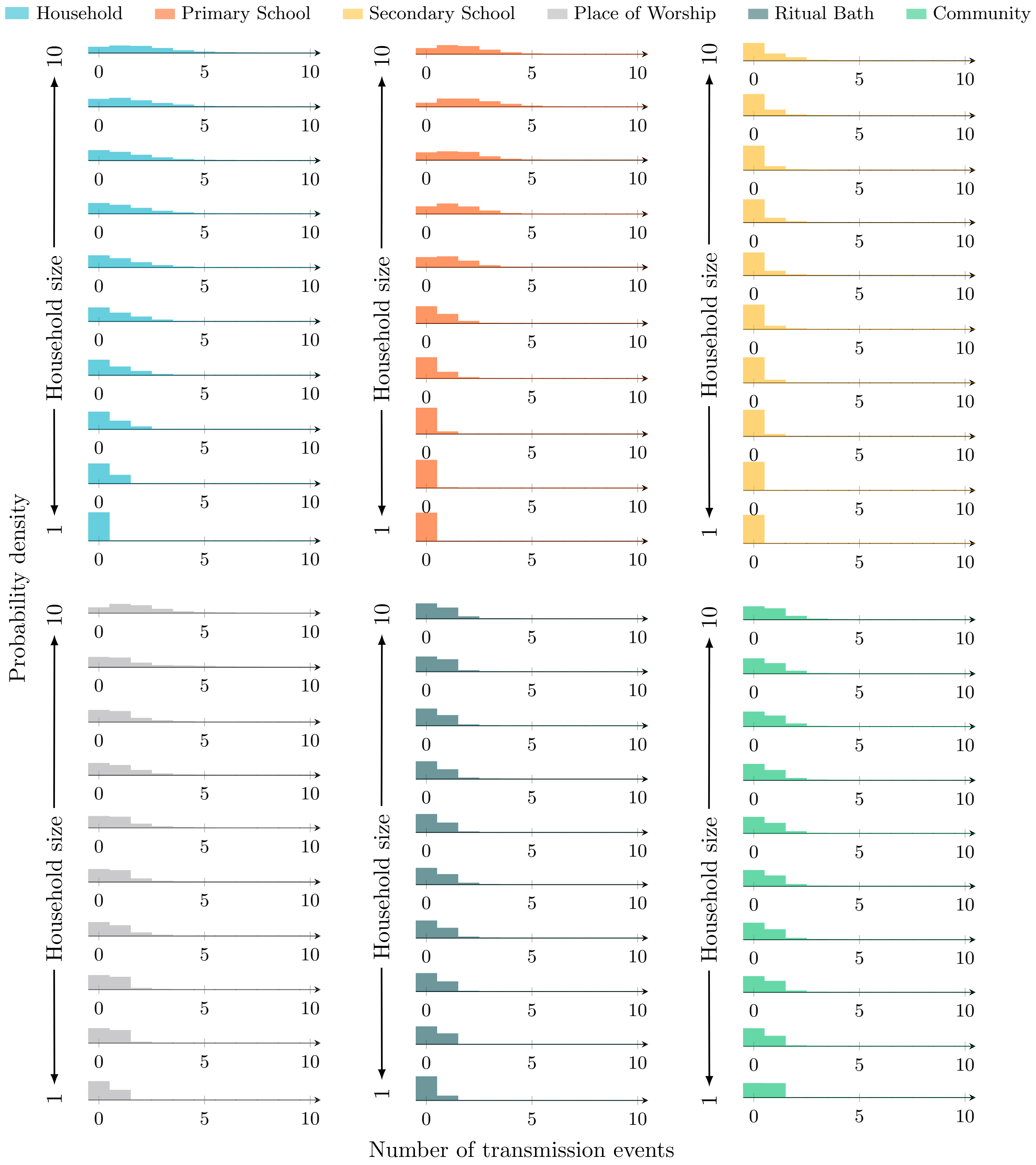


Figure 40: Transmission event distributions by source and household size – Splitting places of worship (20th percentile)

##### D.19 Splitting places of worship (10th percentile)

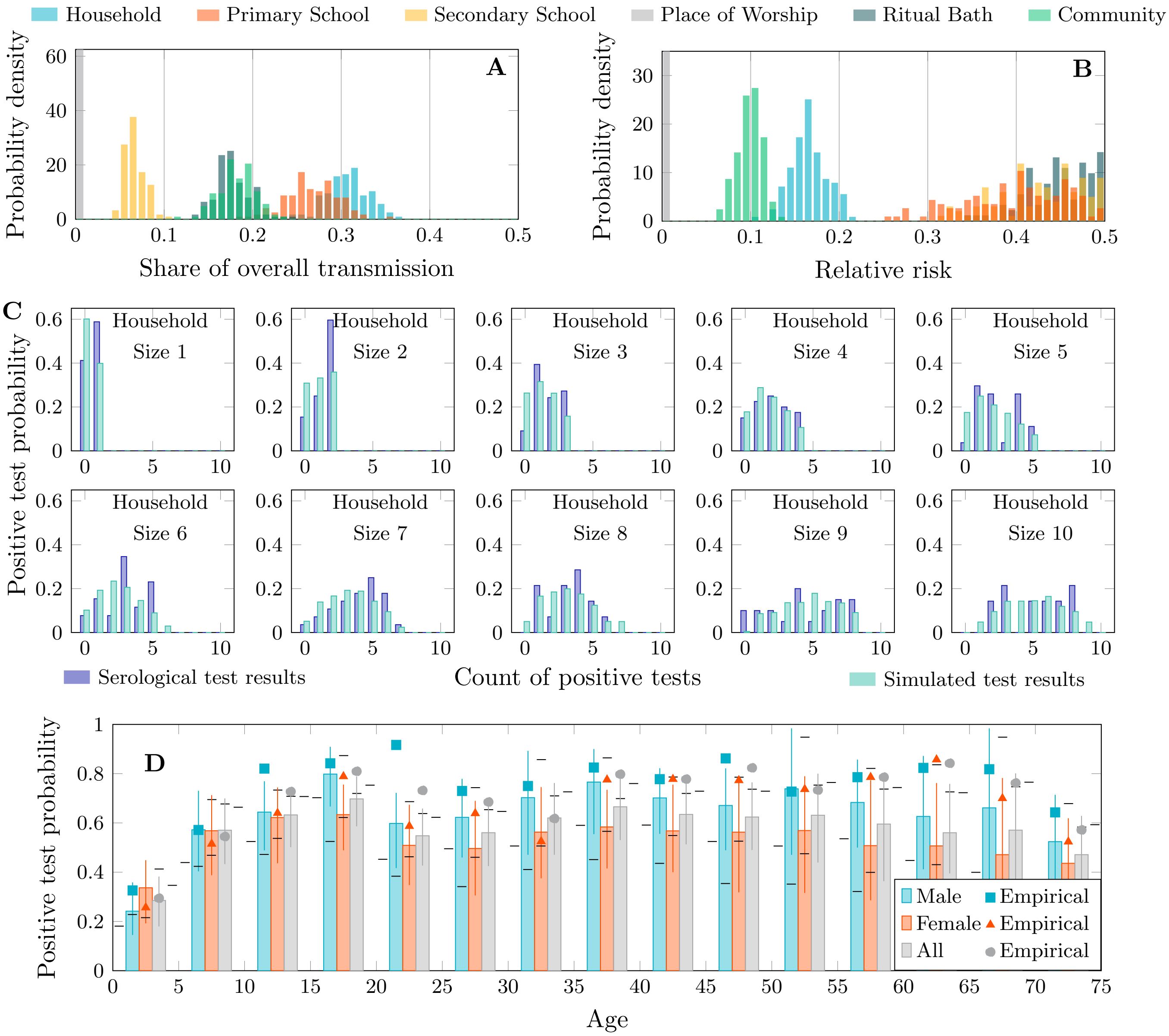


Figure 41: Transmission share, relative risk and postitive test distributions – Splitting places of worship (10th percentile)

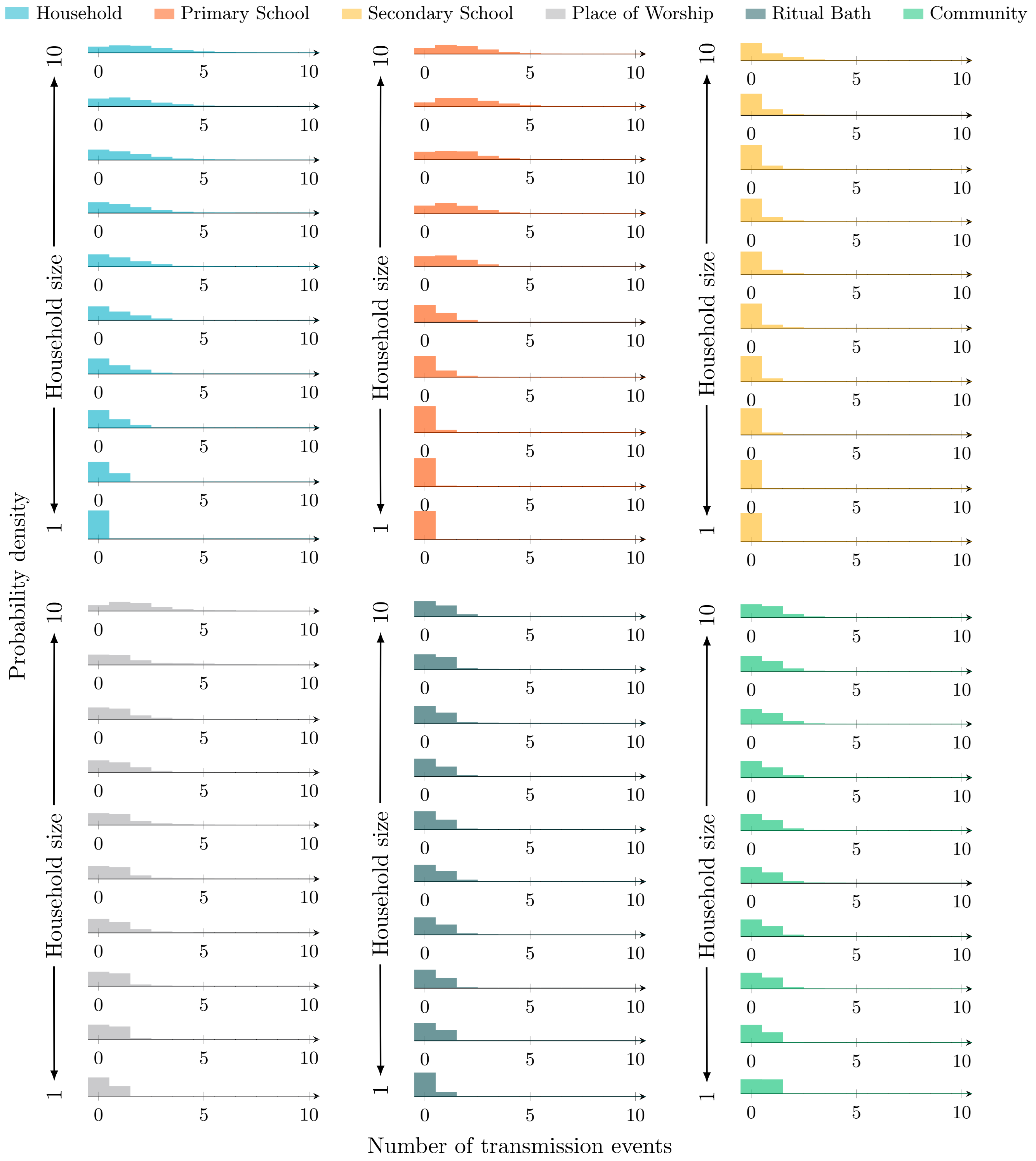


Figure 42: Transmission event distributions by source and household size – Splitting places of worship (10th percentile)

### E Sensitivity to network size

To ascertain the statistical power of the data, whether there is sufficient information in the bipartite network of the sampled size to estimate transmission dynamics in the entire community, we evaluate the sensitivity of the results to the network size. We both increase and decrease the network size. To decrease the size, we simply choose a fraction of households to delete. To increase the network size, we choose households at random and clone them, including the connections of individuals in the chosen households to the various places. We find that these perturbations, from 90% of network size to 120% do not materially affect the result. Statistics depicted in the figures are summarised in Table 3.

| **Setting** | **Transmission activity** | | **Relative risk** | |
|---|---|---|---|---|
| | Network 120% size | | | |
| household | 23.6% | [19.8-27.5%] | 15.8% | [12.6-19.2%] |
| primary school | 20.7% | [16.6-24.5%] | 40.3% | [30.9-48.3%] |
| secondary school | 5.0% | [3.4-6.9%] | 39.7% | [29.8-48.5%] |
| place of worship | 22.5% | [18.3-26.8%] | 39.0% | [31.6-46.1%] |
| ritual bath | 11.9% | [9.4-14.7%] | 39.7% | [32.5-47.3%] |
| community | 16.3% | [12.8-19.8%] | 11.0% | [8.1-13.9%] |
| | Network 110% size | | | |
| household | 23.9% | [19.9-27.7%] | 15.7% | [12.4-19.2%] |
| primary school | 20.7% | [16.5-24.5%] | 38.7% | [28.8-47.4%] |
| secondary school | 5.0% | [3.3-6.8%] | 39.2% | [29.6-48.5%] |
| place of worship | 22.9% | [18.6-27.4%] | 38.7% | [31.5-45.7%] |
| ritual bath | 12.2% | [9.6-15.2%] | 40.1% | [32.7-47.7%] |
| community | 15.3% | [11.9-18.9%] | 10.1% | [7.5-13.0%] |
| | Network 105% size | | | |
| household | 24.1% | [20.2-28.0%] | 15.9% | [12.5-19.3%] |
| primary school | 20.8% | [16.7-24.9%] | 38.9% | [29.3-47.9%] |
| secondary school | 5.0% | [3.2-7.0%] | 40.1% | [29.6-49.0%] |
| place of worship | 22.9% | [18.7-27.3%] | 38.2% | [31.1-44.8%] |
| ritual bath | 12.2% | [9.5-15.1%] | 39.7% | [31.8-47.1%] |
| community | 14.9% | [11.5-18.3%] | 9.8% | [7.1-12.4%] |
| | Network 99% size | | | |
| household | 24.3% | [20.2-28.3%] | 15.8% | [12.2-19.4%] |
| primary school | 20.8% | [16.6-24.9%] | 38.5% | [28.4-47.5%] |
| secondary school | 5.1% | [3.2-7.0%] | 41.1% | [30.8-49.4%] |
| place of worship | 23.1% | [18.9-27.4%] | 38.0% | [30.5-44.6%] |
| ritual bath | 12.4% | [9.6-15.5%] | 39.6% | [31.5-47.3%] |
| community | 14.3% | [10.9-17.7%] | 9.3% | [6.6-12.0%] |
| | Network 98% size | | | |
| household | 24.4% | [20.3-28.5%] | 15.9% | [12.5-19.5%] |
| primary school | 20.8% | [16.4-24.8%] | 38.7% | [28.6-47.5%] |
| secondary school | 5.0% | [3.1-7.1%] | 40.8% | [30.5-49.1%] |
| place of worship | 23.3% | [18.9-27.8%] | 38.3% | [31.2-45.0%] |
| ritual bath | 12.4% | [9.6-15.5%] | 39.5% | [31.8-47.3%] |
| community | 14.1% | [10.9-17.4%] | 9.2% | [6.6-11.8%] |
| | Network 95% size | | | |
| household | 24.5% | [20.5-28.5%] | 15.9% | [12.1-19.4%] |
| primary school | 20.9% | [16.4-25.1%] | 38.4% | [27.6-47.7%] |
| secondary school | 5.0% | [3.1-7.0%] | 41.1% | [30.0-49.5%] |

| | | | | |
|---|---|---|---|---|
| place of worship | 23.3% | [19.1-27.9%] | 38.2% | [30.8-45.2%] |
| ritual bath | 12.5% | [9.7-15.6%] | 39.6% | [31.7-47.7%] |
| community | 13.8% | [10.5-17.2%] | 8.9% | [6.5-11.5%] |
| | | Network 90% size | | |
| household | 24.7% | [20.7-28.9%] | 15.7% | [12.0-19.3%] |
| primary school | 20.8% | [16.4-25.1%] | 37.6% | [27.4-47.2%] |
| secondary school | 5.1% | [3.2-7.1%] | 40.5% | [29.4-49.2%] |
| place of worship | 23.5% | [19.1-28.2%] | 37.7% | [30.5-45.1%] |
| ritual bath | 12.7% | [9.8-15.9%] | 39.8% | [31.9-47.5%] |
| community | 13.1% | [9.9-16.6%] | 8.3% | [5.8-11.1%] |

Table 3: Sensitivity to network size: transmission activity and relative risk

#### E.1 120% network size

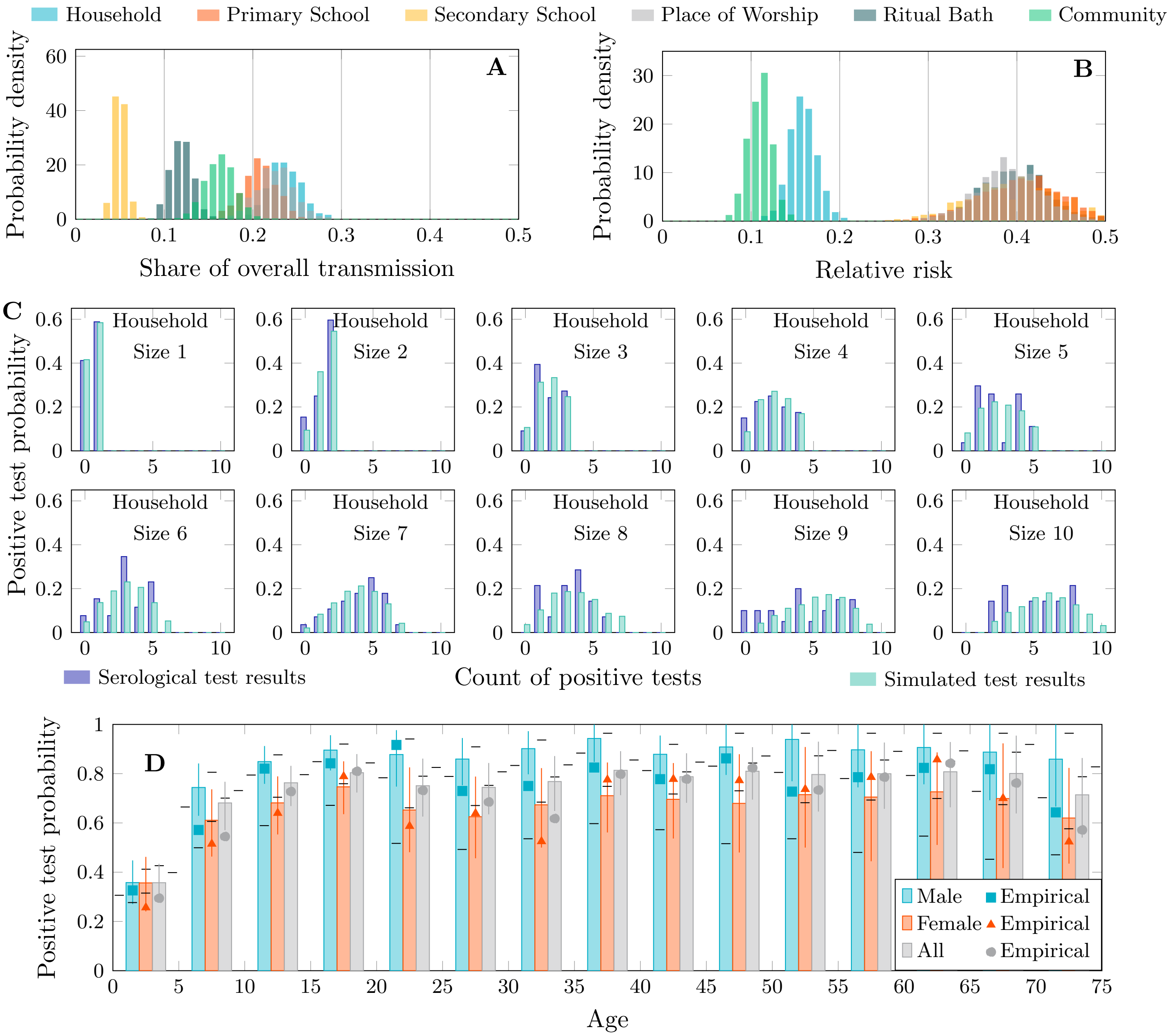


Figure 43: Transmission share, relative risk and postitive test distributions – 120% network size

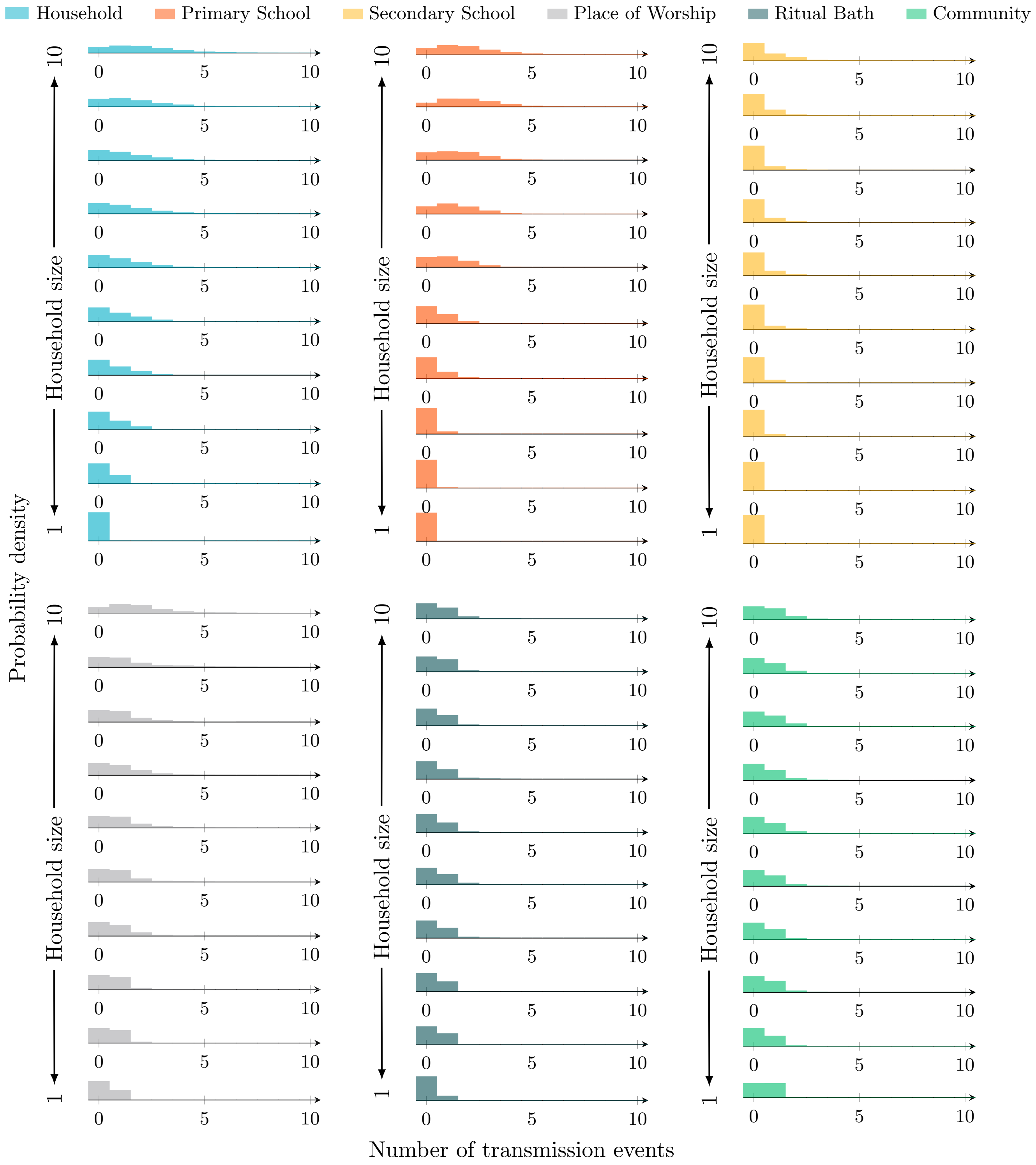


Figure 44: Transmission event distributions by source and household size – 120% network size

#### E.2 110% network size

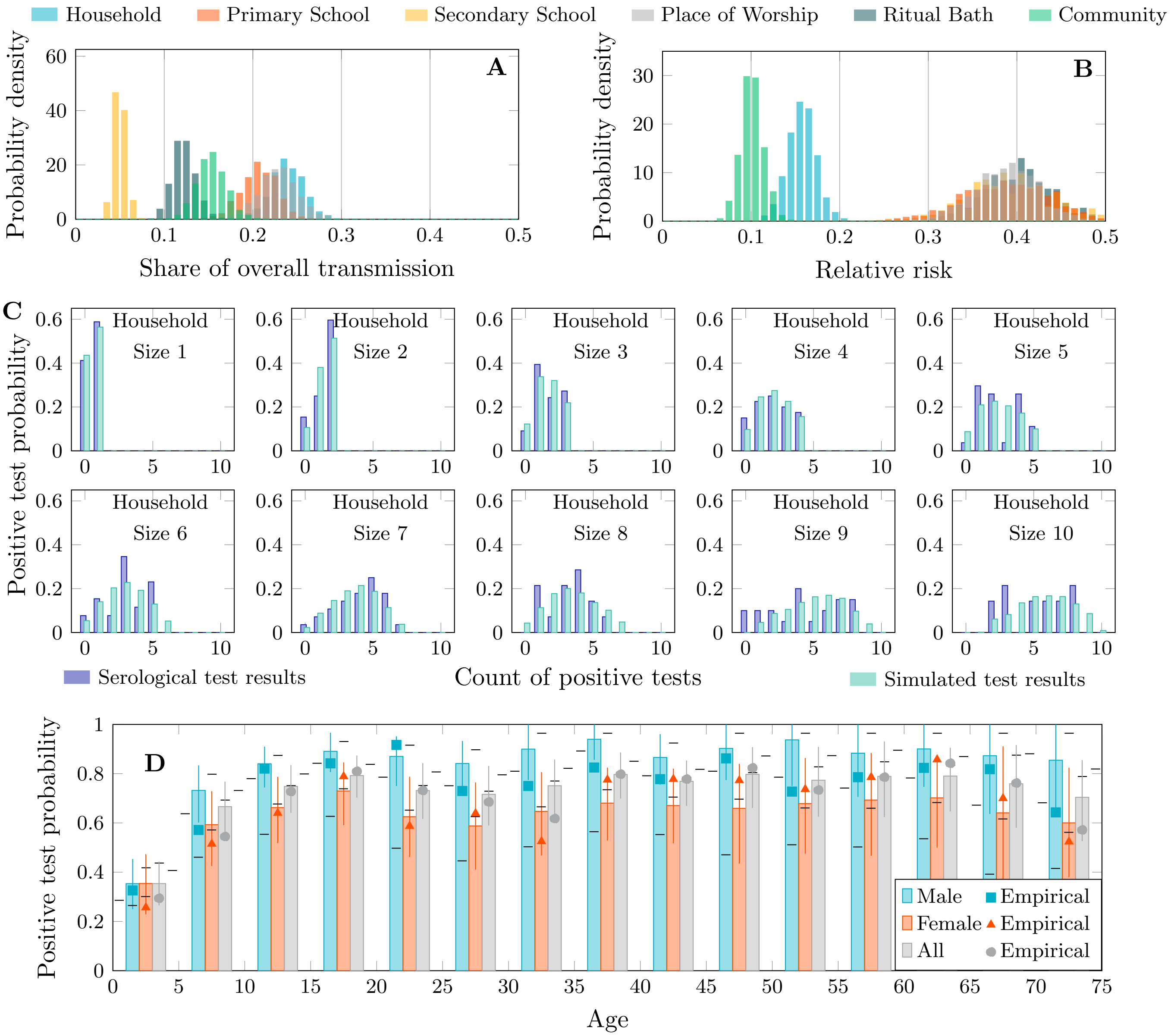


Figure 45: Transmission share, relative risk and postitive test distributions – 110% network size

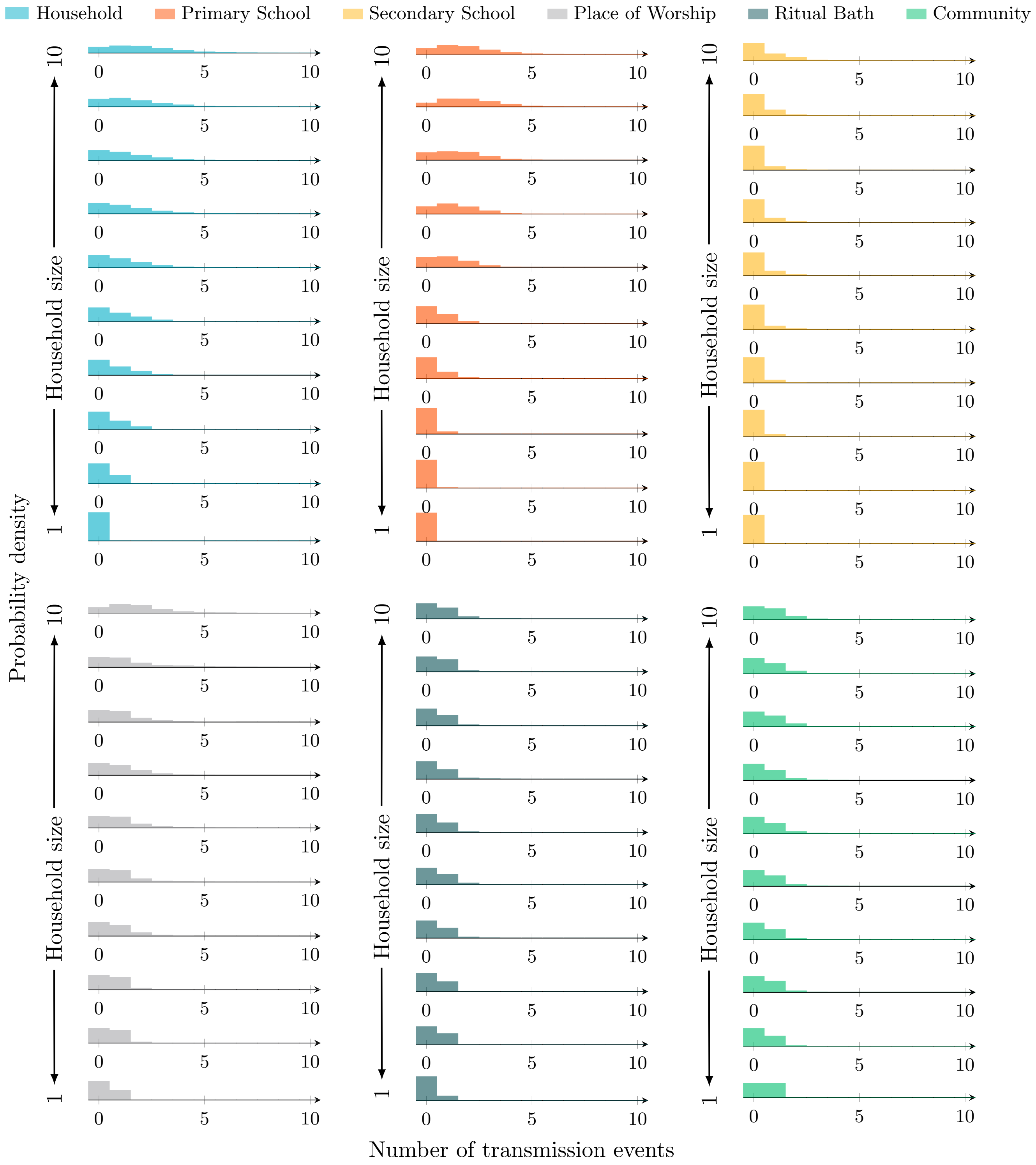


Figure 46: Transmission event distributions by source and household size – 110% network size

#### E.3 105% network size

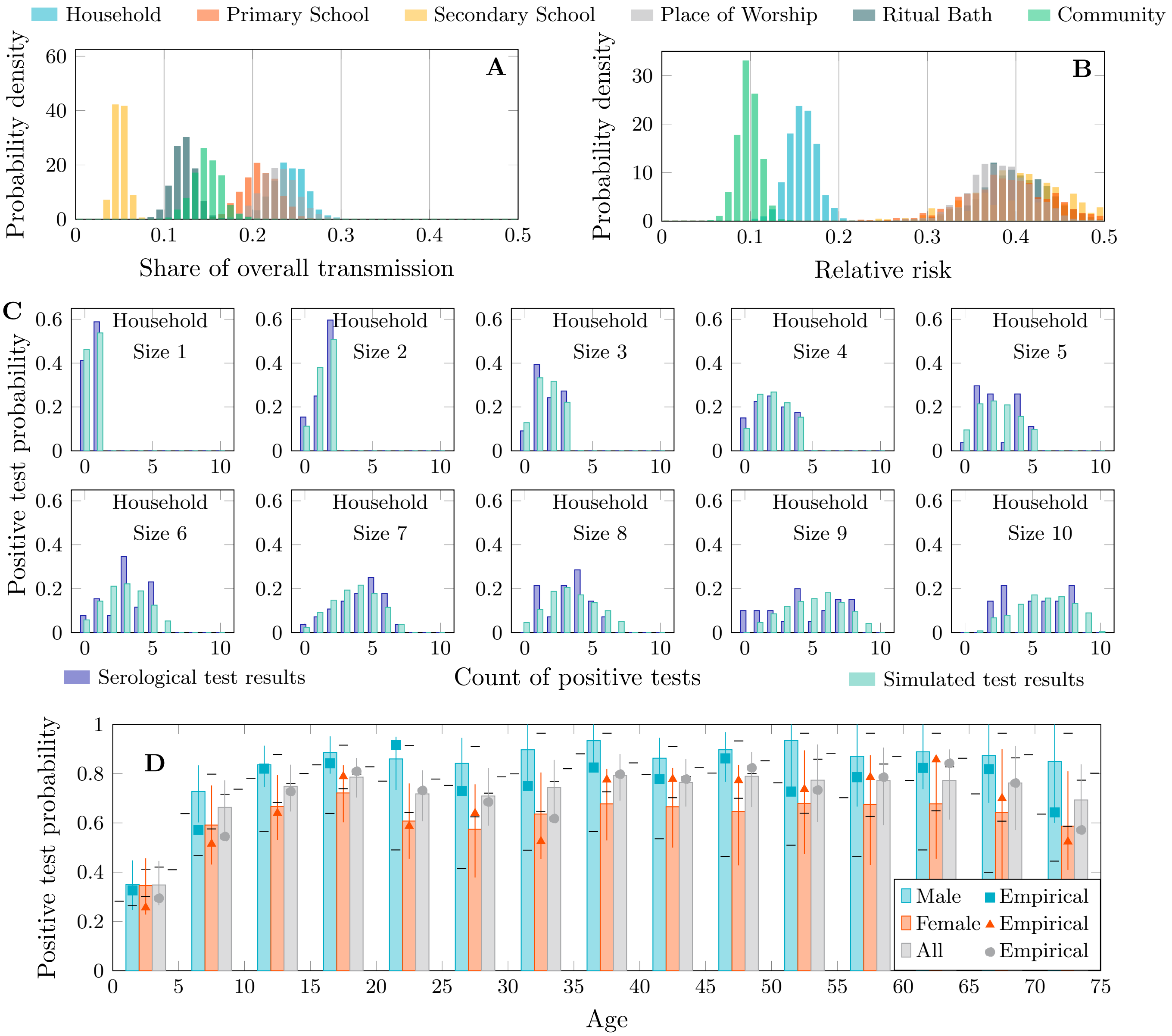


Figure 47: Transmission share, relative risk and postitive test distributions – 105% network size

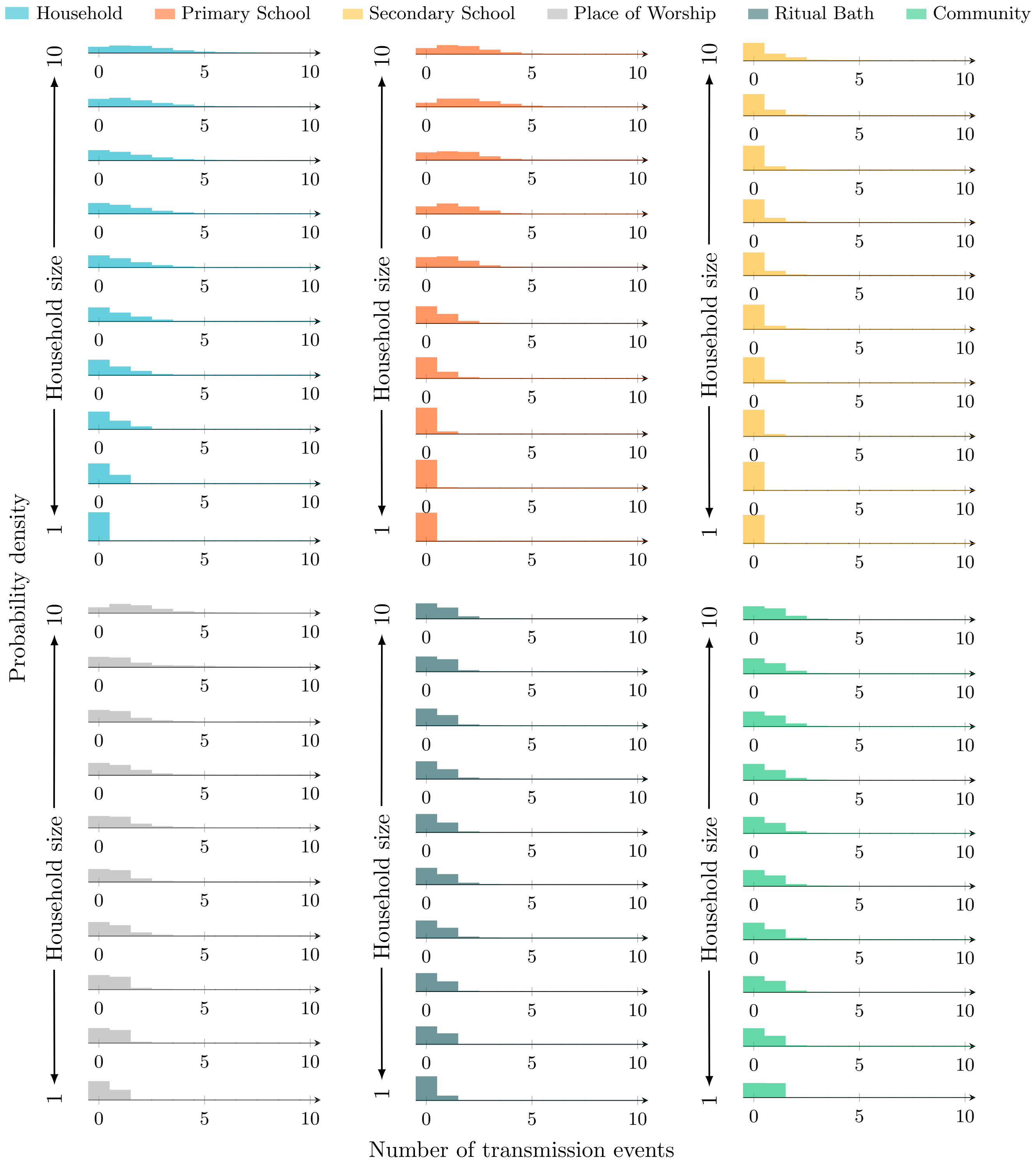


Figure 48: Transmission event distributions by source and household size – 105% network size

#### E.4 99% network size

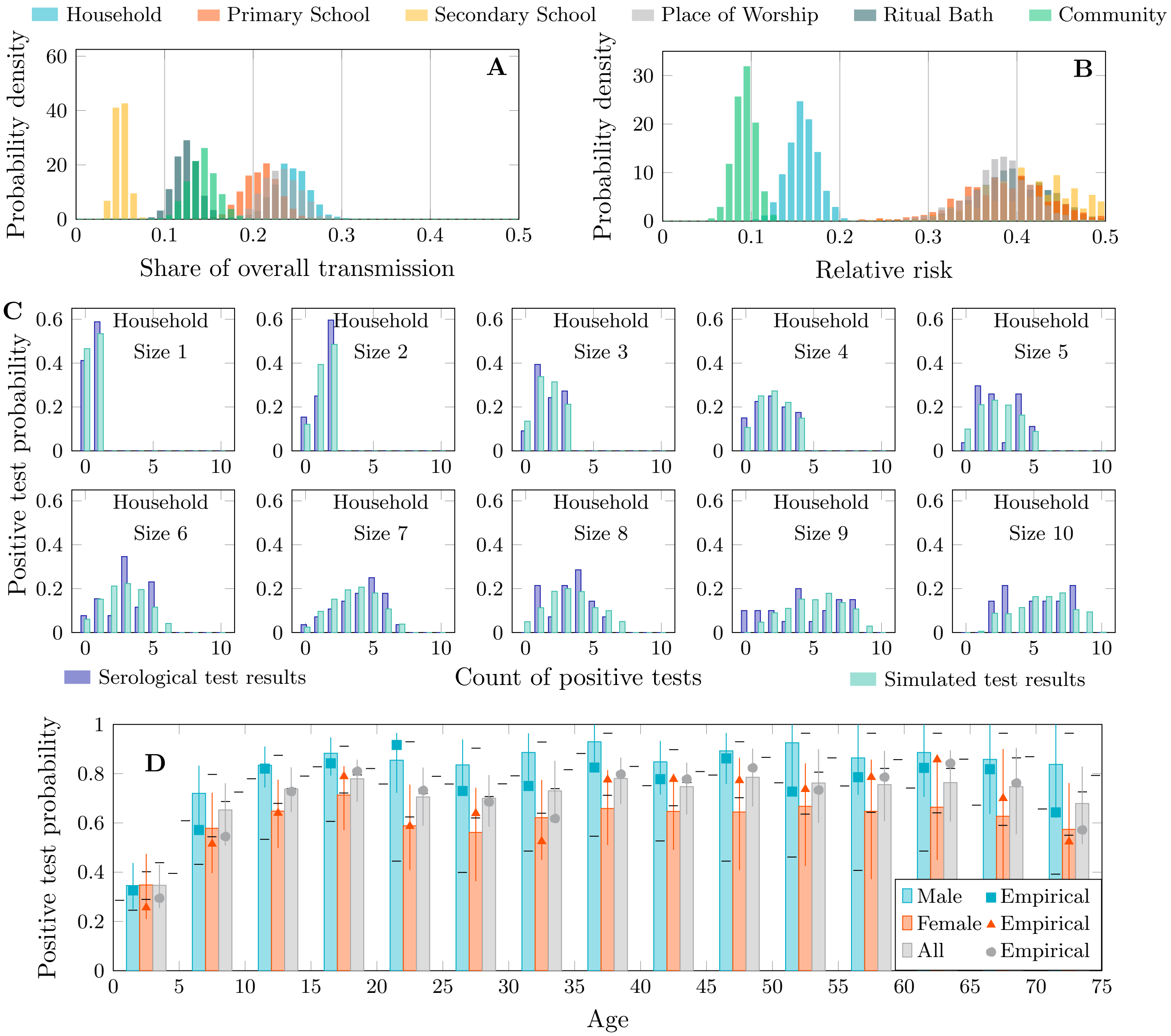


Figure 49: Transmission share, relative risk and postitive test distributions – 99% network size

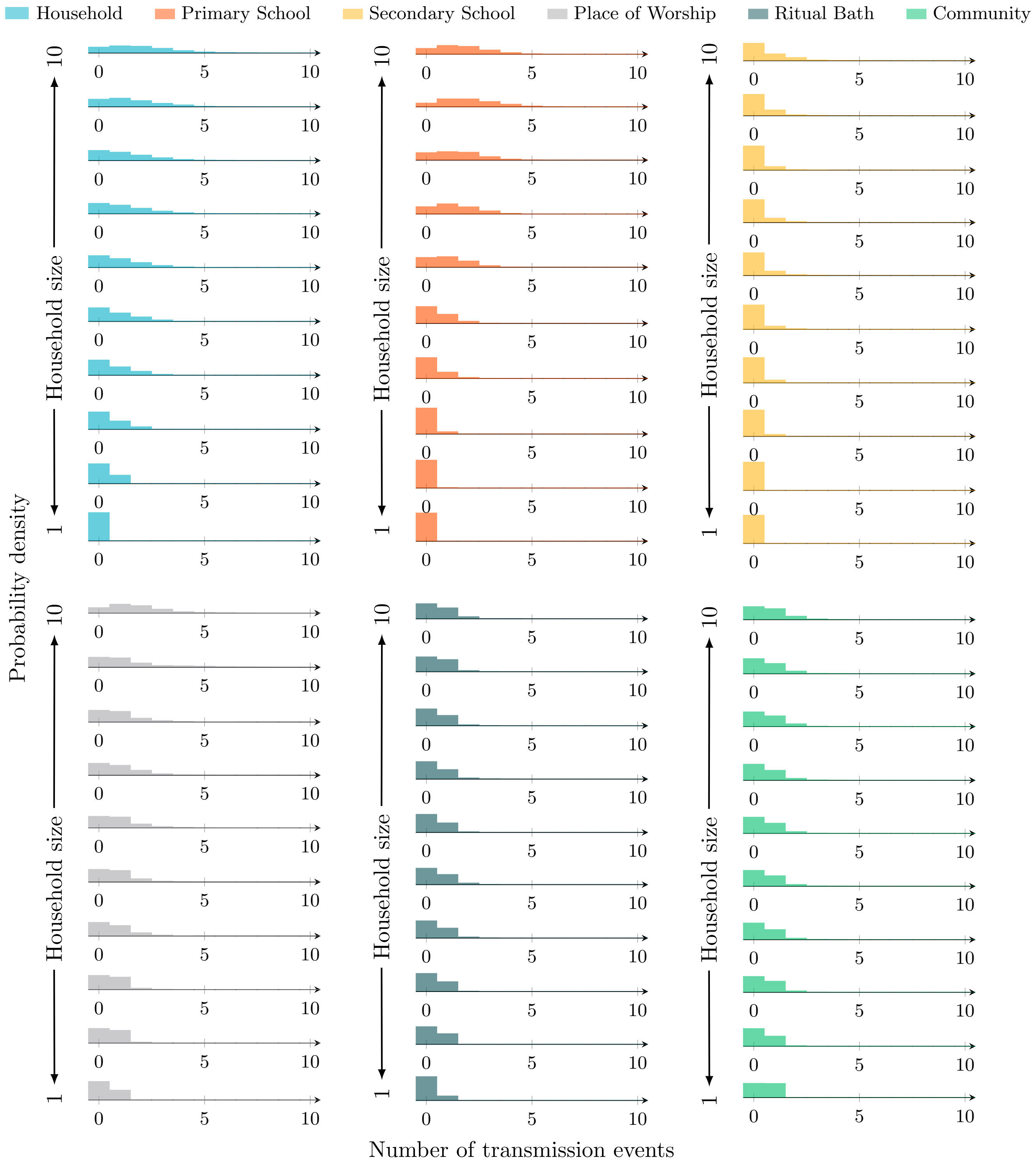


Figure 50: Transmission event distributions by source and household size – 99% network size

##### E.5 98% network size

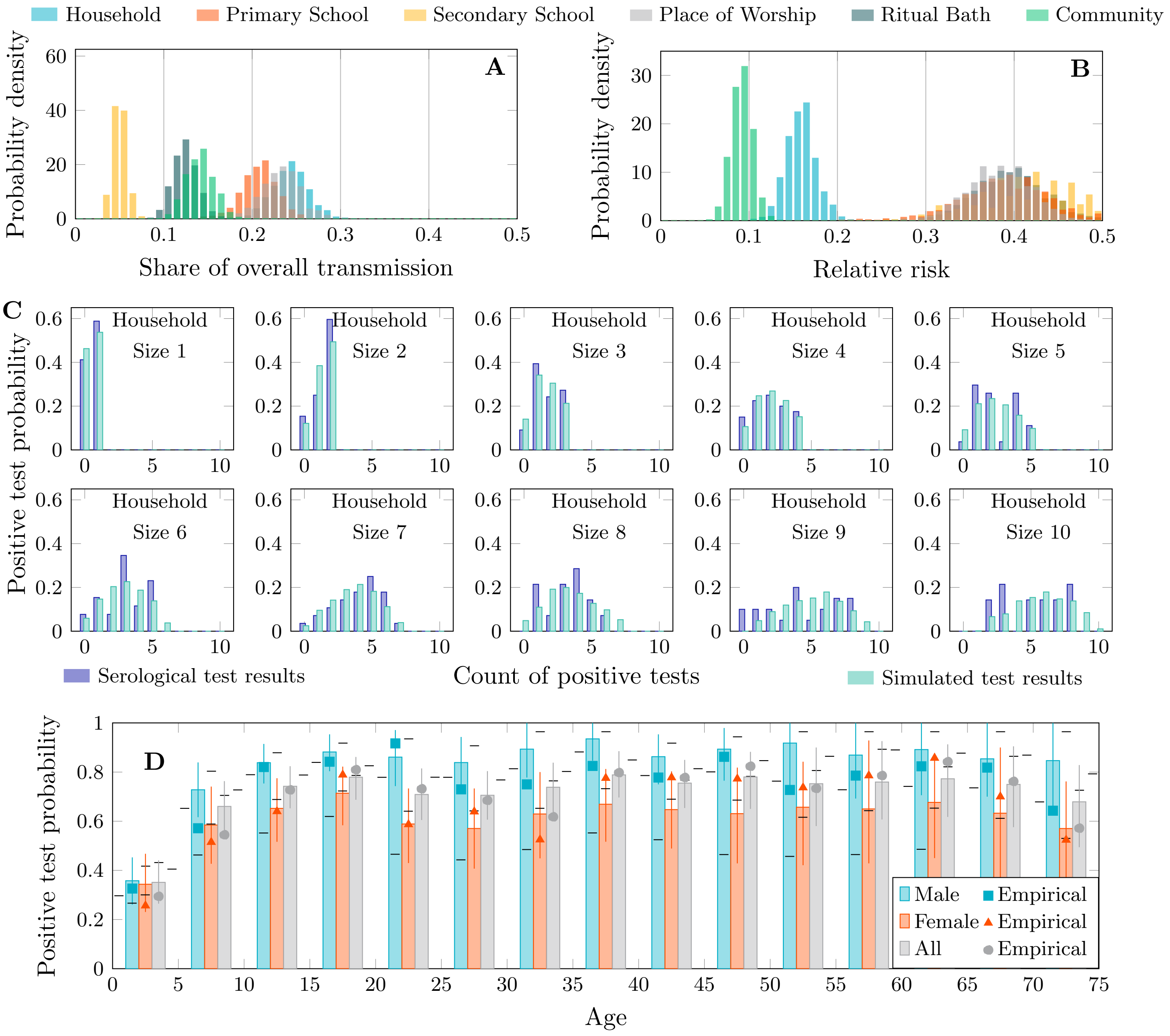


Figure 51: Transmission share, relative risk and postitive test distributions – 98% network size

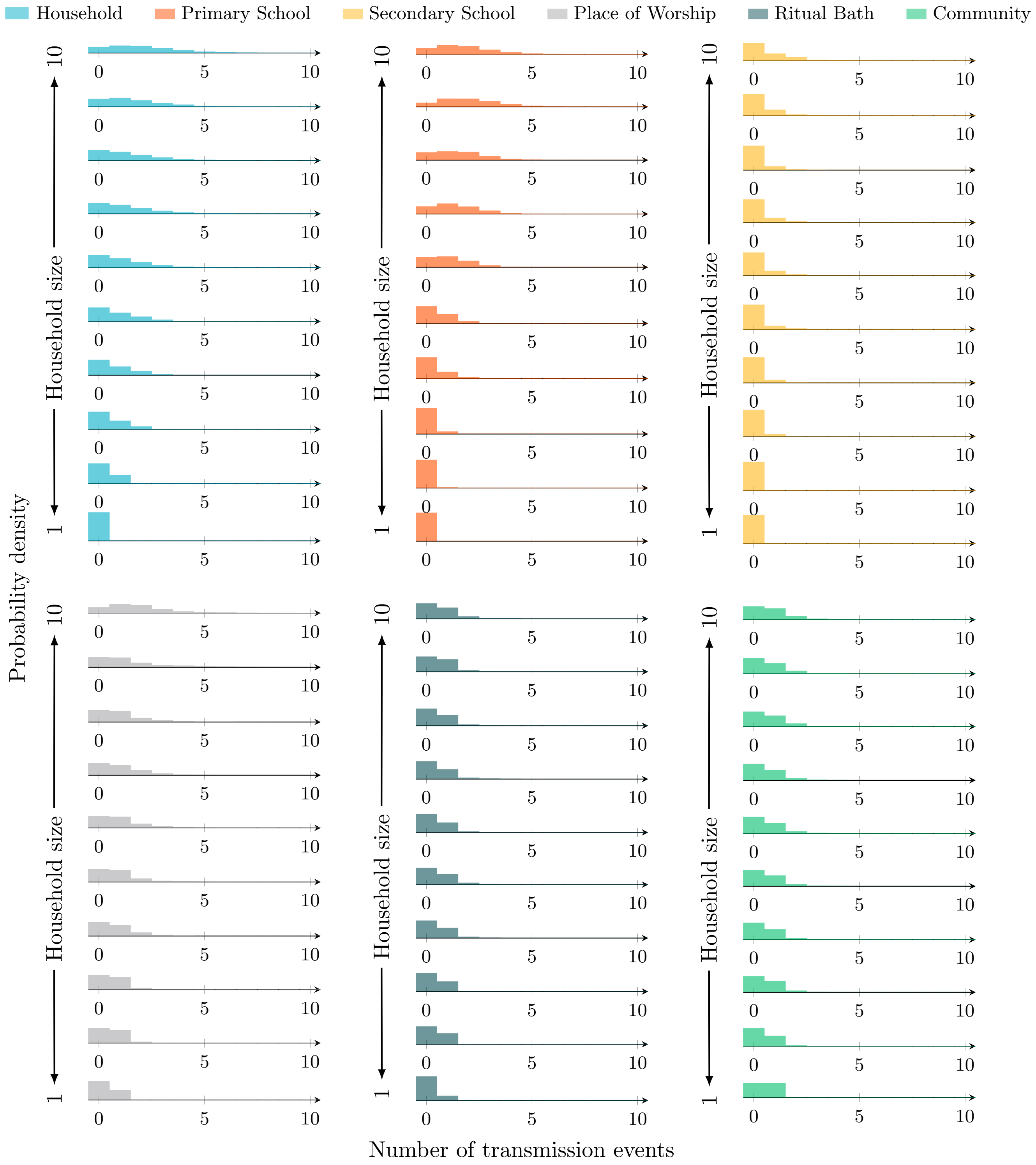


Figure 52: Transmission event distributions by source and household size – 98% network size

#### E.6 95% network size

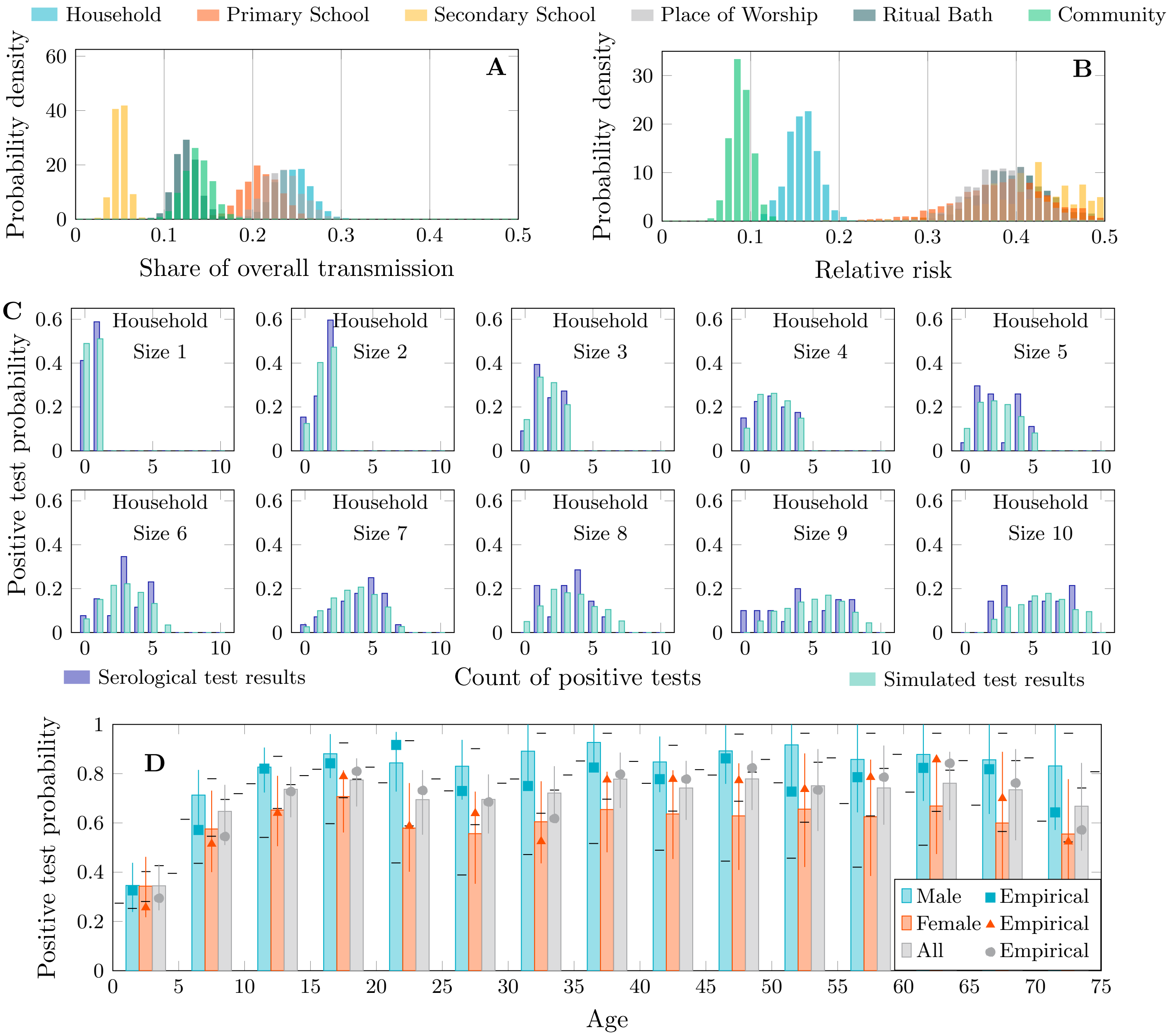


Figure 53: Transmission share, relative risk and postitive test distributions – 95% network size

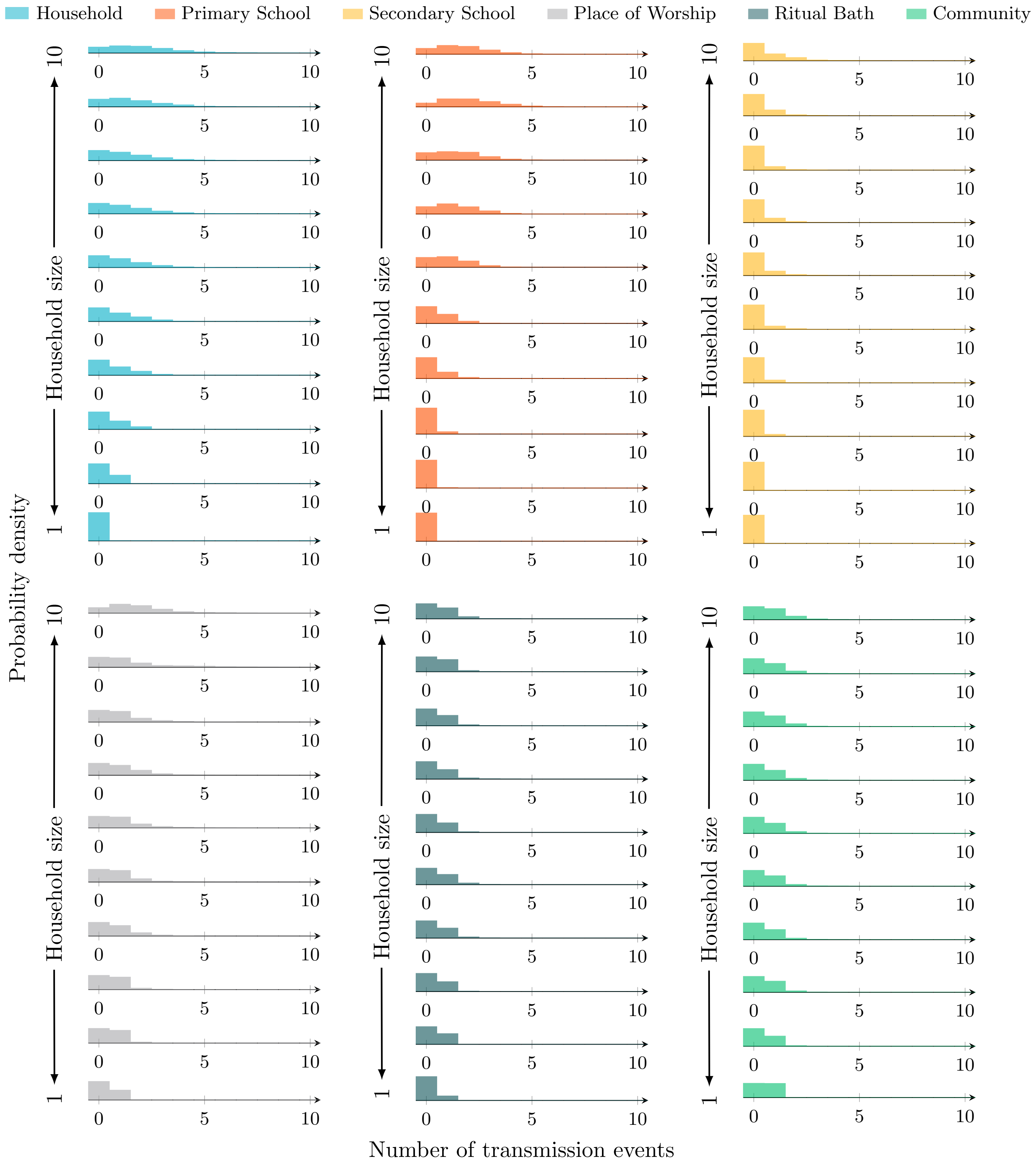


Figure 54: Transmission event distributions by source and household size – 95% network size

##### E.7 90% network size

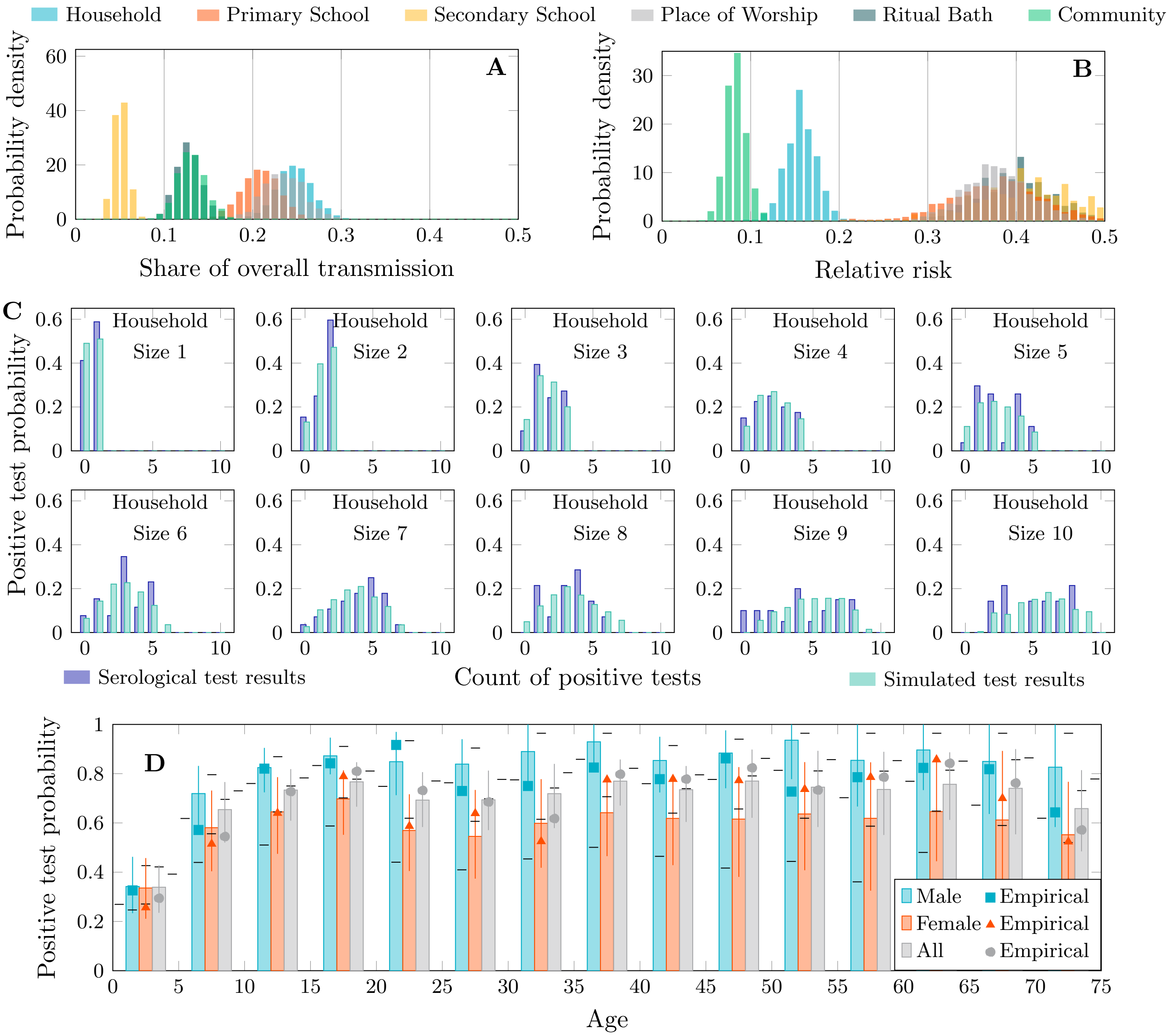


Figure 55: Transmission share, relative risk and postitive test distributions – 90% network size

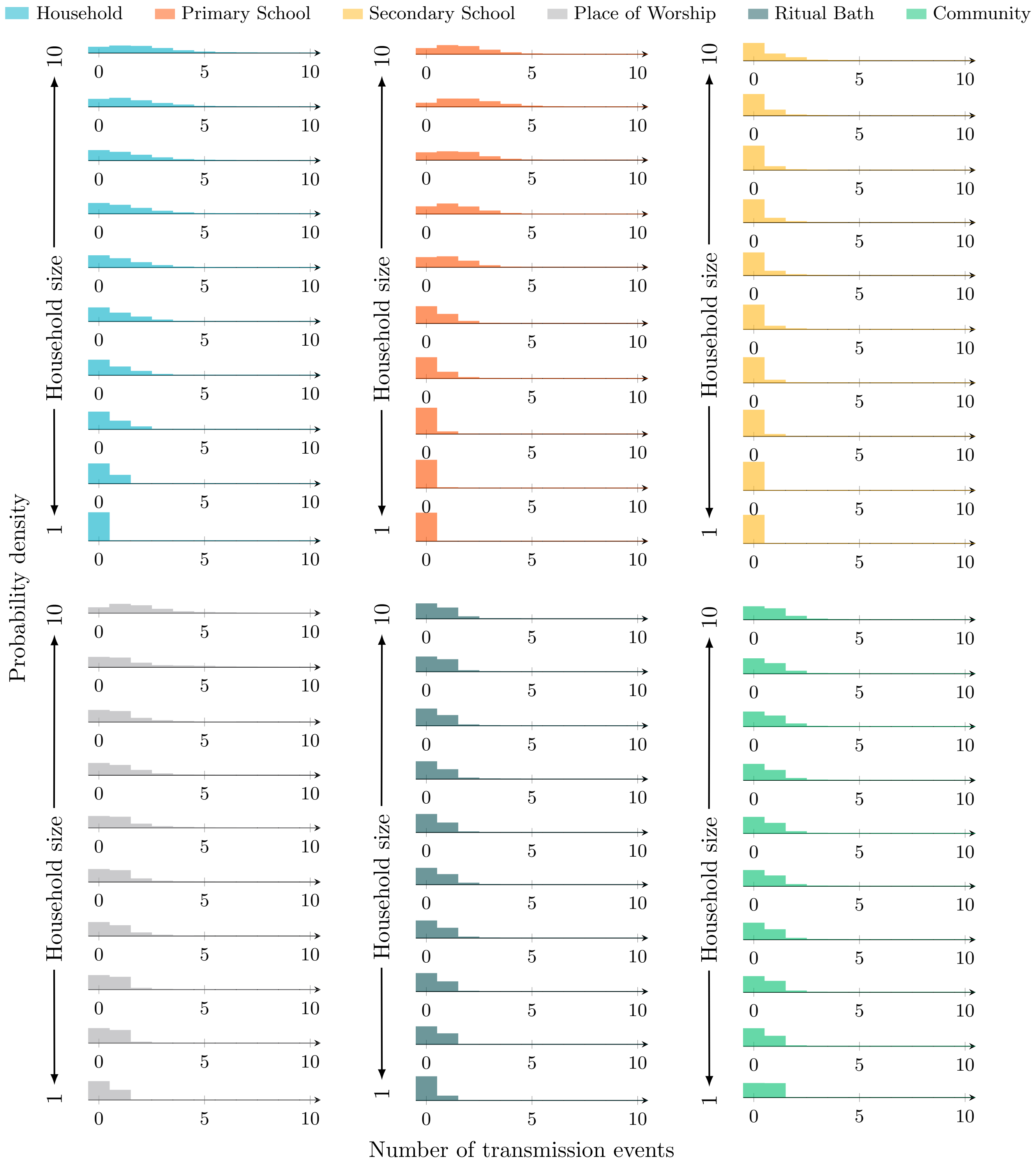


Figure 56: Transmission event distributions by source and household size – 90% network size

### F Transmission Model

This is the model as simulated. It is lightly modified from the actual model used: variable names have been changed to reflect community sensitivity about identification of specific religious practices.

```
// -*- mode: kappa -*-

// rate of progression from exposed to infectious
%var:   alpha   0.2
// rate of recovery or removal
%var:   gamma   0.15

// a person with a disease progression state, and a site
// representing the location of interactions, and a site
// i for recording the transmission setting
%agent: person(c{s e i r}, i, loc)
%agent: household(loc)
%agent: worship(loc)
%agent: primary(loc)
%agent: secondary(loc)
%agent: bath(loc)

// a bipartite graph is externally supplied, but we give it
// a name here. people are in one partition, places are in
// the other
%graph: g

// standard rules for disease progression
'progression' person(c{e}) -> person(c{i}) @ alpha
'removal'     person(c{i}) -> person(c{r}) @ gamma

// places partition the population. assume that interaction
// within each partition is well-mixed.
//
// read these rules as, a place causes infection to a susceptible
// person in that place proportionally to the fraction of
// infectious people in that place.
//
// the transmission probabilities per unit time for each place
// beta_h, beta_g, beta_p, beta_s and beta_m, are supplied
// separately because we allow them to vary for fitting
//
// the transmission rules also record the setting where the
// infection happened (the i{} site) for post-hoc analysis.
'infection_household' household(loc[1])[p], person(c{s}, i{none}, loc[1])[q] \
                   -> household(loc[1]),    person(c{e}, i{household}, loc[1]) \
                    @ beta_h*inf(g,p)*sus(g,q)

'infection_worship' worship(loc[1])[p], person(c{s}, i{none}, loc[1]) \
                 -> worship(loc[1]),    person(c{e}, i{worship}, loc[1]) \
                  @ beta_g*inf(g,p)

'infection_primary' primary(loc[1])[p], person(c{s}, i{none}, loc[1]) \
                 -> primary(loc[1]),    person(c{e}, i{primary}, loc[1]) \
                  @ beta_p*inf(g,p)

'infection_secondary' secondary(loc[1])[p], person(c{s}, i{none}, loc[1]) \
                   -> secondary(loc[1]),    person(c{e}, i{secondary}, loc[1])
                    @ beta_s*inf(g,p)

'infection_bath' bath(loc[1])[p], person(c{s}, i{none}, loc[1]) \
```

```
               -> bath(loc[1]),    person(c{e}, i{bath}, loc[1]) \
                @ beta_m*inf(g,p)

// these are well-mixed rules, the *> symbol signals to the
// simulator to not enumerate all possible embeddings but
// simply to choose agents on the left-hand side at random
// as with a petri net
//
// separate rules for each lifecycle phase for variable
// susceptibility
'infection_c_preschool' person(c{s}, i{none}, lifecycle{child}), person(c{i}) \
                      *> person(c{e}, i{community}, lifecycle{child}), person(c{i}) \
                       @ s_preschool*beta_e/1942
'infection_c_primary' person(c{s}, i{none}, lifecycle{primary}), person(c{i}) \
                    *> person(c{e}, i{community}, lifecycle{primary}), person(c{i}) \
                     @ s_primary*beta_e/1942
'infection_c_secondary' person(c{s}, i{none}, lifecycle{secondary}), person(c{i}) \
                      *> person(c{e}, i{community}, lifecycle{secondary}), person(c{i}) \
                       @ s_secondary*beta_e/1942
'infection_c_adult' person(c{s}, i{none}, lifecycle{adult}), person(c{i}) \
                  *> person(c{e}, i{community}, lifecycle{adult}), person(c{i}) \
                   @ beta_e/1942

// an additional well-mixed rule for adult females due to
// lack of data about their associations with places
'infection_c_female' person(c{s}, i{none}, lifecycle{adult}, sex{female}), person(c{i}) \
                   *> person(c{e}, i{community}, lifecycle{adult}, sex{female}), person(c{i}) \
                    @ beta_f/1942

// observables corresponding to standard infectious disease
// modelling "compartments"
%obs: S |person(c{s})|
%obs: E |person(c{e})|
%obs: I |person(c{i})|
%obs: R |person(c{r})|
// observables for infectious and recovered individuals by
// sex
%obs: Mi |person(sex{male}, c{i})|
%obs: Fi |person(sex{female}, c{i})|
%obs: Mr |person(sex{male}, c{r})|
%obs: Fr |person(sex{female}, c{r})|
// observable for the distance measure of simulation state
// to the empirical data
%obs: Dist emsar(g, "household")+sbal(g)
// observables for rule activity, counting the number of time
// each rule has been used
%obs: ACTH  activity(infection_household)
%obs: ACTG  activity(infection_worship)
%obs: ACTP  activity(infection_primary)
%obs: ACTS  activity(infection_secondary)
%obs: ACTM  activity(infection_bath)
%obs: ACTE  activity(infection_c_preschool)+activity(infection_c_primary)+activity(infection_c_secondary)+activity(infection_c_adult)+activity(infection_c_female)

// unlike regular kappa, these values are the fraction of the
// nodes in the supplied graph that should have their initial
// states set according to this specification
%init: 0.99 person(c{s}, i{none})
%init: 0.01 person(c{i}, i{init})
```
